# Trans-ethnic genome-wide meta-analysis of 35,732 cases and 34,424 controls identifies novel genomic cross-ancestry loci contributing to lung cancer susceptibility

**DOI:** 10.1101/2020.10.06.20207753

**Authors:** Jinyoung Byun, Younghun Han, Yafang Li, Jun Xia, Xiangjun Xiao, Ryan Sun, Kyle M. Walsh, Ivan Gorlov, Olga Gorlova, Wen Zhou, Rowland W Pettit, Zhuoyi Song, Susan M Rosenberg, Dakai Zhu, John K. Wiencke, Demetrius Albanes, Stephan Lam, Adonina Tardon, Chu Chen, Gary Goodman, Stig Bojeson, Hermann Brenner, Maria Teresa Landi, Mattias Johansson, Thomas Mulley, Angela Risch, H-Erich Wichmann, Heike Bickeböller, David C. Christiani, Gad Rennert, Susanne Arnold, John K. Field, Sanjay Shete, Loic LeMarchand, Ole Melander, Hans Brunnstrom, Geoffrey Liu, Angeline S. Andrew, Lambertius A. Kiemeney, Hongbing Shen, Shan Zienolddiny, Kjell Grankvist, Mikael Johansson, Neil Caporaso, Angela Cox, Yun-Chul Hong, Jian-Min Yuan, Philip Lazarus, Matthew B. Schabath, Melinda C. Aldrich, Apla Patel, Qing Lan, Nat Rothman, Fiona Taylor, Margaret Spitz, Paul Brennan, Xihong Lin, James McKay, Rayjean J. Hung, Christopher I. Amos, INTEGRAL Consortium

**Author notes:** These authors have equal contributions (Jinyoung Byun, Younghun Han, Yafang Li, Jun Xia). Corresponding Author: Christopher I. Amos, Ph.D., Institute for Clinical and Translational Research, Baylor College of Medicine, Houston, TX, USA. **Conflict of Interest:** All authors declare that they have no conflict of interest.

## Abstract

Lung cancer is the leading cause of cancer death worldwide. Genome-wide association studies have revealed genetic risk factors, highlighting the role of smoking, family history, telomere regulation, and DNA damage-repair in lung cancer etiology. Many studies have focused on a single ethnic group to avoid confounding from variability in allele frequencies across populations; however, comprehensive multi-ethnic analyses may identify variants that are more likely to be causal. This large-scale, multi-ethnic meta-analyses identified 28 novel risk loci achieving genome-wide significance. Leading candidates were further studied using single-cell methods for evaluating DNA-damage. DNA-damage promoting activities were confirmed for selected genes by knockdown genes and overexpression studies.

## INTRODUCTION

Lung cancer is a multifactorial disease driven by environmental exposures, especially cigarette smoking, inherited germline genetic variants, and an accumulation of somatic genetic events^1^. Although genome-wide association studies (GWAS) have identified many significant contributing risk loci, the genetic underpinnings of lung cancer according to racial and or ethnic disparities remains incompletely understood^2-6^. Most GWAS have focused on genetically homogeneous case-control studies from European-ancestry populations^7^. Trans-ethnic studies have been useful in examining the heritability of common quantitative traits and the genetic architecture of complex diseases (e.g. type II diabetes)^7-9^.

Trans-ethnic meta-analysis of genome-wide association studies have been used to increase study power by increasing the total study sample size. Also, trans-ethnic analysis may improve signal detection for rare variants if they are more frequent in one population and for causal variants if there is variability in linkage disequilibrium (LD) among causal variants and marker alleles among populations ^10^.

Consistency in effects among populations further supports causal effects^7,8^.

In the past two decades, about 45 GWAS of lung cancer susceptibility have been published^2,3,11,12^. The heritability of lung cancer attributable to genetic factors is estimated at 12-21%^1,6,13^. Ethnic differences in the incidence of lung cancer suggest underlying heterogeneity in the genetic architecture of lung cancer among human populations. Building on the recently completed OncoArray lung cancer GWAS^12,14-19^, we performed a trans-ethnic GWAS meta-analysis with the goal of comprehensively characterizing common and rare lung cancer genetic susceptibility loci across multiple ancestral populations. We expected that combining GWAS summary data across populations of diverse ancestry would improve the power and precision for detecting associations for lung cancer development and differences in LD structure could be exploited to identify causal variants driving the observed associations with common or rare single nucleotide polymorphisms (SNPs)^18,20^.

## RESULTS

### Association analyses of lung cancer

We included 70,156 individuals (35,732 cases and 34,424 controls) from 12 studies of ethnically diverse populations. Most individuals were inferred as having European descent (74%), with 18% having Asian descent and 8% having African descent^21^. Prior to association analyses, all samples from 12 studies were imputed using 32,470 samples from the Haplotype Reference Consortium (HRC)^22^ as a reference panel. Detailed quality control processes are described in Online Methods. We conducted ancestry-stratified analyses (Table 1 and Supplementary Tables 1) and then meta-analyzed GWAS results from the stratified populations of European (CEU), Asian (CHB), and African (YRI) ancestries^21^. We also performed a cross-ancestry fixed-effect meta-analyses to detect additional loci associated with predominant histological types; lung adenocarcinoma (ADE), squamous cell lung carcinoma (SQC), and small cell lung carcinoma (SCC) (Supplementary Table 1). There were no genomic inflations for lung cancer and any histologic subtypes (Figure 1) suggesting no residual population stratification for each of the ethnicity-stratified analysis or combined analyses.

**Table 1.** Description of individual samples after entire GWAS quality control process.

| Study | Ethnicity (Case/Control) |  |  |
| --- | --- | --- | --- |
|  | European | Asian | African |
| **AFFY** | 1358/2580 | 48/80 | 6/18 |
| **FLCCA** | 0/0 | 4,756/3,688 | 0/0 |
| **GELCC** | 740/781 | 3/3 | 6/5 |
| **EAGLE** | 1,919/1,972 | 0/0 | 0/0 |
| **GERMAN** | 419/471 | 0/0 | 0/0 |
| **IARC** | 1,829/2,429 | 0/0 | 0/0 |
| **ICR** | 1,366/1,627 | 0/0 | 0/0 |
| **4MDACC** | 1,146/1,129 | 0/0 | 0/1 |
| **NCI** | 5/13 | 1/5 | 1,704/3,469 |
| **ONCO** | 17,417/13,599 | 2,254/1,596 | 271/286 |
| **PLCO** | 159/241 | 0/0 | 0/0 |
| **SLRI** | 325/436 | 0/0 | 0/0 |
| **Total** | **26,683/25,278** | **7,062/5,372** | **1,987/3,779** |

**Figure 1.**
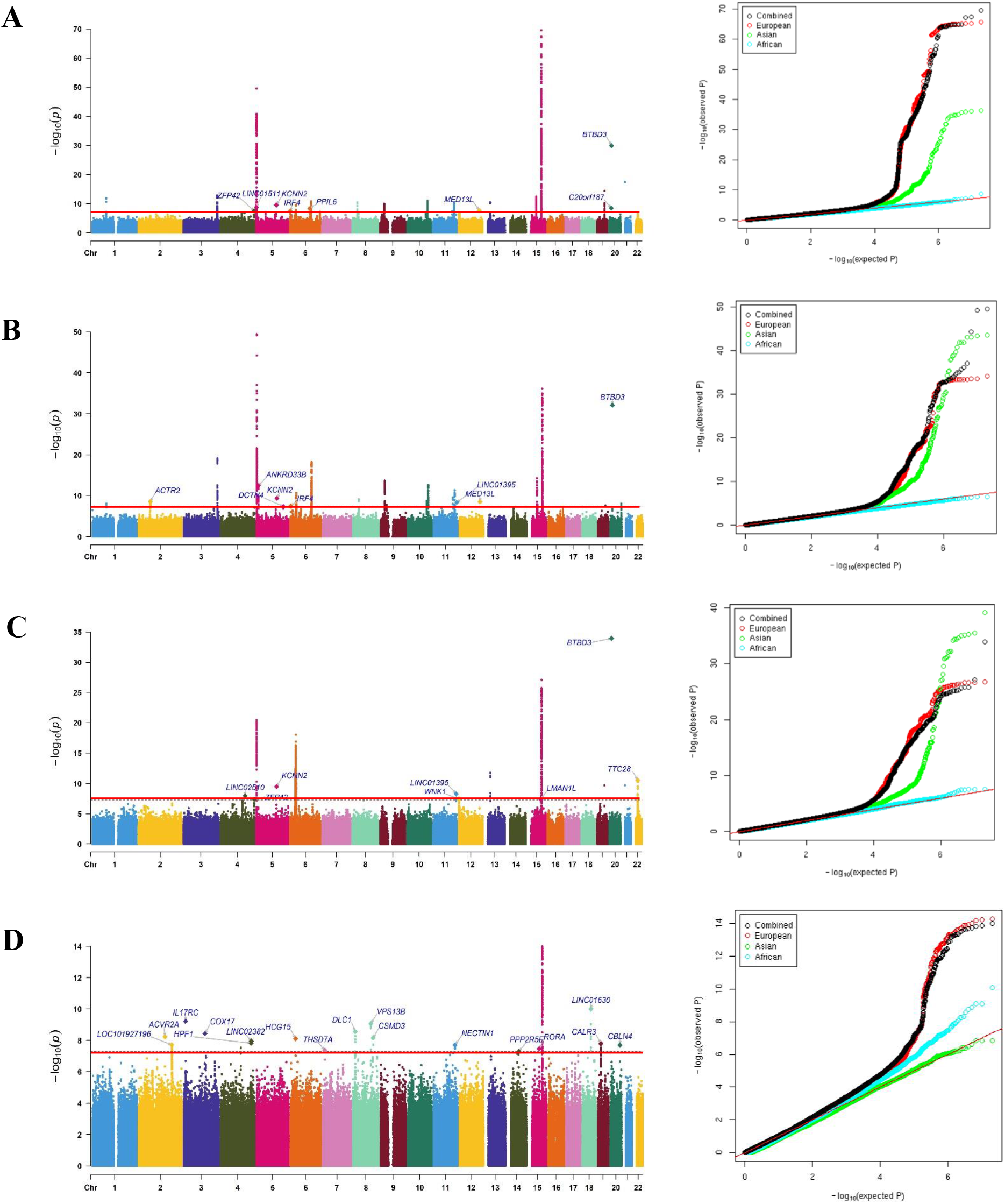
Manhattan plots and quantile-quantile plots of the GWAS meta-analysis for lung cancer in the cross-ancestry analyses. (A) Lung carcinoma: 35,732 cases and 34,424 controls. (B) Adenocarcinoma: 14,716 cases and 29,648 controls. (C) Squamous cell carcinoma: 7,628 cases and 29,648 controls. (D) Small cell carcinoma: 2,229 cases and 21,612 controls. The x-axis represents chromosomal location, and the y-axis -log10(P-value). The newly identified genes are represented in blue. The red horizontal line denotes −*log*_10_*P* = − *log*_10_ 5 × 10^−8^.

### Genome-wide discovery in multiple ancestries

The multi-ethnic combined meta-analysis across three intercontinental populations of lung cancer identified 990, 991, 1234, and 173 SNPs for lung cancer, ADE, SQC, and SCC at genome-wide statistical significance levels (5×10^−8^), respectively. Ethnic-specific GWAS meta-analysis results of diverse ancestries showed that 45 cross-ancestry variants had I^2^ values^23^ less than 30%, indicating similarity of effect across populations (Supplementary Table 2). All genomic cross-ancestry loci contributing to lung cancer susceptibility with a P-value less than 1×10^−5^ are reported in Supplementary Table 3A-3D. As shown in Figure 1, we observed highly heterogeneous genetic architecture of lung cancer among selected well-known loci and additional candidates with 35 cross-ancestry variants having I^2^ values greater than 30% (Supplementary Table 2). We observed significant but heterogeneous association signals on 5p15.33 in lung cancer, and non-small cell lung cancer (NSCLC) including ADE, and SQC, but not in SCC (Supplementary Figure 1A and Supplementary Table 2 and 3A-3C). Similarly, the well known significant genetic signal on 15q25.1 was observed in lung cancer and all histological subtypes (Supplementary Figure 1B and Supplementary Table 2 and 3A-3D). Some genetic signals such as *BTBD3* commonly identified across ethnic groups were detected in lung cancer and NSCLC but not were found to influence SCC specifically (Supplementary Table 2 and 3A-3D). Many associations were found in SCC that were not identified for lung cancer and NSCLC (Supplementary Table 2 and 3A-3D).

**Table 2.**
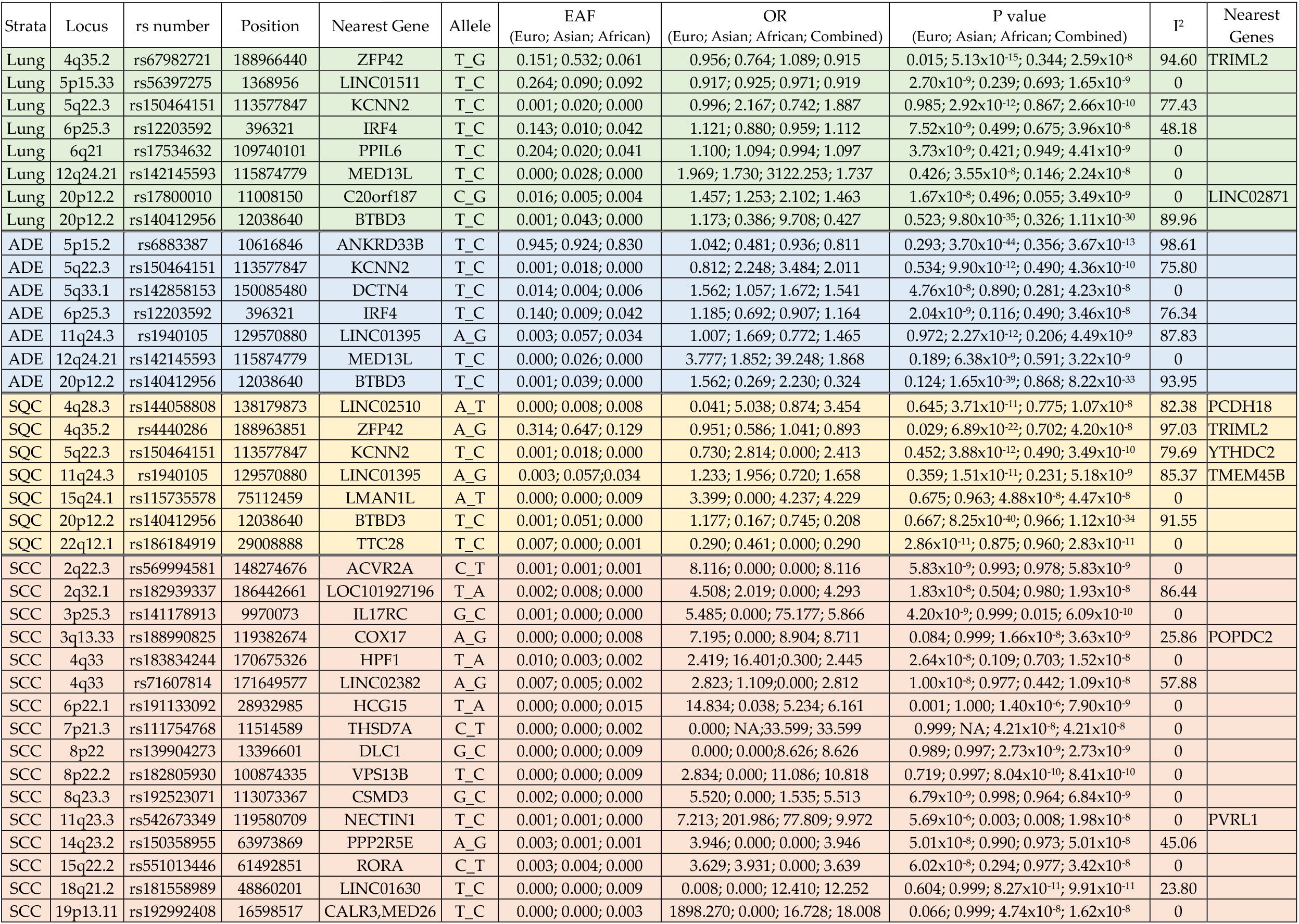
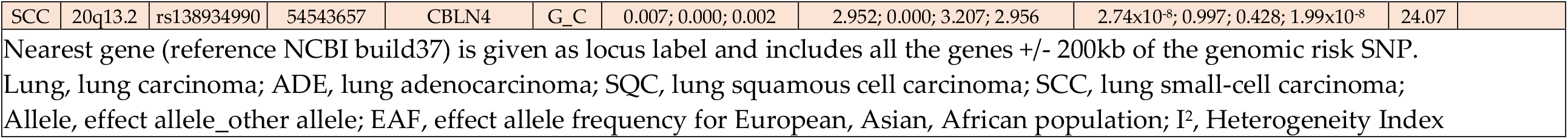
Novel genome-wide associations identified from the ethnic-specific and trans-ethnic meta-analysis in lung cancer and histological subtype analyses.

| Strata | Locus | rs number | Position | Nearest Gene | Allele | EAF<br>(Euro; Asian; African) | OR<br>(Euro; Asian; African; Combined) | P value<br>(Euro; Asian; African; Combined) | I<sup>2</sup> | Nearest Genes |
| --- | --- | --- | --- | --- | --- | --- | --- | --- | --- | --- |
| Lung | 4q35.2 | rs67982721 | 188966440 | ZFP42 | T_G | 0.151; 0.532; 0.061 | 0.956; 0.764; 1.089; 0.915 | 0.015; 5.13x10<sup>-15</sup>; 0.344; 2.59x10<sup>-8</sup> | 94.60 | TRIML2 |
| Lung | 5p15.33 | rs56397275 | 1368956 | LINC01511 | T_C | 0.264; 0.090; 0.092 | 0.917; 0.925; 0.971; 0.919 | 2.70x10<sup>-9</sup>; 0.239; 0.693; 1.65x10<sup>-9</sup> | 0 |  |
| Lung | 5q22.3 | rs150464151 | 113577847 | KCNN2 | T_C | 0.001; 0.020; 0.000 | 0.996; 2.167; 0.742; 1.887 | 0.985; 2.92x10<sup>-12</sup>; 0.867; 2.66x10<sup>-10</sup> | 77.43 |  |
| Lung | 6p25.3 | rs12203592 | 396321 | IRF4 | T_C | 0.143; 0.010; 0.042 | 1.121; 0.880; 0.959; 1.112 | 7.52x10<sup>-9</sup>; 0.499; 0.675; 3.96x10<sup>-8</sup> | 48.18 |  |
| Lung | 6q21 | rs17534632 | 109740101 | PPIL6 | T_C | 0.204; 0.020; 0.041 | 1.100; 1.094; 0.994; 1.097 | 3.73x10<sup>-9</sup>; 0.421; 0.949; 4.41x10<sup>-9</sup> | 0 |  |
| Lung | 12q24.21 | rs142145593 | 115874779 | MED13L | T_C | 0.000; 0.028; 0.000 | 1.969; 1.730; 3122.253; 1.737 | 0.426; 3.55x10<sup>-8</sup>; 0.146; 2.24x10<sup>-8</sup> | 0 |  |
| Lung | 20p12.2 | rs17800010 | 11008150 | C20orf187 | C_G | 0.016; 0.005; 0.004 | 1.457; 1.253; 2.102; 1.463 | 1.67x10<sup>-8</sup>; 0.496; 0.055; 3.49x10<sup>-9</sup> | 0 | LINC02871 |
| Lung | 20p12.2 | rs140412956 | 12038640 | BTBD3 | T_C | 0.001; 0.043; 0.000 | 1.173; 0.386; 9.708; 0.427 | 0.523; 9.80x10<sup>-35</sup>; 0.326; 1.11x10<sup>-30</sup> | 89.96 |  |
| ADE | 5p15.2 | rs6883387 | 10616846 | ANKRD33B | T_C | 0.945; 0.924; 0.830 | 1.042; 0.481; 0.936; 0.811 | 0.293; 3.70x10<sup>-44</sup>; 0.356; 3.67x10<sup>-13</sup> | 98.61 |  |
| ADE | 5q22.3 | rs150464151 | 113577847 | KCNN2 | T_C | 0.001; 0.018; 0.000 | 0.812; 2.248; 3.484; 2.011 | 0.534; 9.90x10<sup>-12</sup>; 0.490; 4.36x10<sup>-10</sup> | 75.80 |  |
| ADE | 5q33.1 | rs142858153 | 150085480 | DCTN4 | T_C | 0.014; 0.004; 0.006 | 1.562; 1.057; 1.672; 1.541 | 4.76x10<sup>-8</sup>; 0.890; 0.281; 4.23x10<sup>-8</sup> | 0 |  |
| ADE | 6p25.3 | rs12203592 | 396321 | IRF4 | T_C | 0.140; 0.009; 0.042 | 1.185; 0.692; 0.907; 1.164 | 2.04x10<sup>-9</sup>; 0.116; 0.490; 3.46x10<sup>-8</sup> | 76.34 |  |
| ADE | 11q24.3 | rs1940105 | 129570880 | LINC01395 | A_G | 0.003; 0.057; 0.034 | 1.007; 1.669; 0.772; 1.465 | 0.972; 2.27x10<sup>-12</sup>; 0.206; 4.49x10<sup>-9</sup> | 87.83 |  |
| ADE | 12q24.21 | rs142145593 | 115874779 | MED13L | T_C | 0.000; 0.026; 0.000 | 3.777; 1.852; 39.248; 1.868 | 0.189; 6.38x10<sup>-9</sup>; 0.591; 3.22x10<sup>-9</sup> | 0 |  |
| ADE | 20p12.2 | rs140412956 | 12038640 | BTBD3 | T_C | 0.001; 0.039; 0.000 | 1.562; 0.269; 2.230; 0.324 | 0.124; 1.65x10<sup>-39</sup>; 0.868; 8.22x10<sup>-33</sup> | 93.95 |  |
| SQC | 4q28.3 | rs144058808 | 138179873 | LINC02510 | A_T | 0.000; 0.008; 0.008 | 0.041; 5.038; 0.874; 3.454 | 0.645; 3.71x10<sup>-11</sup>; 0.775; 1.07x10<sup>-8</sup> | 82.38 | PCDH18 |
| SQC | 4q35.2 | rs4440286 | 188963851 | ZFP42 | A_G | 0.314; 0.647; 0.129 | 0.951; 0.586; 1.041; 0.893 | 0.029; 6.89x10<sup>-22</sup>; 0.702; 4.20x10<sup>-8</sup> | 97.03 | TRIML2 |
| SQC | 5q22.3 | rs150464151 | 113577847 | KCNN2 | T_C | 0.001; 0.018; 0.000 | 0.730; 2.814; 0.000; 2.413 | 0.452; 3.88x10<sup>-12</sup>; 0.490; 3.49x10<sup>-10</sup> | 79.69 | YTHDC2 |
| SQC | 11q24.3 | rs1940105 | 129570880 | LINC01395 | A_G | 0.003; 0.057; 0.034 | 1.233; 1.956; 0.720; 1.658 | 0.359; 1.51x10<sup>-11</sup>; 0.231; 5.18x10<sup>-9</sup> | 85.37 | TMEM45B |
| SQC | 15q24.1 | rs115735578 | 75112459 | LMAN1L | A_T | 0.000; 0.000; 0.009 | 3.399; 0.000; 4.237; 4.229 | 0.675; 0.963; 4.88x10<sup>-8</sup>; 4.47x10<sup>-8</sup> | 0 |  |
| SQC | 20p12.2 | rs140412956 | 12038640 | BTBD3 | T_C | 0.001; 0.051; 0.000 | 1.177; 0.167; 0.745; 0.208 | 0.667; 8.25x10<sup>-40</sup>; 0.966; 1.12x10<sup>-34</sup> | 91.55 |  |
| SQC | 22q12.1 | rs186184919 | 29008888 | TTC28 | T_C | 0.007; 0.000; 0.001 | 0.290; 0.461; 0.000; 0.290 | 2.86x10<sup>-11</sup>; 0.875; 0.960; 2.83x10<sup>-11</sup> | 0 |  |
| SCC | 2q22.3 | rs569994581 | 148274676 | ACVR2A | C_T | 0.001; 0.001; 0.001 | 8.116; 0.000; 0.000; 8.116 | 5.83x10<sup>-9</sup>; 0.993; 0.978; 5.83x10<sup>-9</sup> | 0 |  |
| SCC | 2q32.1 | rs182939337 | 186442661 | LOC101927196 | T_A | 0.002; 0.008; 0.000 | 4.508; 2.019; 0.000; 4.293 | 1.83x10<sup>-8</sup>; 0.504; 0.980; 1.93x10<sup>-8</sup> | 86.44 |  |
| SCC | 3p25.3 | rs141178913 | 9970073 | IL17RC | G_C | 0.001; 0.000; 0.000 | 5.485; 0.000; 75.177; 5.866 | 4.20x10<sup>-9</sup>; 0.999; 0.015; 6.09x10<sup>-10</sup> | 0 |  |
| SCC | 3q13.33 | rs188990825 | 119382674 | COX17 | A_G | 0.000; 0.000; 0.008 | 7.195; 0.000; 8.904; 8.711 | 0.084; 0.999; 1.66x10<sup>-8</sup>; 3.63x10<sup>-9</sup> | 25.86 | POPDC2 |
| SCC | 4q33 | rs183834244 | 170675326 | HPF1 | T_A | 0.010; 0.003; 0.002 | 2.419; 16.401; 0.300; 2.445 | 2.64x10<sup>-8</sup>; 0.109; 0.703; 1.52x10<sup>-8</sup> | 0 |  |
| SCC | 4q33 | rs71607814 | 171649577 | LINC02382 | A_G | 0.007; 0.005; 0.002 | 2.823; 1.109; 0.000; 2.812 | 1.00x10<sup>-8</sup>; 0.977; 0.442; 1.09x10<sup>-8</sup> | 57.88 |  |
| SCC | 6p22.1 | rs191133092 | 28932985 | HCG15 | T_A | 0.000; 0.000; 0.015 | 14.834; 0.038; 5.234; 6.161 | 0.001; 1.000; 1.40x10<sup>-6</sup>; 7.90x10<sup>-9</sup> | 0 |  |
| SCC | 7p21.3 | rs111754768 | 11514589 | THSD7A | C_T | 0.000; 0.000; 0.002 | 0.000; NA; 33.599; 33.599 | 0.999; NA; 4.21x10<sup>-8</sup>; 4.21x10<sup>-8</sup> | 0 |  |
| SCC | 8p22 | rs139904273 | 13396601 | DLC1 | G_C | 0.000; 0.000; 0.009 | 0.000; 0.000; 8.626; 8.626 | 0.989; 0.997; 2.73x10<sup>-9</sup>; 2.73x10<sup>-9</sup> | 0 |  |
| SCC | 8p22.2 | rs182805930 | 100874335 | VPS13B | T_C | 0.000; 0.000; 0.009 | 2.834; 0.000; 11.086; 10.818 | 0.719; 0.997; 8.04x10<sup>-10</sup>; 8.41x10<sup>-10</sup> | 0 |  |
| SCC | 8q23.3 | rs192523071 | 113073367 | CSMD3 | G_C | 0.002; 0.000; 0.000 | 5.520; 0.000; 1.535; 5.513 | 6.79x10<sup>-9</sup>; 0.998; 0.964; 6.84x10<sup>-9</sup> | 0 |  |
| SCC | 11q23.3 | rs542673349 | 119580709 | NECTIN1 | T_C | 0.001; 0.001; 0.000 | 7.213; 201.986; 77.809; 9.972 | 5.69x10<sup>-6</sup>; 0.003; 0.008; 1.98x10<sup>-8</sup> | 0 | PVRL1 |
| SCC | 14q23.2 | rs150358955 | 63973869 | PPP2R5E | A_G | 0.003; 0.001; 0.001 | 3.946; 0.000; 0.000; 3.946 | 5.01x10<sup>-8</sup>; 0.990; 0.973; 5.01x10<sup>-8</sup> | 45.06 |  |
| SCC | 15q22.2 | rs551013446 | 61492851 | RORA | C_T | 0.003; 0.004; 0.000 | 3.629; 3.931; 0.000; 3.639 | 6.02x10<sup>-8</sup>; 0.294; 0.977; 3.42x10<sup>-8</sup> | 0 |  |
| SCC | 18q21.2 | rs181558989 | 48860201 | LINC01630 | T_C | 0.000; 0.000; 0.009 | 0.008; 0.000; 12.410; 12.252 | 0.604; 0.999; 8.27x10<sup>-11</sup>; 9.91x10<sup>-11</sup> | 23.80 |  |
| SCC | 19p13.11 | rs192992408 | 16598517 | CALR3, MED26 | T_C | 0.000; 0.000; 0.003 | 1898.270; 0.000; 16.728; 18.008 | 0.066; 0.999; 4.74x10<sup>-8</sup>; 1.62x10<sup>-8</sup> | 0 |  |
| SCC | 20q13.2 | rs138934990 | 54543657 | CBLN4 | G_C | 0.007; 0.000; 0.002 | 2.952; 0.000; 3.207; 2.956 | 2.74x10<sup>-8</sup>; 0.997; 0.428; 1.99x10<sup>-8</sup> | 24.07 |  |
| Nearest gene (reference NCBI build37) is given as locus label and includes all the genes +/- 200kb of the genomic risk SNP.<br>Lung, lung carcinoma; ADE, lung adenocarcinoma; SQC, lung squamous cell carcinoma; SCC, lung small-cell carcinoma;<br>Allele, effect allele_other allele; EAF, effect allele frequency for European, Asian, African population; I<sup>2</sup>, Heterogeneity Index |  |  |  |  |  |  |  |  |  |  |

Ethnic-specific and the trans-ethnic meta-analysis regional association plots for novel cross-ancestry genetic variants are shown for each region in Supplementary Figure 2.

**Figure 2.**
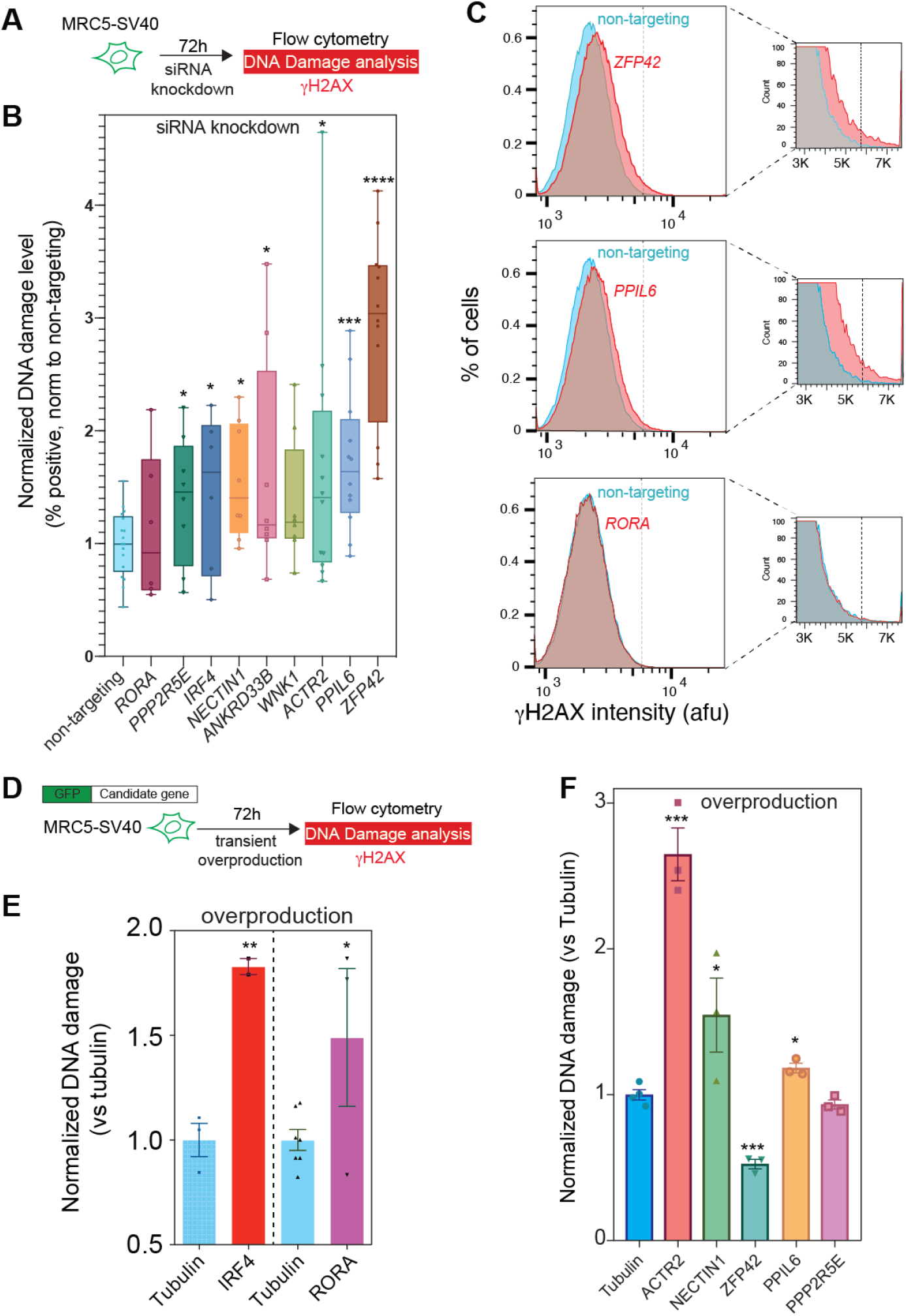
The functional validation of the DNA damageome genes and proteins from trans-ethnic lung cancer GWAS. (A) siRNA knockdown DNA damage assay scheme. (B) Multiple genes knockdown increase DNA damage (γH2AX) (mean± SEM, n>=5), including *ZFP42, PPIL6, ACTR2. γ*H2AX positive cells are quantified using a threshold described in methods. All candidates are normalized to non-targeting pooled siRNAs. (C) Representative flow cytometric histograms of (B) for ZFP42, PPIL6 (positive), and RORA (negative). Dash lines in the box: DNA damage threshold. (D) Overproduction DNA damage assay scheme. (E) Two transcription factors IRF4 and RORA overproduction increases γH2AX. N-GFP fused Tubulin as control. Normalized DNA damage summary: mean ± SEM, n=2∼7. (F) ACTR2, NECTIN1 are among the best DNA damage promotion protein tested. ZFP42 overproduction reduces the endogenous γH2AX level. Mean ± SEM, n>=3.

### Novel loci in trans-ethnic meta-analyses for lung carcinoma

Of 990 genome-wide significant risk SNPs located in 68 distinct genomic regions, 28 regions contained SNPs not previously identified at genome-wide significance (Supplementary Table 2). In Table 2 we present 6 novel lung cancer risk loci based on trans-ethnic meta-analysis results of all loci, with sentinel variants showing the lowest P-value at each locus, LD structure based on all populations of the 1000 Genome Project, and putative target genes identified using the functional mapping and annotation” (FUMA) platform (Online Method)^24^. An intergenic variant (rs67982721) associated with lung cancer development on 4q35.2 near *ZFP42* and *TRIML2* was detected. *ZFP42* has previously been included in methylation models that predict the occurrence of lung cancer^25,26^. *TRIML2* variants have been reported to be associated with metabolism of cigarette-associated xenobiotics and eye morphology^27,28^. At the locus 5p15.33 that contains the known lung adenocarcinoma-associated genes *TERT* and *CLPTM1L*^29-33^, an ncRNA intronic variant (rs56397275) in the long intergenic non-protein coding RNA *LINC01511* was associated with lung cancer risk independent of prior hits in the region. Other *LINC01511* variants have previously been associated with prostate cancer and lupus. We identified rs150464151 near *KCNN2* on 5q22.3, a region previously associated with educational attainment an psychiatric illnesses^34^. A novel intronic associations was detected at rs12203592 in *IRF4* (Interferon Regulatory Factor 4) on 6p25.3, a variant previously associated with numerous pigmentation traits^35^. The IRF family member is lymphocyte specific and negatively regulates Toll-like-receptor (TLR) signaling, which is central to the activation of innate and adaptive immune systems. The novel association at rs17534632 in *PPIL6* (Peptidylprolyl Isomerase Like 6) on 6q21 is also associated with eosinophil count in European populations^36^. An association with intergenic variant rs142145593 on 12q24.21 was observed near *MED13L* (Mediator Complex Subunit 13L) whose encoded protein is involved in early development of the heart and brain, and *TBX3* (T-Box Transcription Factor 3) involved in the regulation of developmental processes and reported in colorectal cancer development and prostatic hyperlasia^37,38^. Novel variants, rs17800010 of *C20orf187* (*LINC02871*) and rs140412956 of near *BTBD3*, on 20p12.2 were identified. *LINC02871* was previously associated with bladder carcinoma, adolescent idiopathic scoliosis, and white blood cell count^36,39,40^. *BTBD3* (BTB Domain Containing 3) has been associated with lung function, hepatocellular carcinoma, smoking behaviors, and leukemia^41-44^. *BTBD3* was also reported in the association with response to first-line carboplatin or cisplatin plus etoposide chemotherapy in patients with SCC^45^.

### Novel loci in trans-ethnic meta-analyses for small cell lung carcinoma

We further investigated the genetic association with specific lung cancer histological subtypes and identified additional 22 novel loci achieving genome-wide significance (Table 2 and Supplementary Table 2). For SCC, we detected 17 novel rare variants in 16 genomic loci. At 2q22.3, an intergenic variant rs569994581 near *ACVR2A*-*PABPC1P2* was associated with SCC development and showed a strong association signal driven by the European-ancestry subset. We identified another intergenic variant rs182939337 near *LOC101927196*-*LOC105373782* on 2q32.1. At 3p25.3, an exonic variant rs141178913 of *IL17RC* (interleukin 17 receptor C), involved in the progression of inflammatory and autoimmune diseases as part of the IL-17 pathway, was found in association with SCC susceptibility. We found an intergenic variant near *COX17*-*POPDC2*, rs188990825 on 3q13.33. A protein coding gene, *COX17* (Cytochrome C Oxidase copper chaperone 17) has been previously associated with Menkes disease and metal metabolism disorder, and implicated as a therapeutic target of NSCLC^46^. Two variants, rs183834244 (*HPF1*) and rs71607814 (near *LINC02382* and *LINC01612*) on 4q33 were associated with SCC risk. Histone parylation factor 1 (*HPF1*) is a protein coding gene and a cofactor interacting with *PARP1* and *PARP2*, which are primarily involved in DNA repair and other cell functions^47^. A novel association at rs191133092 near HLA complex group 15 (*HCG15*) was detected, which is an RNA gene associated with unipolar depression^48^. We observed a novel association in Thrombospondin type 1 domain containing 7A (*THSD7A*) at rs111754768 on 7p21.3 that demonstrated a strong genetic signal in African-ancestry subjects. *THSD7A* is known for coronary artery disease susceptibility^49^. We found novel SNPs with strong evidence of associations with SCC at rs139904273 in *DLC1* on 8p22 and at rs182805930 in *VPS13B* on 8q22.2 presenting significant genetic signal in African-ancestry populations, and intergenic variant rs192523071 near *CSMD3* on 8q23.3 with significant signal in European-ancestry populations. *DLC1* encodes a tumor suppressor gene involved in a number of common cancers, including prostate, lung, colorectal, and breast cancers and reported in many GWAS of BMI^50^, lung function^36^, emphysema^2^, and SCC – but not previously achieving genome-wide significance^14^. An intronic variant rs542673349 of *NECTIN1* (*PVRL1*) on 11q23.3 showed genomic signals across all diverse populations. Nectin cell adhesion molecule 1 (*NECTIN1*) belongs to the immunoglobulin superfamily and is involved in susceptibility to bacterial meningitis measurement ^52^ and Tourette syndrome^53^. We also identified a novel association at rs150358955 in *PPP2R5E* on 14q23.2 showing strong association in European populations. Another variant (rs551013446) in the protein-coding gene RAR related orphan receptor A (*RORA*) on 15q22.22 demonstrated a strong genetic signal in Europeans and has emerged in prior GWAS of asthma^54,55^, pulmonary function^56^, metabolites level study^57^, atopic disease (asthma, hay fever, or eczema)^58^, multiple sclerosis^59^, allergic rhinitis^60^, BMI^2^, and schizophrenia^62^. We identified 2 novel loci, rs181558989 near *LINC01630*-*MEX3C* on 18q21.2 and rs192992408 in *CALR3* on 19p13.11 showing strongest evidence of genetic influence in African populations. A novel rare intergenic variant, rs138934990 in *CBLN4*-*LINC01441* on 20q13.2, was significantly associated across ethnicities, with the strongest association in Europeans.

### Novel loci in trans-ethnic meta-analyses for non-small cell lung carcinoma

At 4q28.3, intergenic variant rs144058808 between *LINC02510* and *PCDH18* was associated with squamous cell carcinoma risk (Table 2, Supplementary Table 2, and Supplementary Figure 2). Protocadherin 18, *PCDH18* has been reported in GWAS of blood pressure^63^, cardiovascular disease^36^, and use of beta-blocking agents^64^. We found an intergenic variant rs4440286 associated with squamous cell carcinoma development on 4q35.2 near *ZFP42* and *TRIML2* also identified the trans-ethnic meta-association with lung cancer.

*ZFP42* was reported as a SQC oncogene in the RNA-Seq study^25^. There were 3 novel associations on chromosome 5; rs6883387 in *ANKRD33B* on 5p15.2 with NSCLC risk, rs142858153 of near *DCTN4*-*RBM22* on 5q33.1 with ADE risk, and rs150464151 of near *KCNN2*-*YTHDC2* on 5q22.3 with all lung cancer including NSCLC. *ANKRD33B*, ankyrin repeat domain 33B has been previously associated with eosinophil counts^36^. Potassium calcium-activated channel subfamily N member 2, *KCNN2* was reported in a multi-ethnic GWAS of atrial fibrillation^65^ and coronary aneurysm^66^. Dynactin subunit 4, *DCTN4* demonstrated an association with traits for lung adenocarcinoma^14^, cholangiocarcinoma^67^, and lung function^2^ but had not reached genome-wide significance in the previous study of lung cancer. An intronic variant rs12203592 of *IRF4* on 6p25.3 was also associated with ADE as well as lung cancer.

Novel variant rs1940105 near *LINC01395*-*TMEM45B* showed strong genetic signal in Asians and was found to be associated with ADE and SQC development. There was a low-frequency variant rs142145593 in an intergenic region *MED13L*-*TBX3* on 12q24.21 with large ORs for ADE and lung cancer. This variant was the most frequent in Asian population (2.6%). A rare variant, rs115735578 in *LAMN1L* on 15q24.1 showed strong signals in African-ancestry populations and appeared to have a genetic influence on SQC development. A sentinel association was found on 20p12.2 at rs140412956 near *BTBD3* with lung cancer, ADE, and SQC risk and demonstrating the strongest association in Asian-ancestry populations. At 22q12.1, a rare *TTC28* variant, rs186184919 was associated with SQC risk.

*TTC28* has previously shown the association with breast cancer^68^, white blood cell count^69^, BMI^50^, prostate carcinoma in African-ancestry subjects^70^, and serous invasive ovarian cancer^2^.

### New signals of association in the previously reported loci

We performed conditional trans-ethnic meta-analysis, including dosages of the lead variant and principal components as covariates. Along with searching the top signals in lung multi-ethnic case-control meta-analyses, we further investigated secondary association signals at 5p15.33, conditioning on rs2853677, to test whether there are any other independent SNP association in the region^72^. We found two additional independent variants, *TERT*^14,33,73-75^ rs13167280 and *CLPTM1L*^14,29,32,76^ rs31487 on 5p15.33 were associated with lung cancer risk (Supplementary Figure 3 and Supplementary Table 4). GWAS have identified several lung cancer risk loci on the human leukocyte antigen (HLA) region on chromosome 6. Two variants at 6p21.32, rs11757382 near *HLA-DRB1* and *HLA-DQA1*^14,77^ for lung cancer and rs3129860 near *HLA-DRA*^14^ and *LOC101929163* for ADE were observed. The genetic locus at 6p21.33 (rs9267123 near *HCP5* and *LINC01149*) was shown to be associated with SQC having consistent genetic signals across diverse populations^14^. HLA complex P5, *HCP5* has been reported in many traits and diseases including lymphocyte count^69^, myositis^78^, psoriasis^79^ and HIV-1 infection^80^. A protein coding intergenic variant, rs9374662 near *DCBLD1*-*GOPC* was associated with lung cancer and ADE development^14,73,81^. We identified an intronic variant rs72477506 of *EPHX2* (*CHRNA2*)^14^ on 8p21.2 associated with lung cancer and showing a strong protective effect in European-ancestry populations compared to other populations. Epoxide hydrolase 2, *EPHX2* and cholinergic receptor nicotinic alpha 2 subunit, *CHRNA2* are also associated with smoking status measurement^43^. Two intergenic variants, rs1414259 and rs7850447 near *MTAP*-*MIR31HG* on 9p21.3 associated with lung cancer and ADE, respectively. Both SNPs on 9p21.3 showed strong genetic signals in European-ancestry population and moderate association in Asian-ancestry populations. Methylthioadenosine phosphorylase, *MTAP* is associated with melanoma^82-84^, lung carcinoma^14^, and lung function^85^. *MTAP* encodes an enzyme with a major role in polyamine metabolism and the encoded enzyme is deficient in many cancers due to co-deletion of *MTAP* and tumor suppressor p16 gene. At 10q25.2, rs41292624 in *VTI1A* demonstrated association in East Asians^73,81^. A new intronic variant rs11607355 of *JAML*^14^ (*AMICA1*) on 11q23.3 was associated with lung cancer susceptibility. We found a genetic locus at 15q21.1, near *COPS2*-*SECISBP2L*^14,73^, where rs2413932 was associated with lung cancer in Europeans, with similar OR in Asians and slightly weaker effect in African-Americans (Supplementary table 3A). The genetic variant rs268853 at 2p14 was associated with ADE and is located in *ACTR2* with a consistent direction of genetic effect across all populations (Table 2, Supplementary Table 2, and Supplementary Figure 2). *ACTR2*, actin-related protein 2 is a protein-coding gene and recently reported in ADE^14,73^. The 9p13.3 locus has been suggested as ADE susceptibility locus^14,73^ and a new sentinel variant rs4879704 of near *AQP3*-*AQP7* was identified for ADE. We found a new intergenic variant rs7902587 near *STN1*-*SLK*^14^ on 10q24.33 associated with ADE. The 11q22.3 locus, recently identified in Ashkenazi Jewish population^19^, harbors a rare exonic variant rs56009889 of *ATM* associated with ADE risk that was confirmed in our trans-ethnic combined studies, showing the genetic signal in European-ancestry populations. We found another new SQC association at rs12305739 of *WNK1* on 12p13.33. *WNK1* has been associated with lung carcinoma, SQC, lung cancer in ever smokers^14^, blood protein levels^86^, colorectal cancer in East Asians^87^, BMI^36^ and eosinophil count^69^. A ncRNA intronic variant near *RTEL1* rs75031349 on 20q13.33, showed strong evidence for ADE risk and was confirmed at genome-wide significance. Regulator of telomere elongation helicase 1, *RTEL1* that is an ATP-dependent DNA helicase identified initially in mice as a dominant telomere length regulator is linked to lung cancer development in a Chinese Han population^88^.

### Functional annotation of candidate causal alleles

We prioritized 61 unique genes from 22 of lung cancer susceptibility loci, of which 31 genes were identified by positional mapping of deleterious coding SNPs (CADD ≥ 12.37) and 29 genes were detected by gene expression quantitative trait loci (eQTL) associated with the expression of lung tissue samples using FUMA platform^24,89^ (Supplementary Table 5, Supplementary Table 6A-6D). Detailed characterization of causal genomic variants is described in Online Methods. Among lung histological subtypes, 71, 245, and 43 unique genes were prioritized for ADE, SQC, and SCC, respectively. Of these, 28, 78, and 14 deleterious coding SNPs were discovered and 32, 48, and 3 genes were associated with mRNA expression from lung tissue using GTEx (v8/Lung) (Supplementary Table 5). We further performed gene expression quantitative trait locus (eQTL) analysis using GTEx (v8/Lung) to link SNPs with the genes they regulate. The genetic variant rs12203592 in the IRF4 gene was significantly associated with gene expression in eQTL analysis from 515 noncancerous lung tissues with European descent (Supplementary Figure 4). As presented in multi-tissue eQTL comparison of Supplementary Figure 4, a single-tissue eQTL analysis with 515 lung samples indicated the strongest significant P-value (P = 8.1×10^−14^) and the largest posterior probability ranging between 0 and 1 that the lung tissue is predicted to have an eQTL effect (m-value = 1).

### Gene-based analysis

We performed gene-based analysis to complement the single-variant analysis in GWAS. Since the conventional single-variant analysis for common variants is often underpowered for SNPs with small effect sizes or rare alleles, it is challenging to elucidate the substantial contributions to missing heritability that are provided by such SNPs. Here we applied an aggregated Cauchy association test (ACAT) combining different single variant-level P-values to a group^90,91^. ACAT provides a guideline regarding the accuracy of the ACAT P-value^2^. When the ACAT P-value is very small (P < 10^−5^), the type I error is generally well-controlled under arbitrary correlation structures. When the ACAT P-value is moderately small (10^−3^ < P <10^−5^), the P-value is still generally accurate but may show slight inflation. In case of large ACAT P-values (P > 10^−3^), there can be potential type I error inflation due to moderately strong correlations among summary statistics. As presented in Supplementary Table 7, *IRF4* showed strong association with lung cancer and ADE (P = 1.16×10^−5^ and P = 1.15×10^−6^), respectively.

*TTC28*, tetratricopeptide repeat domain 28 is located near *CHEK2*, checkpoint kinase2 gene described as a tumor suppressor with proapoptotic, cell-cycle checkpoint and mitotic functions and associated with lung cancer^92^ and demonstrated that *TTC28* were also found to be strongly associated with SQC (P = 1.84×10^−7^). *IL17RC* encoding a single-pass type I membrane protein, interleukin-17 receptor C was identified as a strong susceptibility locus for SCC (P = 8.41×10^−7^).

### Integrative multi-omic annotation analysis

We integrated a variety of variant functional annotations to prioritize and characterize risk SNPs identified in cross-ancestry loci^93^ (Supplementary Table 8). The Multi-dimensional Annotation Class Integrative Estimator (MACIE) is a generalized linear mixed model designed to predict regulatory and evolutionarily conserved functional SNPs using 36 genome-wide annotations. Specially, the model treats functionality as an unobserved latent class and predicts (1) the probability of regulatory class only (MACIE01), (2) the probability of evolutionarily conserved class only (MACIE10), (3) the probability of neither class (MACIE00), or (4) the probability of both functional classes (MACIE11). In Table 3, MACIE does not predict any of top SNPs in cross-ancestry loci to be functional. However, these SNPs may possess non-regulatory or non-evolutionarily conserved function. IRF4 variant rs2316515 for ADE (OR=1.08, P=8.72×10^−7^) demonstrates a MACIE regulatory prediction of greater than 0.99 (Supplementary Table 9).

### Endogenous DNA damage in lung cancer

Endogenous DNA damage promotes mutations and cancer. The recent discovery of the DNA damageome proteins (DDPs) predicts that many GWAS/TWAS genes are driving cancer by DNA damage–promoting mechanisms ^12,94^. We hypothesized that a fraction of the GWAS-nominated cancer-associated genes promote cancer by increasing endogenous DNA damage and genome instability ^12^. A collection of 9 candidate-gene knockdown and 7 overproduction clones from trans-ethnic meta-summary statistics were screened for increased DNA damage, including *ZFP42* on 4q35.2, *PPIL6* on 6q21, *ACTR2* on 2p14, *IRF4* on 6p25.3, *RORA* on 15q22.2, *PPP2R5E* on 14q23.2, *NECTIN1* on 11q23.3, *ANKRD33B* on 5p15.2, *WNK1* on 12p13.33. We have screened all 16 candidates and confirmed the DNA-damage promoting activities for 7 knockdown genes and 5 overproduction clones (Figure 2). Surprisingly, *ZFP42* overproduction reduces the endogenous DNA damage, indicating a probable protection role. *ZFP42* is a well-known marker for pluripotency ^95^, although *ZFP42* is shown to involve in nucleotide excision repair ^96^ in algae, a similar role in human cells has not been discovered. The detailed mechanism underlying how *ZFP42* suppress DNA damage remains to be explored in future studies.

## DISCUSSION

We conducted trans-ethnic meta-analyses of lung cancer involving 51,961 European descendants, 12,434 Asian descendants, and 5,766 African-American descendants. While GWAS have identified a few dozen loci associated with lung cancer through the analysis based on homogeneous ethnic background, most of the findings are largely biased toward European-ancestry studies because trans-ethnic meta-analysis has not previously been feasible due to a lack of adequate data.

In this study, we identified 8, 7, 7, and 17 novel cross-ancestry SNPs associated with lung carcinoma, adenocarcinoma, squamous cell carcinoma, and small cell carcinoma, respectively. Lung carcinogenesis is a complex process involving the acquisition of genetic mutations and epigenetic changes that alter cellular processes, such as proliferation and differentiation^97^. It also seems to have distinct ethnic and geographical differences in lung cancer risk development. Many studies unveiled the pathogenesis of lung cancer^98^ but identifying novel genetic variants associated with lung cancer is still challenging due to small effect size and the strong contribution of tobacco smoking. To date, only a few lung cancer-specific genes have been detected. Mapping of lung cancer genome is an important process towards better understanding the pathogenesis of lung carcinoma. Therefore, further improved elucidation of cancer genetics in lung cancer health disparities is critical. For instance, better understanding the genomic diversity of oncogenes, tumor suppressor genes, or specific alterations across diverse ethnic populations can provide benefit fro designing population-specific targeted therapies. Also, deciphering the shared genetic variants underlying lung cancer predisposition in populations of diverse ancestry can help refine the risk prediction models for individuals at high-risk across ancestral populations. Our investigation of ancestry-specific and cross-ancestry association with lung cancer and specific histology subtypes resulted in several key findings.

First, we confirmed 9 genomic risk cross-ancestry loci for lung cancer and 12, 5, and 1 for ADE, SQC, and SCC, while concurrently identifying an additional 6 novel genome-wide significant risk loci for lung cancer and 7, 7, and 16 novel risk loci for ADE, SQC, and SCC, respectively.

We discovered ancestry-specific effects of common and rare coding variation on lung cancer among European, Asian, and African populations. A common variant, rs67982721in *ZFP42* showed very strong significant genetic signal among Asians and a weak one among Europeans and Africans. The common variant rs150464151 of *KCNN2* was associated with SQC only among Asians, with effect allele frequency (EAF) of 2% in Asians and 0.1% in other populations. Compared to sample sizes of Europeans and Asians, the number of African-American is still small. We highlighted the second insight of this study demonstrating a few significant genetic signals of association among African-American subjects. As presented in Supplementary Figure 2 including genomic regional association plots, a rare variant in all diverse populations, rs115735578 in *LMAN1L* uncovered the strong association with SQC risk only among African-Americans with EAF of 0.9%. Another population-specific association with SCC is a rare variation of rsrs139904273 in *DLC1*, with OR of 8.63 and P=2.73×10^−9^ in African-Americans with EAF of 0.9%.

A third insight of this study is the biological importance revealed through eQTL analyses. We investigated the functional consequence of several putatively causal alleles. Four genes, *IRF4, HLA-DQA1, JAML*, and *SECISBP2L* identified from lung cancer showed the strong significant association with gene expression in eQTL analysis. Two genes from ADE analysis, *IRF4* and *AQP3* confirmed the strong association with gene expression from 515 normal lung tissue from individuals of European descent.

Our final insight embraced the discovery of new DNA damageome genes and protein from trans-ethnic lung cancer GWAS. We provide the evidence that altered levels of most of these tested lung cancer-associated genes promote DNA damage. Altogether, our findings offer a hypothesis for how these genes are associated with lung cancer pathogenesis by promoting genome instability.

Population-specific GWAS with meta-analysis across these populations can help to elucidate the etiology and mechanisms of lung cancer and to identify more novel susceptibility biomarkers for better polygenic risk models of early detection and diagnosis, targeted therapy, and improved preventive measures.

## Data Availability

The data that support the findings of this study are available at the database of Genotypes and Phenotypes (dbGaP). The access numbers are phs001273.v3.p2 for OncoArray study, phs000876.v2.p1 for Affymetrix study, phs000716.v1.p1 for FLCCA study, phs001210.v1.p1 for NCI study, and phs000629.v1.p1 for GELCC study. Other data that support the findings of this study are available from the corresponding author upon request and are also reported as summary data in phs001273.v3.p2

## Acknowledgements

Our study was supported by the National Institutes of Health (NIH) for Integrative Analysis of Lung Cancer Etiology and Risk (U19CA203654) and Cancer Prevention Research Interest of Texas (CPRIT) award (RR170048). Functional Studies for this research was partially supported by NIH grants (R01CA250905 and DP1-AG072751). The Resource for the Study of Lung Cancer Epidemiology in North Trent (ReSoLuCENT) study was funded by the Sheffield Hospitals Charity, Sheffield Experimental Cancer Medicine Centre and Weston Park Hospital Cancer Charity. FT was supported by a clinical PhD fellowship funded by the Yorkshire Cancer Research/Cancer Research UK Sheffield Cancer Centre.

## Online Methods

### Multi-ethnic lung genome-wide association studies

There are 101,821 samples from 12 studies: Affymetrix Axiome Array Study (AFFY)^1^, the Female Lung Cancer Consortium in Asia (FLCCA)^2^, the Genetic Epidemiology of Lung Cancer (GELCC) Consortium, the Environment and Genetics in Lung cancer Etiology study (EAGLE)^3,4^, Helmholtz-Gemeinschaft Deutscher Forschungszentren Lung Cancer GWAS (GERMAN)^4,5^, the International Agency for Research on Cancer (IARC)^4^, the Institute of Cancer Research (ICR)^4^, MD Anderson Cancer Center Study (MDACC)^4,6^, NCI Lung Cancer and Smoking Phenotypes in African-American Cases and Controls (NCI)^7^, OncoArray Consortium Lung Study (OncoArray)^4,8^, the Prostate, Lung, Colorectal and Ovarian Cancer Screening Trial (PLCO)^4^ and Samuel Lunenfeld Research Institute Study (SLRI)^4^, after call rate of 0.95 was applied (Supplementary Note). Markers from various genotyping platforms were filtered based on the following criterion: only biallelic marker, call rate >=0.95 and no homogeneity. Markers were further checked using McCarthy Haplotype Reference Consortium (HRC) imputation preparation and checking tool (v4.2.11, https://www.well.ox.ac.uk/~wrayner/tools/) to make strand, position, ref/alt assignment consistent with HRC reference panel^9^. We conducted imputation of the phased data through Sanger imputation service in a two-stage strategy of pre-phasing and imputation using SHAPEIT2 (v2.r790) and PBWT (2014. The reference panel was HRC (r1.1), which contains 32,470 samples of predominantly European ancestry and about 40 million markers.

There were 2,854,462 common markers with information score of greater than or equal to 0.6 among 12 studies and were further thinned to 193,050 markers based on r-square value of less than or equal to 0.5. The new set of 193,050 markers was used to calculate principal components and pair-wise identity by descent (IBD) values among 101,821 samples in PLINK. An empirical value of IBD of 0.15 was used as a cutoff to define samples’ related status, and all related samples were categorized into 15,884 clusters. While priority of sample was quantified by scoring properties such as disease status and study specific measurement such as average imputation information score in each cluster and samples with missing disease status were assigned the lowest priority. Lists of independent or less-independent samples were generated and sorted by the total priority score. 70,639 samples with the highest scores in each cluster were finally generated for analysis through clustering and sampling process (Supplementary Note).

### Inference of ancestry memberships (Population stratification)

2,042 ancestry informative markers shared by 70,639 samples and 505 HapMap2 samples of CEU, CHB and YRI ancestry were used to infer ancestry origins using FastPop^10^, and then 51,961 samples of CEU origin, 12,434 samples of CHB origin and 5,766 samples of YRI origin were inferred (Supplementary Note). 14,716 ADE, 7,628 SQC, and 2,229 SCC were defined based on available histological information (Supplementary Note).

### Genome wide meta-analysis in across-ancestry loci

About 6 million markers of information score >=0.4 were analyzed using fixed logistical regression in PLINK (v1.9). The first 20 principal components and 12 study sites as factor variable were included in the model. Analyses were performed for histological in each diverse population such as European-, Asian-, and African-descent, respectively. Meta-analysis was further performed using PLINK to combine the fixed effects across diverse populations.

### Characterization of genomic risk loci using FUMA

We defined the regions of association by the most significant marker using functional mapping and annotation (FUMA) platform that computes LD structure, annotates functions to SNPs, and prioritize candidate genes from GWAS summary statistics^2^. A multi-ethnic meta-analysis across three intercontinental populations of lung cancer, totaling 35,732 cases and 34,424 controls, identified 990, 991, 1238, and 781 SNPs for lung carcinoma, ADE, SQC, and SCC at the genome significant level of 5×10^−8^, respectively.

For defining genomic risk loci for lung cancer susceptibility based on the combined GWAS summary statistics, linkage disequilibrium structure based on all populations of the 1000 Genome Project was used. Genomic risk loci and the subsets of significant SNPs within the loci were identified using the following criteria: (1.a) **independent significant SNPs**, defined as P < 5×10^−8^ and independent from each other at r^2^ < 0.6; (1.b) **candidate SNPs**, defined as those having r^2^ ≥ 0.6 with one of the independent significant SNPs and all of those candidate SNPs in the loci will be subject to further annotation; (2) **lead SNPs**, defined as independently significant SNPs and independent from each other at r^2^ < 0.1; (3) **genomic risk loci**, defined by merging lead SNPs within physically overlapped LD blocks and all SNPs in linkage disequilibrium of r^2^ ≥ 0.6 with one of the independent SNPs.

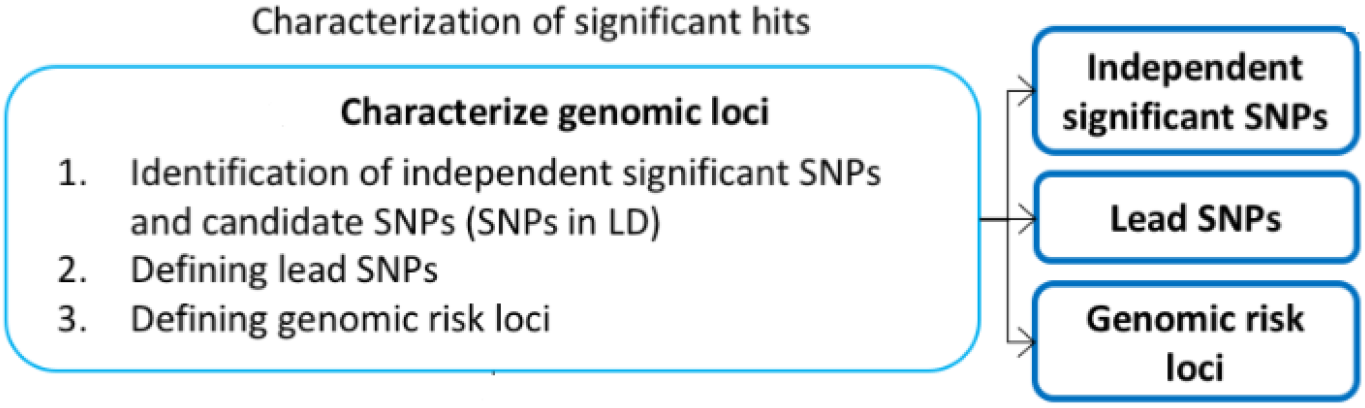

### Integrative multi-omic annotation analysis

We integrate a variety of variant functional annotations in a generalized linear mixed model (GLMM) approach to prioritize and characterize risk SNPs identified in cross-ancestry loci^12^. The Multi-dimensional Annotation Class Integrative Estimator (MACIE) models the regulatory and evolutionary conserved functionality of individual variants using two latent binary classes. Random effects are used to account for correlations among 8 annotations that are modeled as a function of the conserved class as well as 28 annotations that are modeled as a function of the regulatory class. Estimation occurs using an EM algorithm. The fitted model parameters are first found using a training dataset, and then one additional iteration of the EM algorithm is performed using these fitted parameters on the new SNPs of interest identified in this work. The MACIE output is a vector of 2*2 probabilities corresponding to the probabilities of belonging to both functional classes, either one of the classes alone, or neither class. The probabilities necessarily sum to 1. Marginal probabilities of regulatory function or evolutionarily conserved function can be found by simply adding two of the four probabilities. Formulating functionality as a set of multiple characteristics offers a more versatile and more detailed prediction than other integrative methods that produce a one-dimensional score that can be difficult to interpret.

