## Supplementary Figures for "Trans-ethnic genome-wide meta-analysis of 35,732 cases and 34,424 controls identifies novel genomic cross-ancestry loci contributing to lung cancer susceptibility"

### Supplementary Figure 1. Genomic regional plots on 5p15.33 and 15q25.1

#### A. TERT and CLPTM1L on 5p15.33

##### Lung Adenocarcinoma

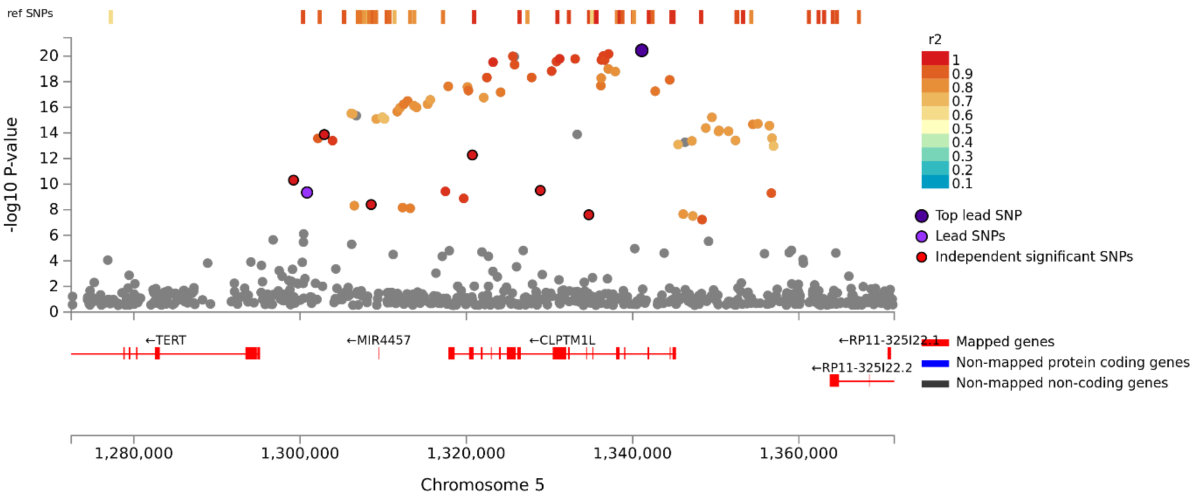


##### Lung Squamous Cell Carcinoma

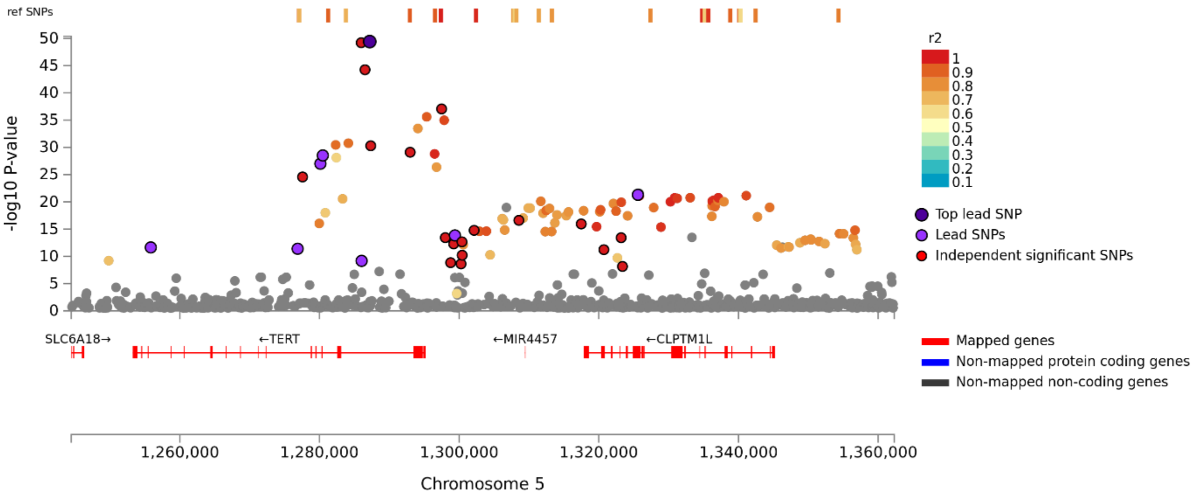

#### B. CHRNA5 on 15q25.1

##### Lung Adenocarcinoma

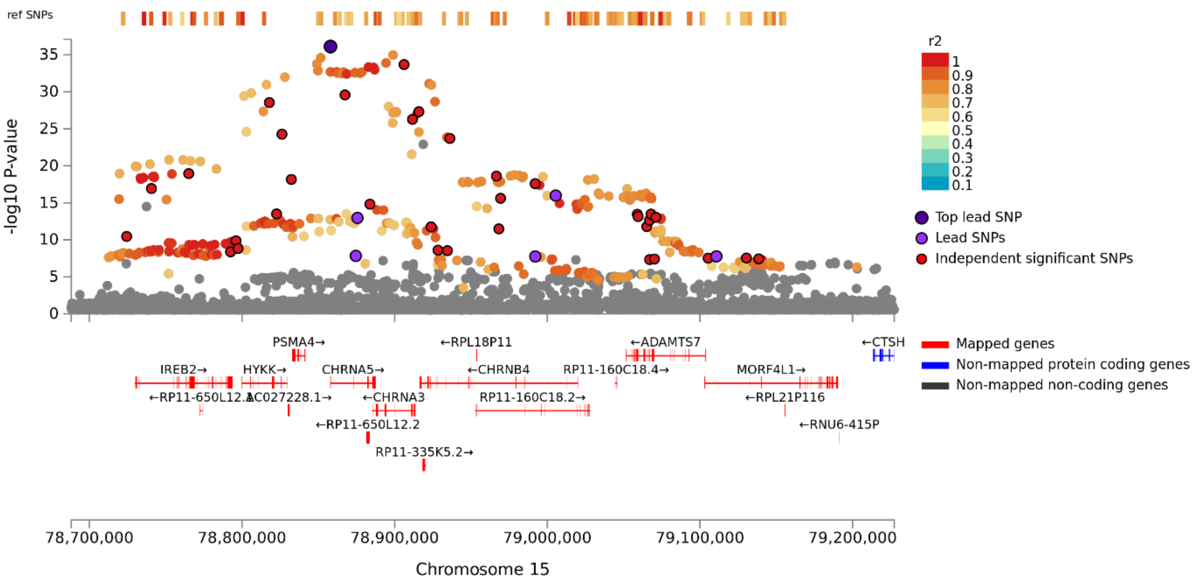


##### Lung Squamous Cell Carcinoma

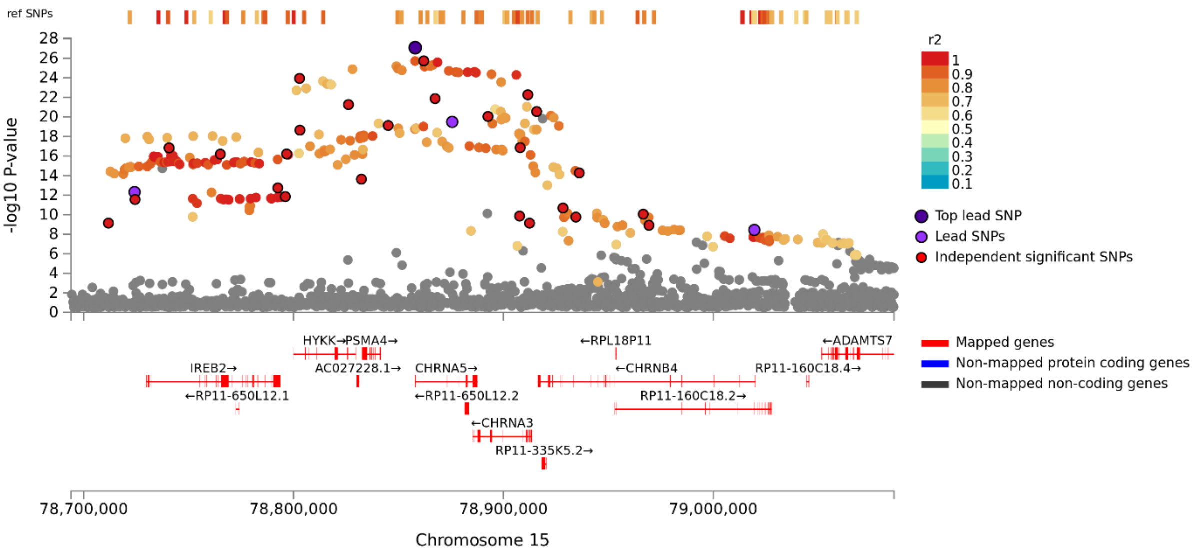

#### Lung Small Cell Carcinoma

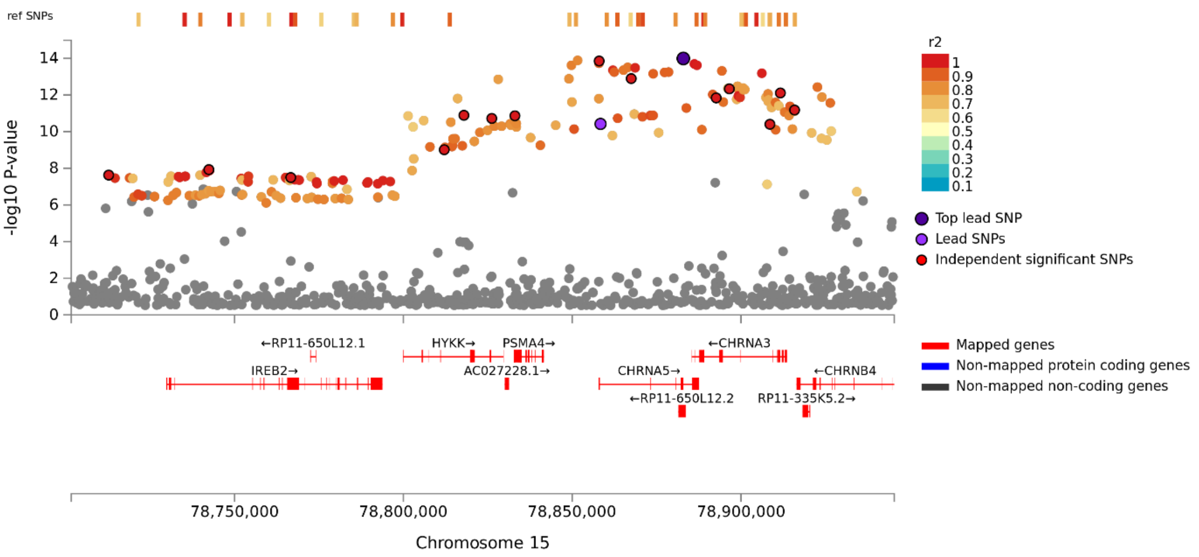

**Title:** Trans-ethnic genome-wide meta-analysis of 35,732 cases and 34,424 controls identifies novel genomic cross-ancestry loci contributing to lung cancer susceptibility

### Supplementary Figure 2
### Genomic Regional Plots

1. **Lung: Lung Carcinoma**
2. **LUAD: Lung Adenocarcinoma**
3. **LUSQC: Lung Squamous Cell Carcinoma**
4. **LUSCC: Lung Small Cell Carcinoma**

### Lung Carcinoma

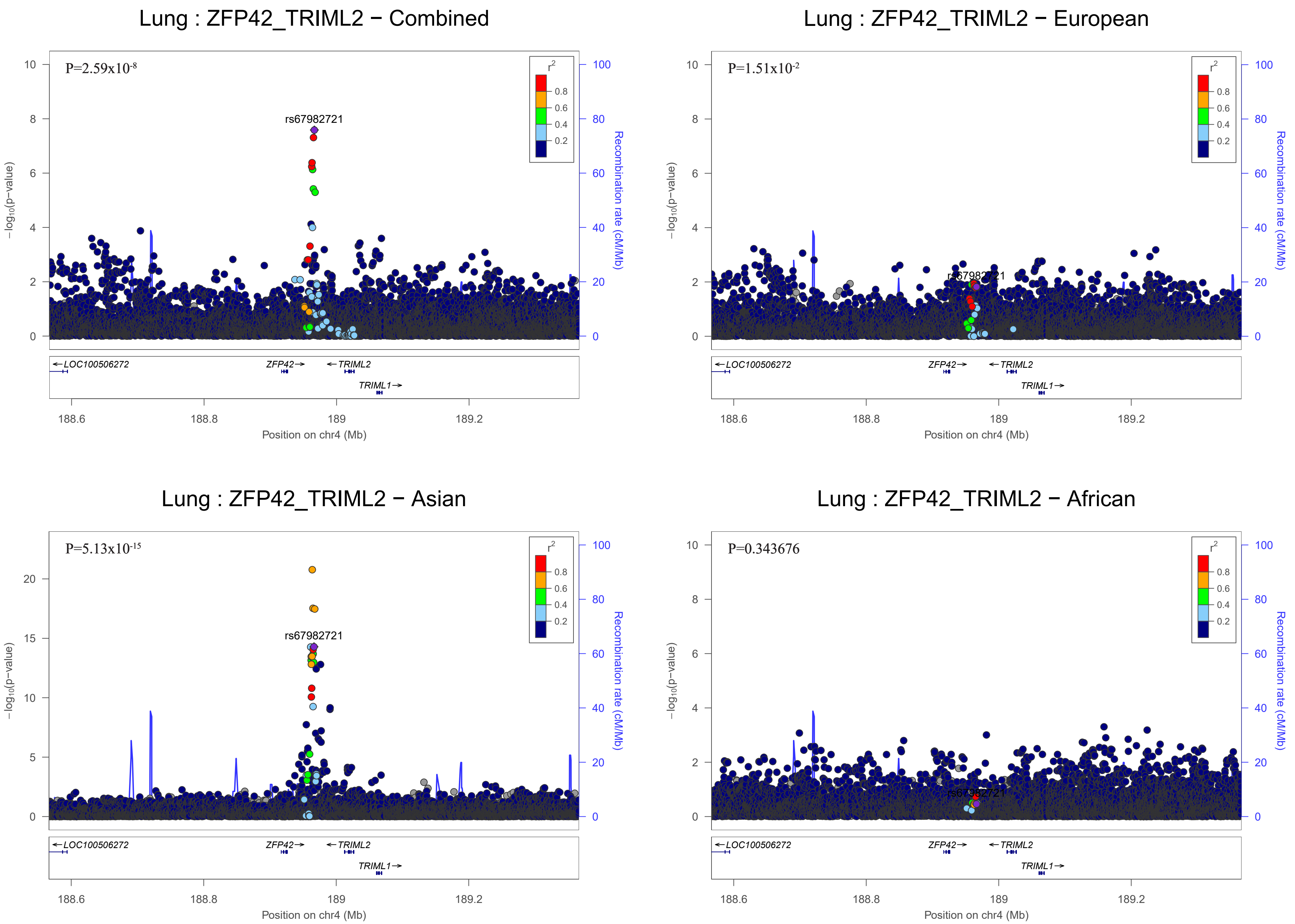

Lung : ZFP42_TRIML2 − Combined
P=2.59x10-8
rs67982721
Lung : ZFP42_TRIML2 − European
P=1.51x10-2
Lung : ZFP42_TRIML2 − Asian
P=5.13x10-15
rs67982721
Lung : ZFP42_TRIML2 − African
P=0.343676
r2
0.8
0.6
0.4
0.2
−log10(p−value)
Recombination rate (cM/Mb)
LOC100506272
ZFP42
TRIML2
TRIML1
188.6
188.8
189
189.2
Position on chr4 (Mb)

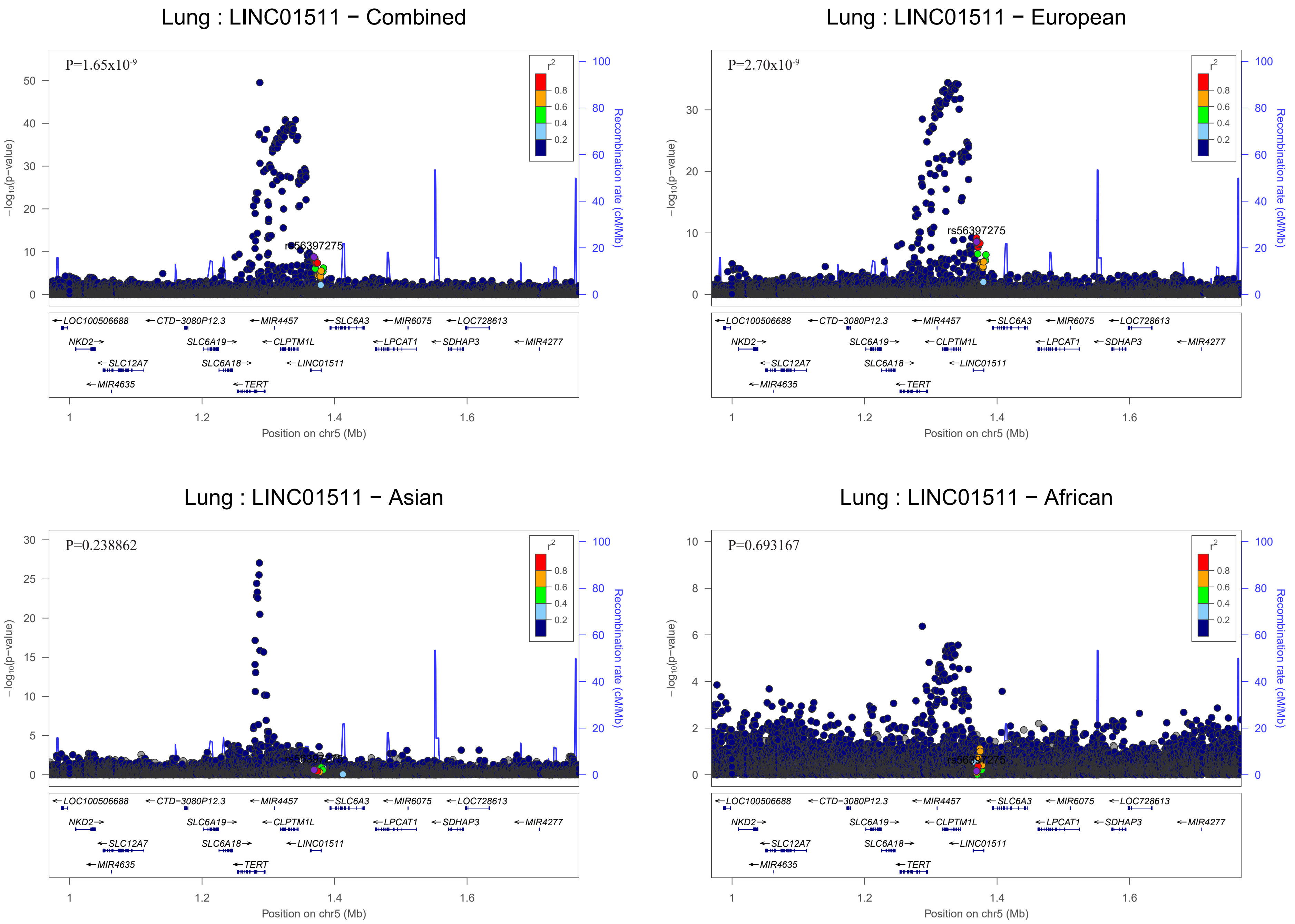
Lung : LINC01511 − Combined
P=1.65x10-9
rs56397275
Lung : LINC01511 − European
P=2.70x10-9
rs56397275
Lung : LINC01511 − Asian
P=0.238862
Lung : LINC01511 − African
P=0.693167
r2
0.8
0.6
0.4
0.2
−log10(p−value)
Recombination rate (cM/Mb)
Position on chr5 (Mb)
LOC100506688
CTD−3080P12.3
MIR4457
SLC6A3
MIR6075
LOC728613
NKD2
SLC6A19
CLPTM1L
LPCAT1
SDHAP3
MIR4277
SLC12A7
SLC6A18
LINC01511
MIR4635
TERT

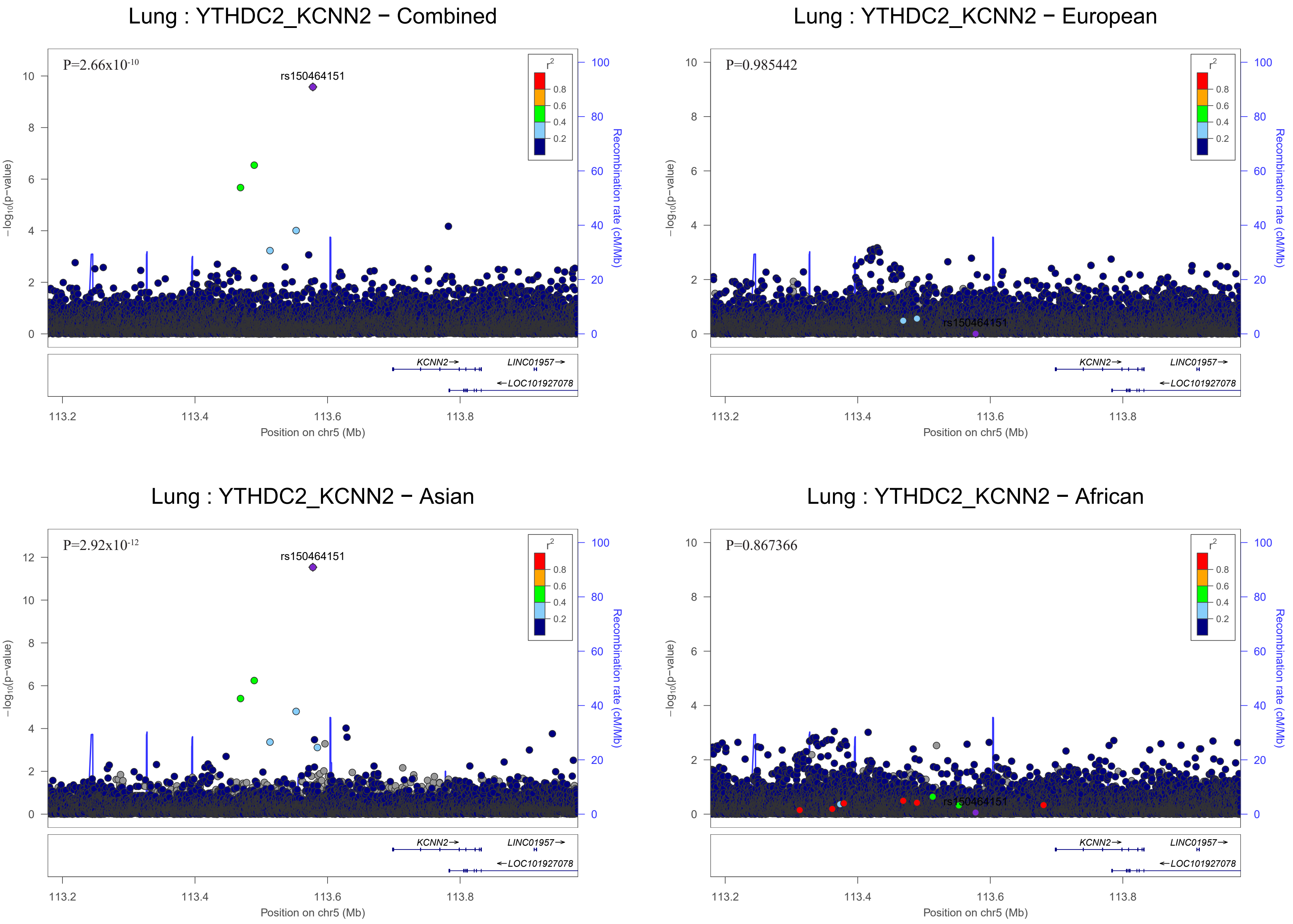
Lung : YTHDC2_KCNN2 − Combined
P=2.66x10-10
rs150464151
−log10(p−value)
Recombination rate (cM/Mb)
r2
0.8
0.6
0.4
0.2
KCNN2→
LINC01957→
←LOC101927078
113.2
113.4
113.6
113.8
Position on chr5 (Mb)
Lung : YTHDC2_KCNN2 − European
P=0.985442
rs150464151
−log10(p−value)
Recombination rate (cM/Mb)
KCNN2→
LINC01957→
←LOC101927078
Position on chr5 (Mb)
Lung : YTHDC2_KCNN2 − Asian
P=2.92x10-12
rs150464151
−log10(p−value)
Recombination rate (cM/Mb)
KCNN2→
LINC01957→
←LOC101927078
Position on chr5 (Mb)
Lung : YTHDC2_KCNN2 − African
P=0.867366
rs150464151
−log10(p−value)
Recombination rate (cM/Mb)
KCNN2→
LINC01957→
←LOC101927078
Position on chr5 (Mb)

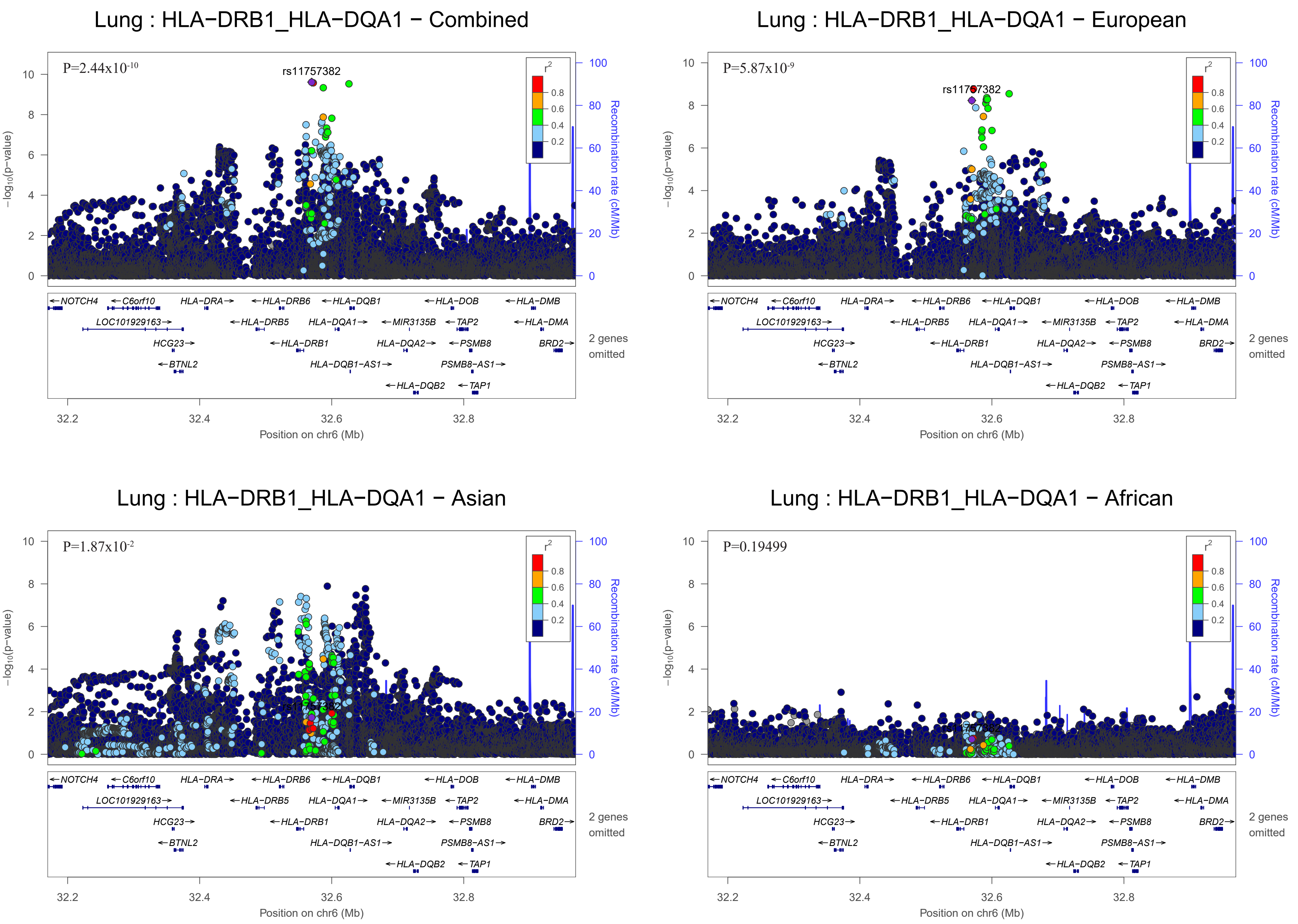
Lung : HLA−DRB1_HLA−DQA1 − Combined
P=2.44x10-10
rs11757382
Lung : HLA−DRB1_HLA−DQA1 − European
P=5.87x10-9
rs11757382
Lung : HLA−DRB1_HLA−DQA1 − Asian
P=1.87x10-2
Lung : HLA−DRB1_HLA−DQA1 − African
P=0.19499
r2
0.8
0.6
0.4
0.2
−log10(p−value)
Recombination rate (cM/Mb)
Position on chr6 (Mb)
NOTCH4
C6orf10
HLA−DRA
HLA−DRB6
HLA−DQB1
HLA−DOB
HLA−DMB
LOC101929163
HLA−DRB5
HLA−DQA1
MIR3135B
TAP2
HLA−DMA
HCG23
HLA−DRB1
HLA−DQA2
PSMB8
BRD2
BTNL2
HLA−DQB1−AS1
PSMB8−AS1
HLA−DQB2
TAP1
2 genes omitted

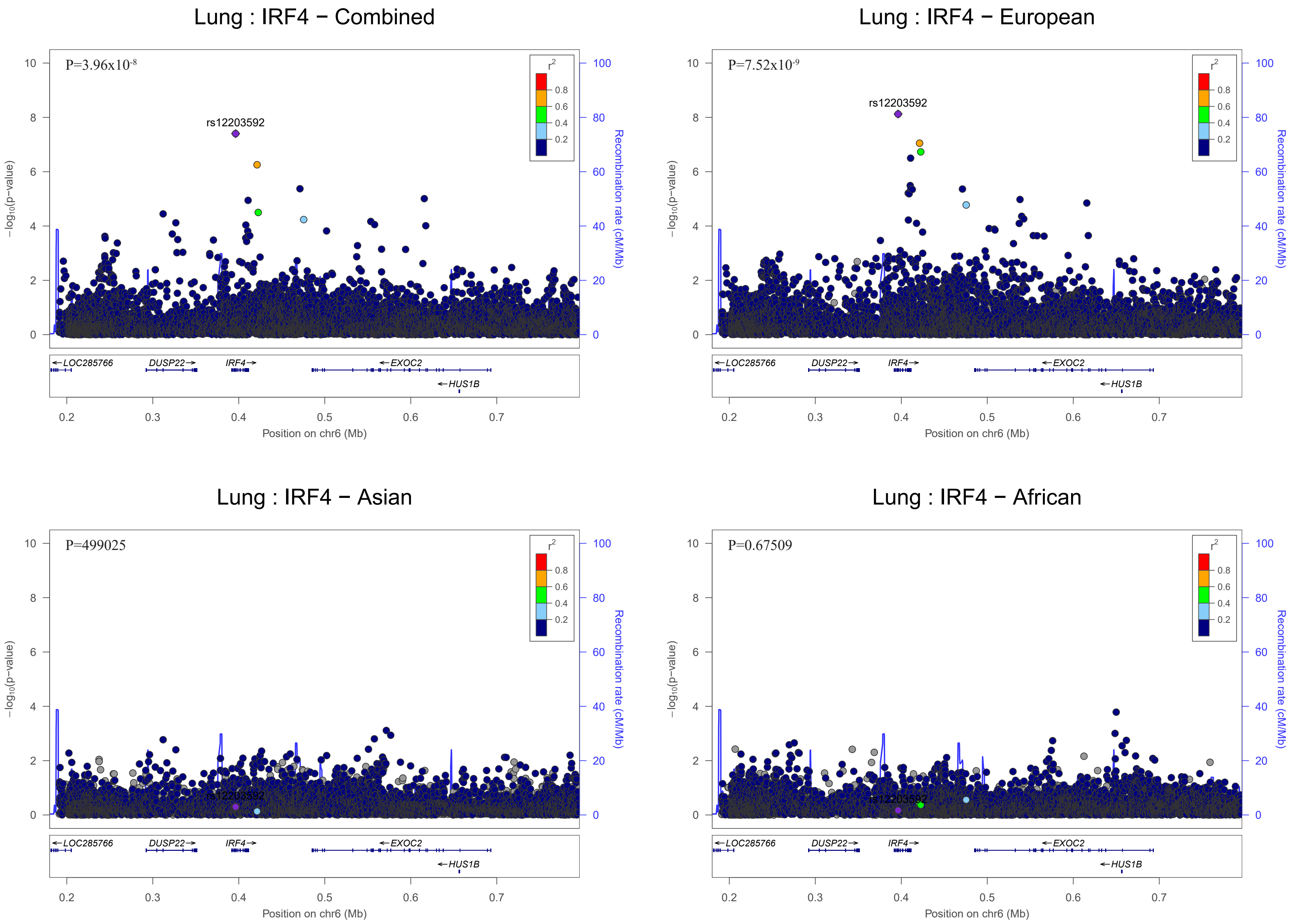

Lung : IRF4 − Combined
P=3.96x10-8
rs12203592
Lung : IRF4 − European
P=7.52x10-9
rs12203592
Lung : IRF4 − Asian
P=499025
rs12203592
Lung : IRF4 − African
P=0.67509
rs12203592
r2
0.8
0.6
0.4
0.2
−log10(p−value)
Recombination rate (cM/Mb)
LOC285766
DUSP22
IRF4
EXOC2
HUS1B
Position on chr6 (Mb)

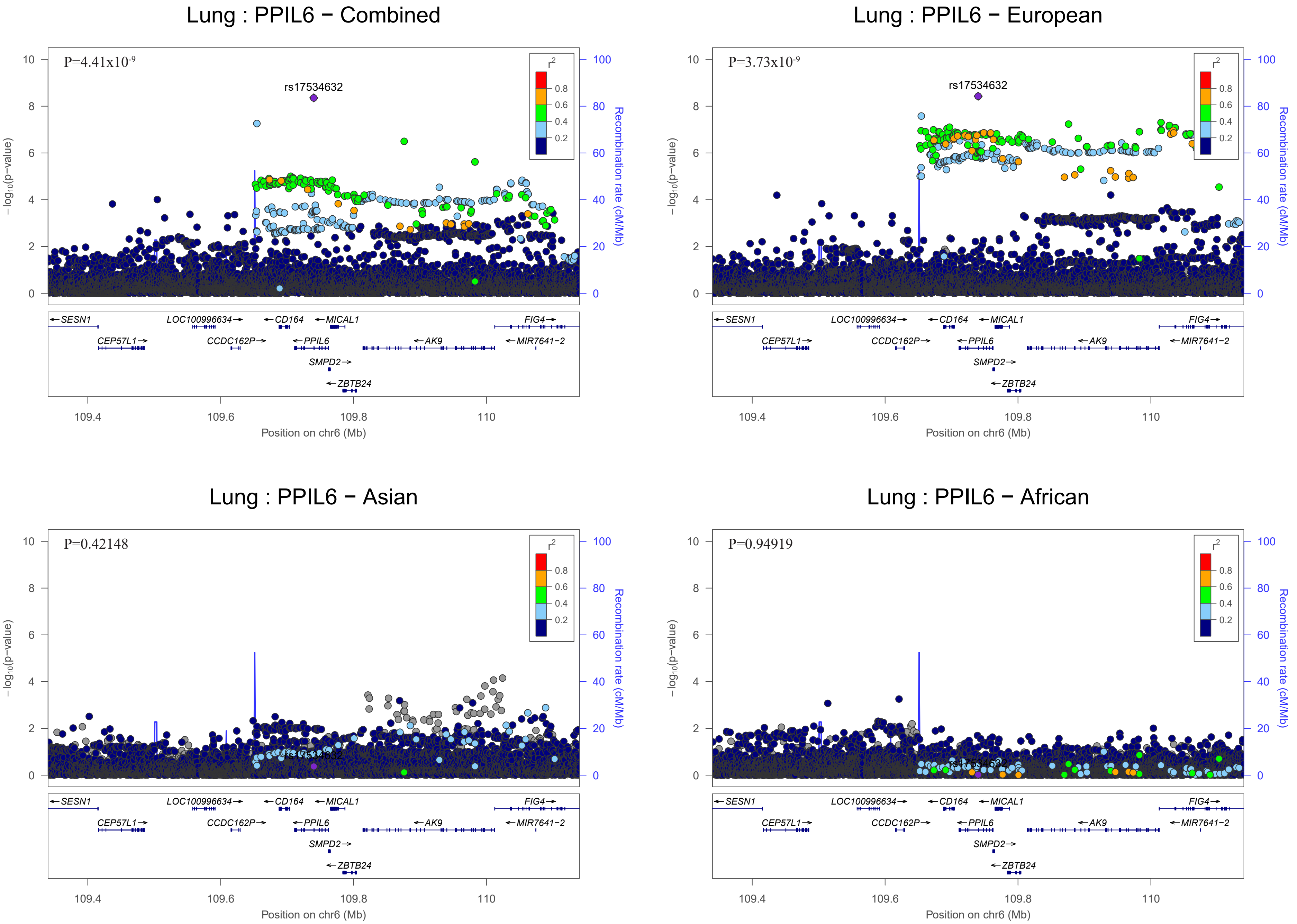
Lung : PPIL6 − Combined
P=4.41x10-9
rs17534632
Lung : PPIL6 − European
P=3.73x10-9
rs17534632
Lung : PPIL6 − Asian
P=0.42148
rs17534632
Lung : PPIL6 − African
P=0.94919
rs17534632
r2
0.8
0.6
0.4
0.2
−log10(p−value)
Recombination rate (cM/Mb)
SESN1
LOC100996634
CD164
MICAL1
FIG4
CEP57L1
CCDC162P
PPIL6
AK9
MIR7641−2
SMPD2
ZBTB24
109.4
109.6
109.8
110
Position on chr6 (Mb)

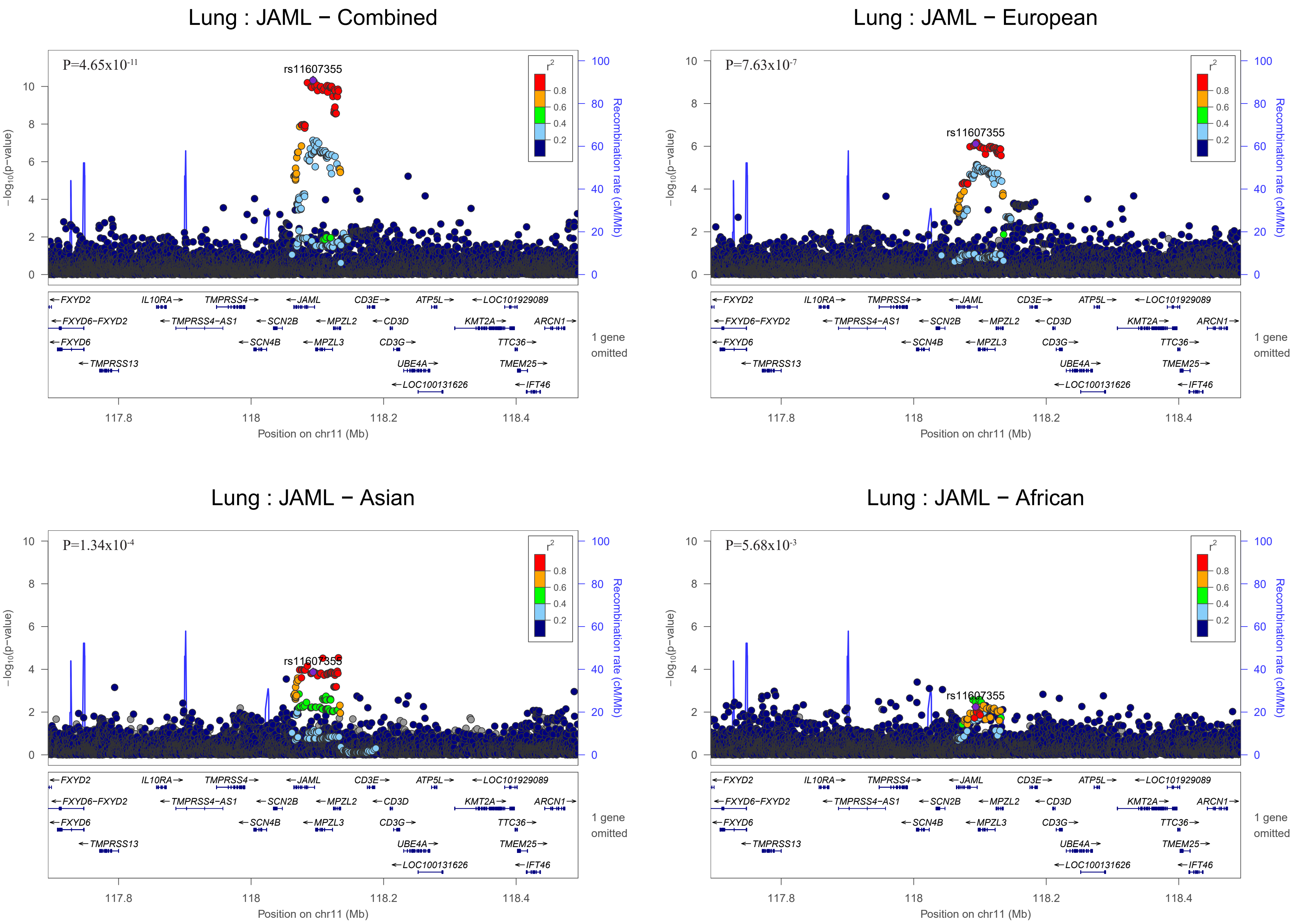
Lung : JAML − Combined
P=4.65x10-11
rs11607355
−log10(p−value)
Recombination rate (cM/Mb)
r2
1 gene omitted
Position on chr11 (Mb)
Lung : JAML − European
P=7.63x10-7
rs11607355
Lung : JAML − Asian
P=1.34x10-4
rs11607355
Lung : JAML − African
P=5.68x10-3
rs11607355
FXYD2
IL10RA
TMPRSS4
JAML
CD3E
ATP5L
LOC101929089
FXYD6−FXYD2
TMPRSS4−AS1
SCN2B
MPZL2
CD3D
KMT2A
ARCN1
FXYD6
SCN4B
MPZL3
CD3G
TTC36
TMPRSS13
UBE4A
TMEM25
LOC100131626
IFT46

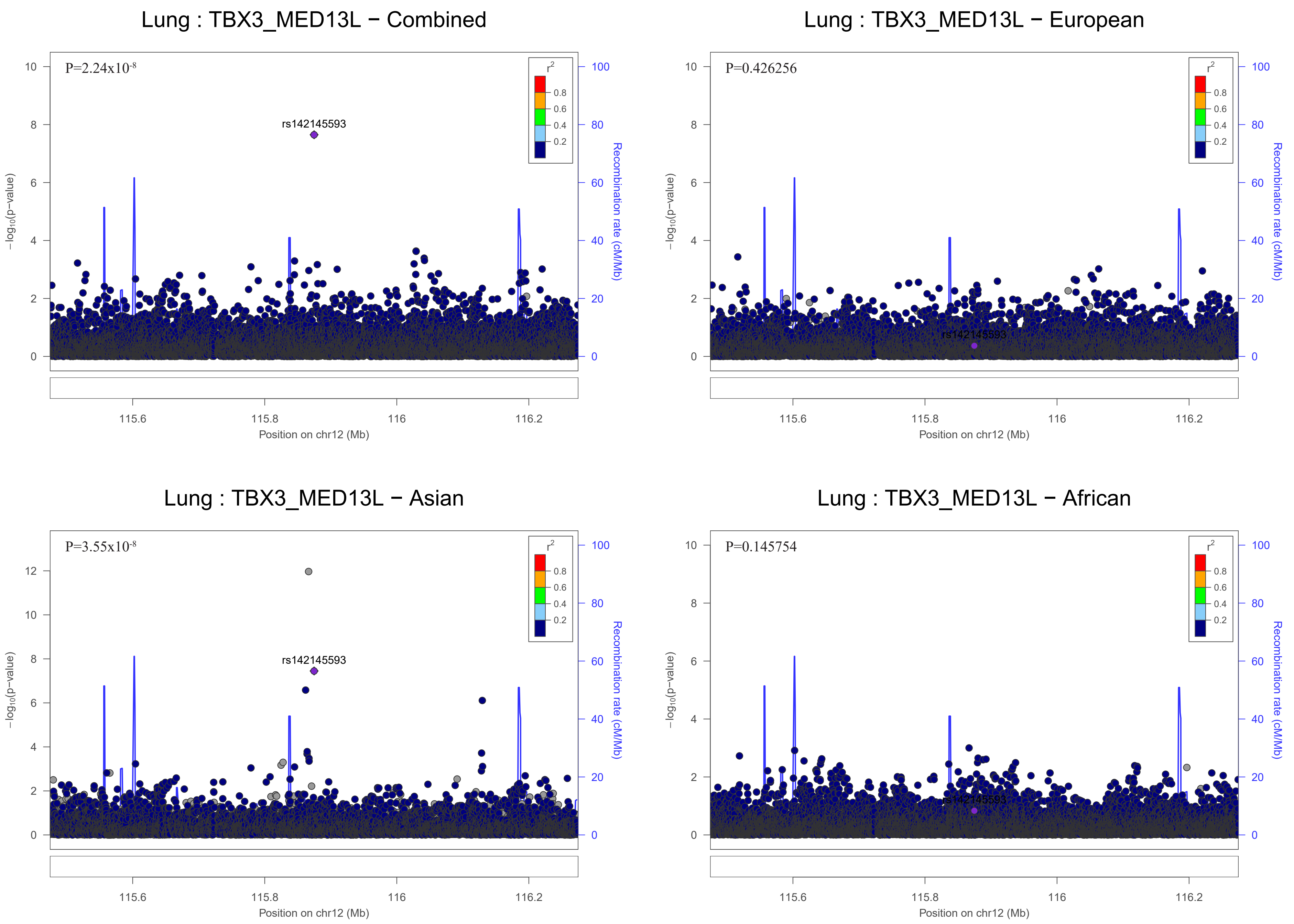

Lung : TBX3_MED13L − Combined
P=2.24x10-8
rs142145593
Lung : TBX3_MED13L − European
P=0.426256
rs142145593
Lung : TBX3_MED13L − Asian
P=3.55x10-8
rs142145593
Lung : TBX3_MED13L − African
P=0.145754
rs142145593
r2
0.8
0.6
0.4
0.2
−log10(p−value)
Recombination rate (cM/Mb)
115.6
115.8
116
116.2
Position on chr12 (Mb)

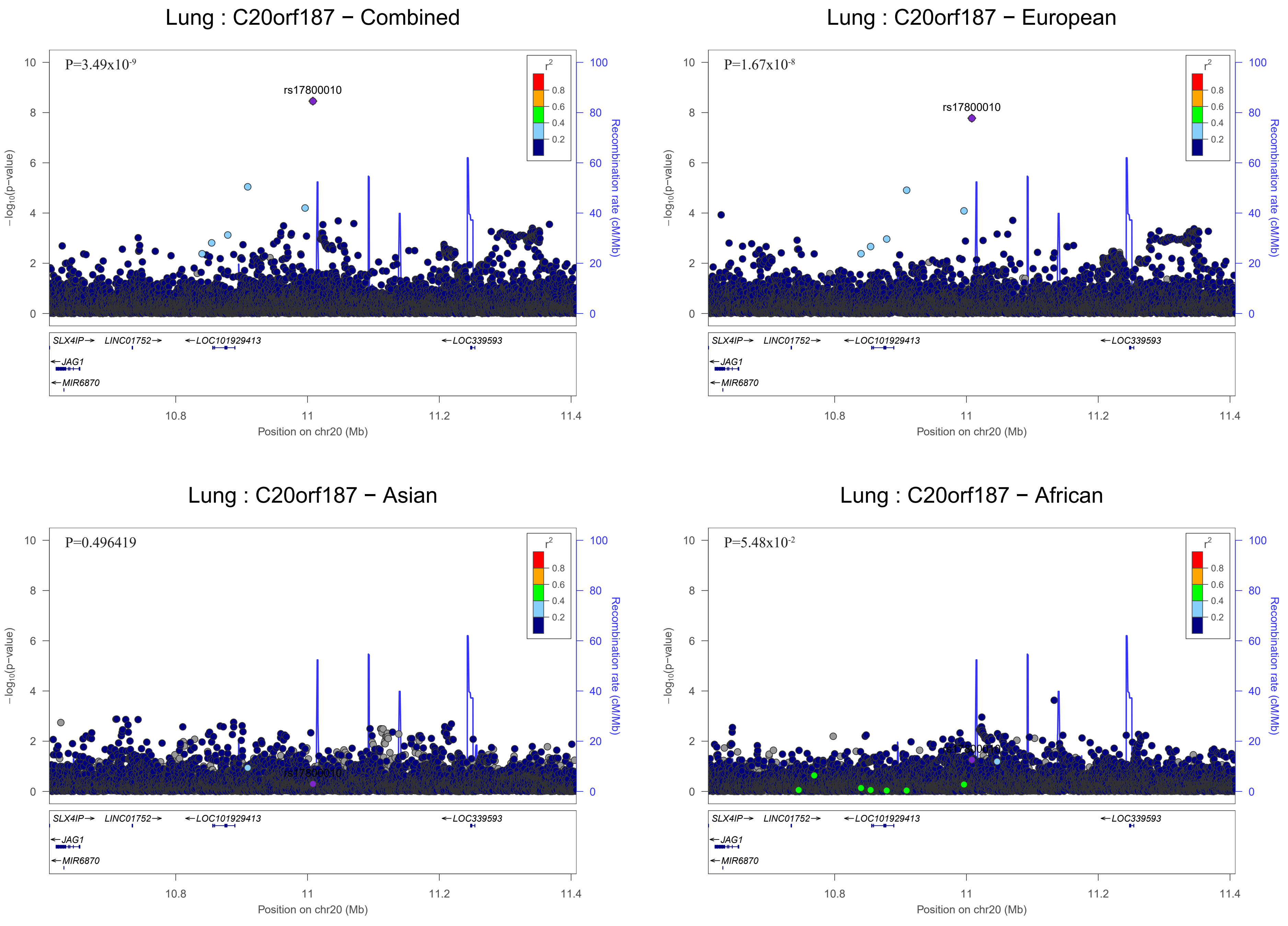

Lung : C20orf187 − Combined
P=3.49x10-9
rs17800010
Lung : C20orf187 − European
P=1.67x10-8
rs17800010
Lung : C20orf187 − Asian
P=0.496419
rs17800010
Lung : C20orf187 − African
P=5.48x10-2
rs17800010
r2
−log10(p−value)
Recombination rate (cM/Mb)
Position on chr20 (Mb)
SLX4IP
LINC01752
LOC101929413
LOC339593
JAG1
MIR6870

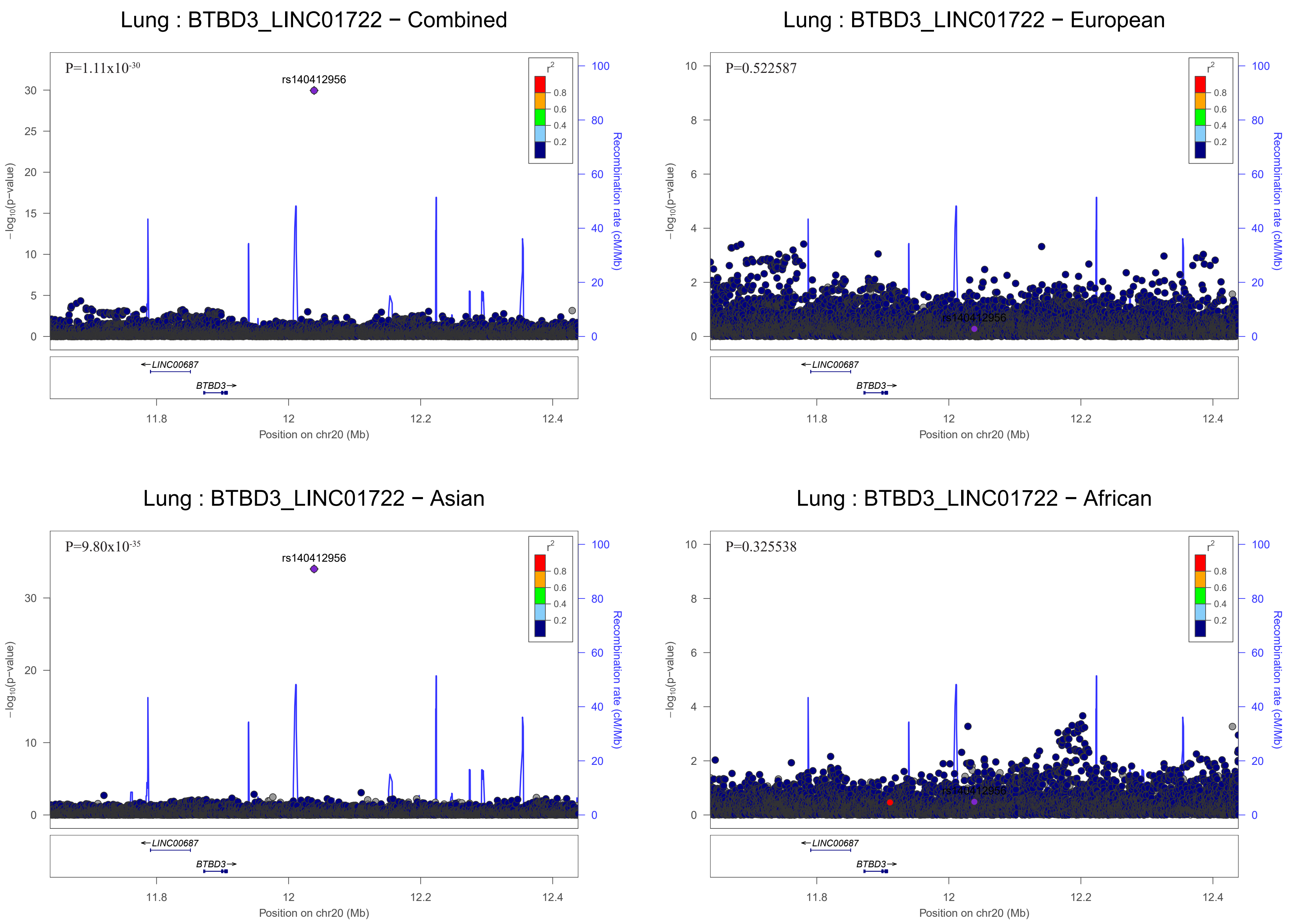

Lung : BTBD3_LINC01722 − Combined
P=1.11x10-30
rs140412956
r2
0.8
0.6
0.4
0.2
−log10(p−value)
Recombination rate (cM/Mb)
LINC00687
BTBD3
11.8
12
12.2
12.4
Position on chr20 (Mb)
Lung : BTBD3_LINC01722 − European
P=0.522587
rs140412956
Position on chr20 (Mb)
Lung : BTBD3_LINC01722 − Asian
P=9.80x10-35
rs140412956
Position on chr20 (Mb)
Lung : BTBD3_LINC01722 − African
P=0.325538
rs140412956
Position on chr20 (Mb)

### Adenocarcinoma

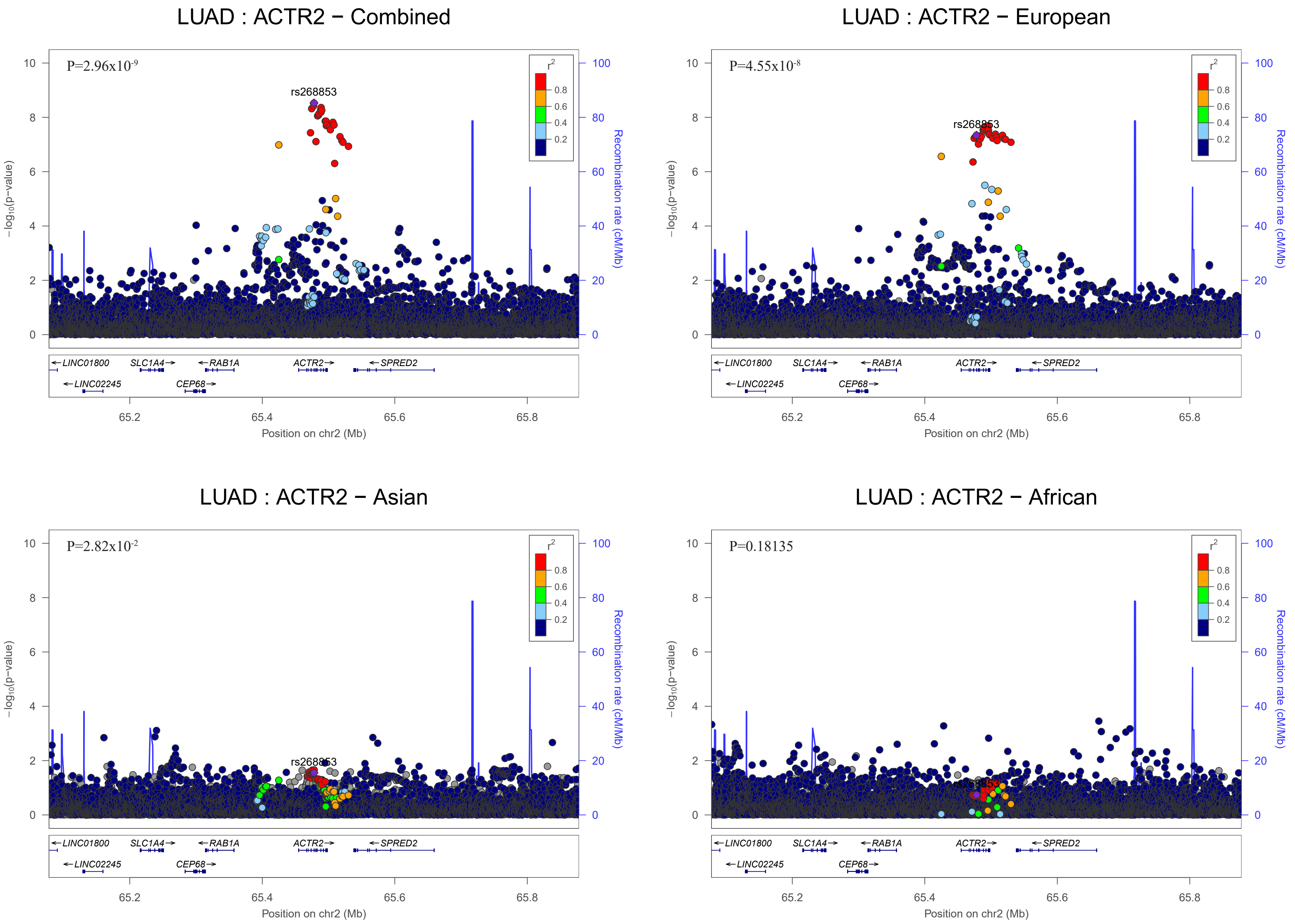

LUAD : ACTR2 − Combined
P=2.96x10-9
rs268853
LUAD : ACTR2 − European
P=4.55x10-8
rs268853
LUAD : ACTR2 − Asian
P=2.82x10-2
rs268853
LUAD : ACTR2 − African
P=0.18135
r2
0.8
0.6
0.4
0.2
−log10(p−value)
Recombination rate (cM/Mb)
LINC01800
SLC1A4
RAB1A
ACTR2
SPRED2
LINC02245
CEP68
65.2
65.4
65.6
65.8
Position on chr2 (Mb)

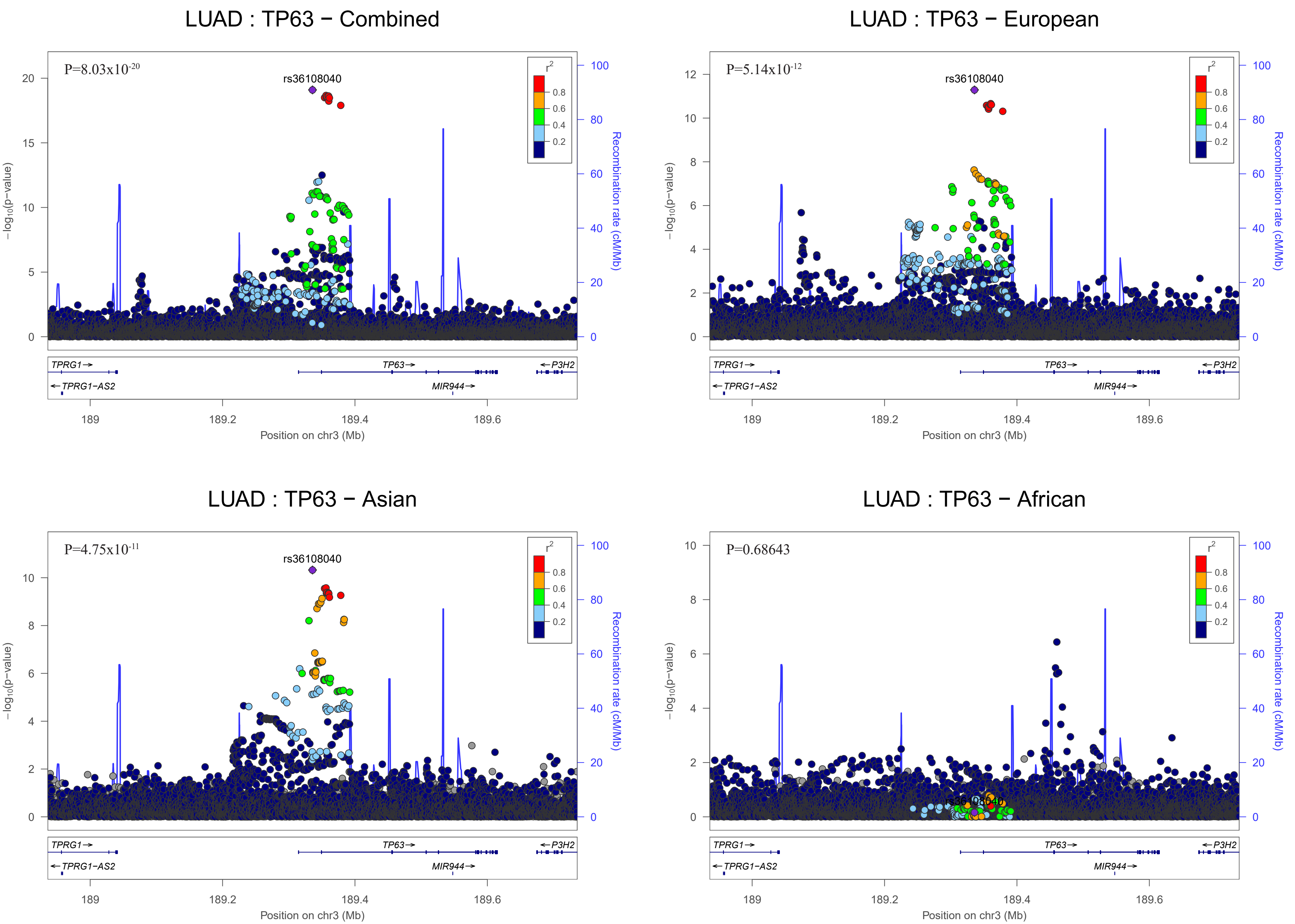
LUAD : TP63 − Combined
P=8.03x10^-20
rs36108040
LUAD : TP63 − European
P=5.14x10^-12
rs36108040
LUAD : TP63 − Asian
P=4.75x10^-11
rs36108040
LUAD : TP63 − African
P=0.68643
r²
0.8
0.6
0.4
0.2
−log10(p−value)
Recombination rate (cM/Mb)
TPRG1→
←TPRG1−AS2
TP63→
MIR944→
←P3H2
189
189.2
189.4
189.6
Position on chr3 (Mb)

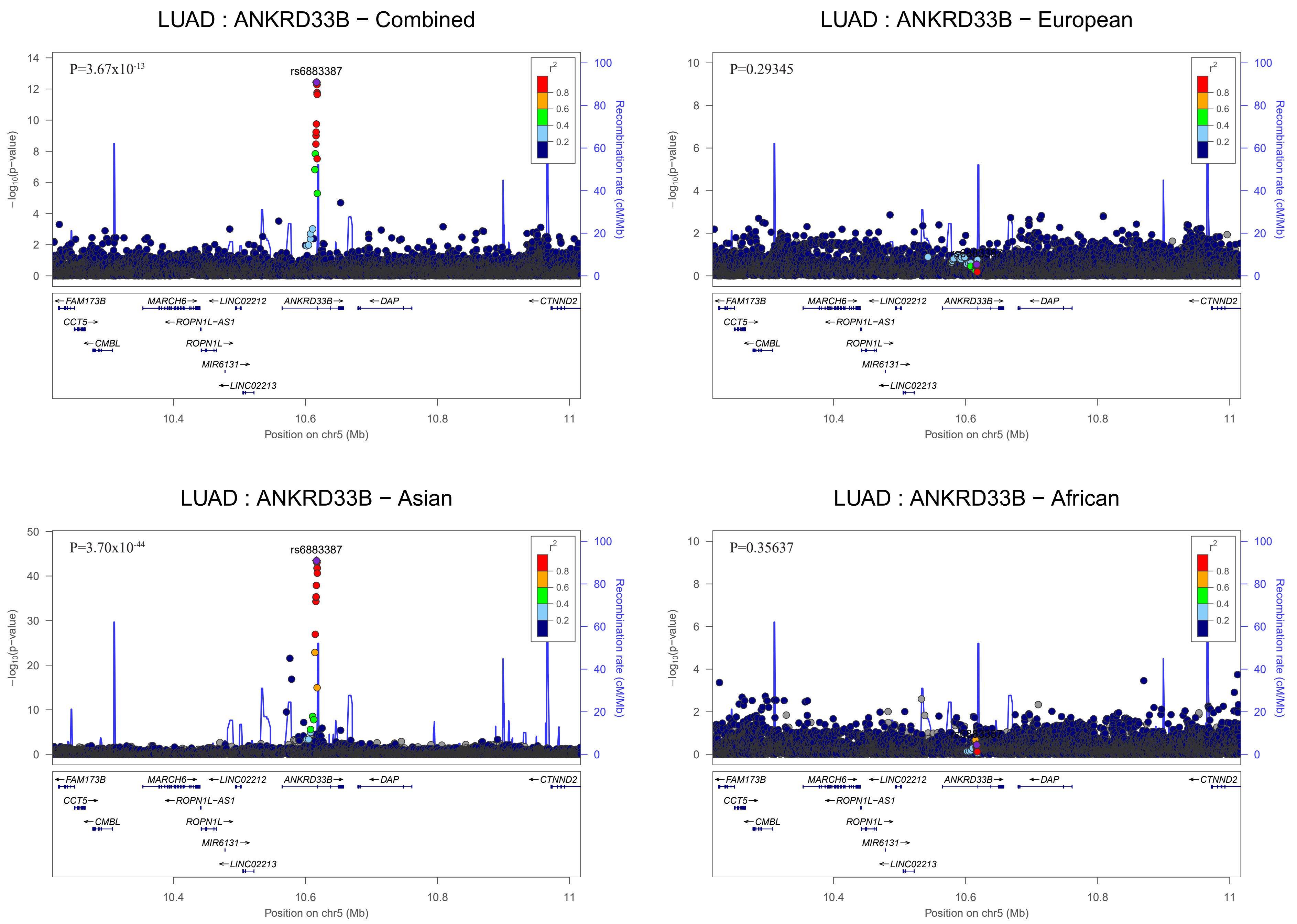

LUAD : ANKRD33B − Combined
P=3.67x10^-13
rs6883387
LUAD : ANKRD33B − European
P=0.29345
LUAD : ANKRD33B − Asian
P=3.70x10^-44
rs6883387
LUAD : ANKRD33B − African
P=0.35637
r²
0.8
0.6
0.4
0.2
−log10(p−value)
Recombination rate (cM/Mb)
FAM173B
CCT5
CMBL
MARCH6
ROPN1L−AS1
ROPN1L
MIR6131
LINC02212
LINC02213
ANKRD33B
DAP
CTNND2
10.4
10.6
10.8
11
Position on chr5 (Mb)

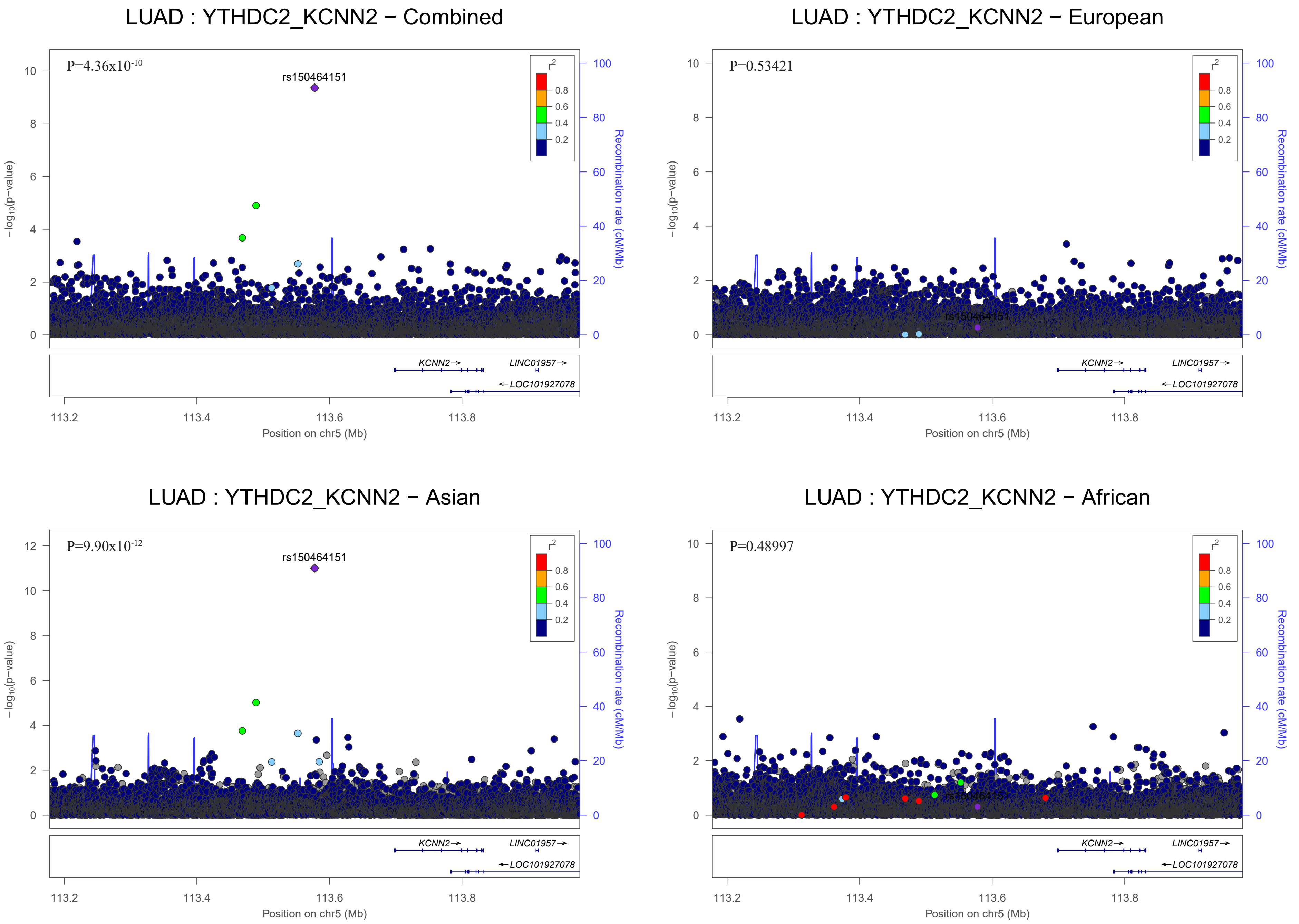

LUAD : YTHDC2_KCNN2 − Combined
P=4.36x10-10
rs150464151
r2
0.8
0.6
0.4
0.2
−log10(p−value)
Recombination rate (cM/Mb)
KCNN2
LINC01957
LOC101927078
Position on chr5 (Mb)
LUAD : YTHDC2_KCNN2 − European
P=0.53421
rs150464151
Position on chr5 (Mb)
LUAD : YTHDC2_KCNN2 − Asian
P=9.90x10-12
rs150464151
Position on chr5 (Mb)
LUAD : YTHDC2_KCNN2 − African
P=0.48997
rs150464151
Position on chr5 (Mb)

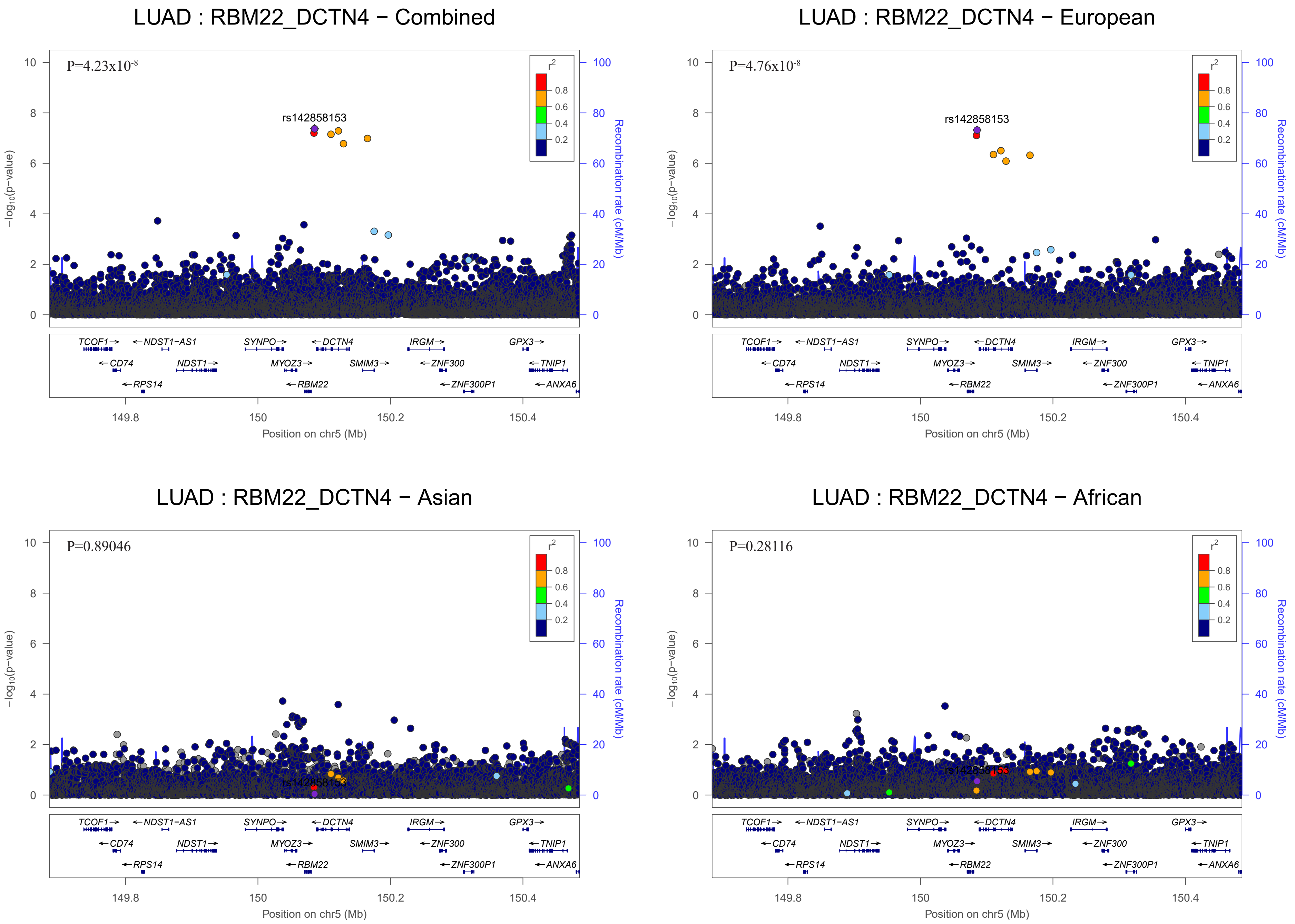
LUAD : RBM22_DCTN4 − Combined
P=4.23x10-8
rs142858153
LUAD : RBM22_DCTN4 − European
P=4.76x10-8
rs142858153
LUAD : RBM22_DCTN4 − Asian
P=0.89046
LUAD : RBM22_DCTN4 − African
P=0.28116
r2
0.8
0.6
0.4
0.2
−log10(p−value)
Recombination rate (cM/Mb)
TCOF1
NDST1−AS1
SYNPO
DCTN4
IRGM
GPX3
CD74
NDST1
MYOZ3
SMIM3
ZNF300
TNIP1
RPS14
RBM22
ZNF300P1
ANXA6
149.8
150
150.2
150.4
Position on chr5 (Mb)

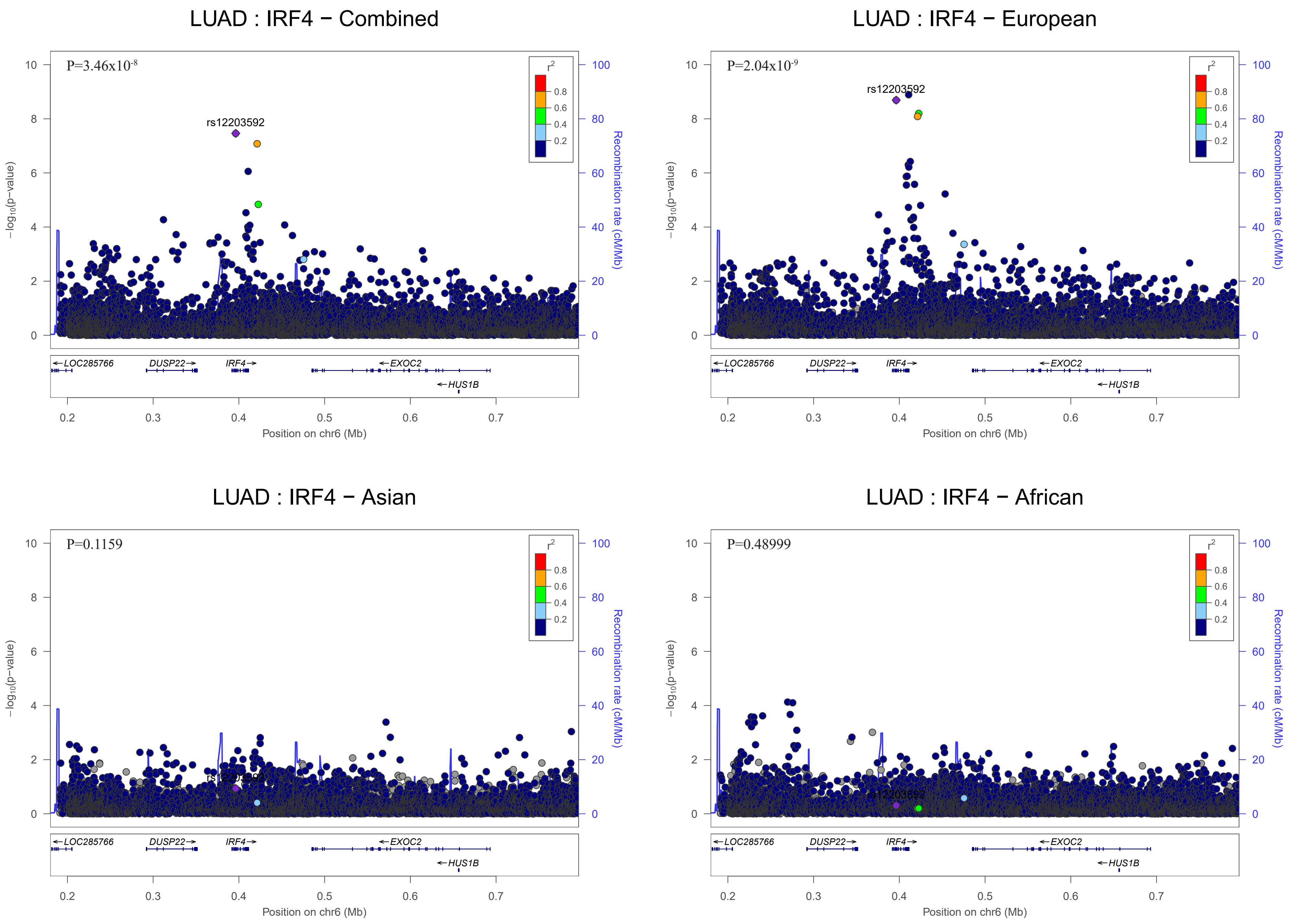
LUAD : IRF4 − Combined
P=3.46x10-8
rs12203592
LUAD : IRF4 − European
P=2.04x10-9
rs12203592
LUAD : IRF4 − Asian
P=0.1159
LUAD : IRF4 − African
P=0.48999
rs12203592
r2
0.8
0.6
0.4
0.2
−log10(p−value)
Recombination rate (cM/Mb)
LOC285766
DUSP22
IRF4
EXOC2
HUS1B
Position on chr6 (Mb)

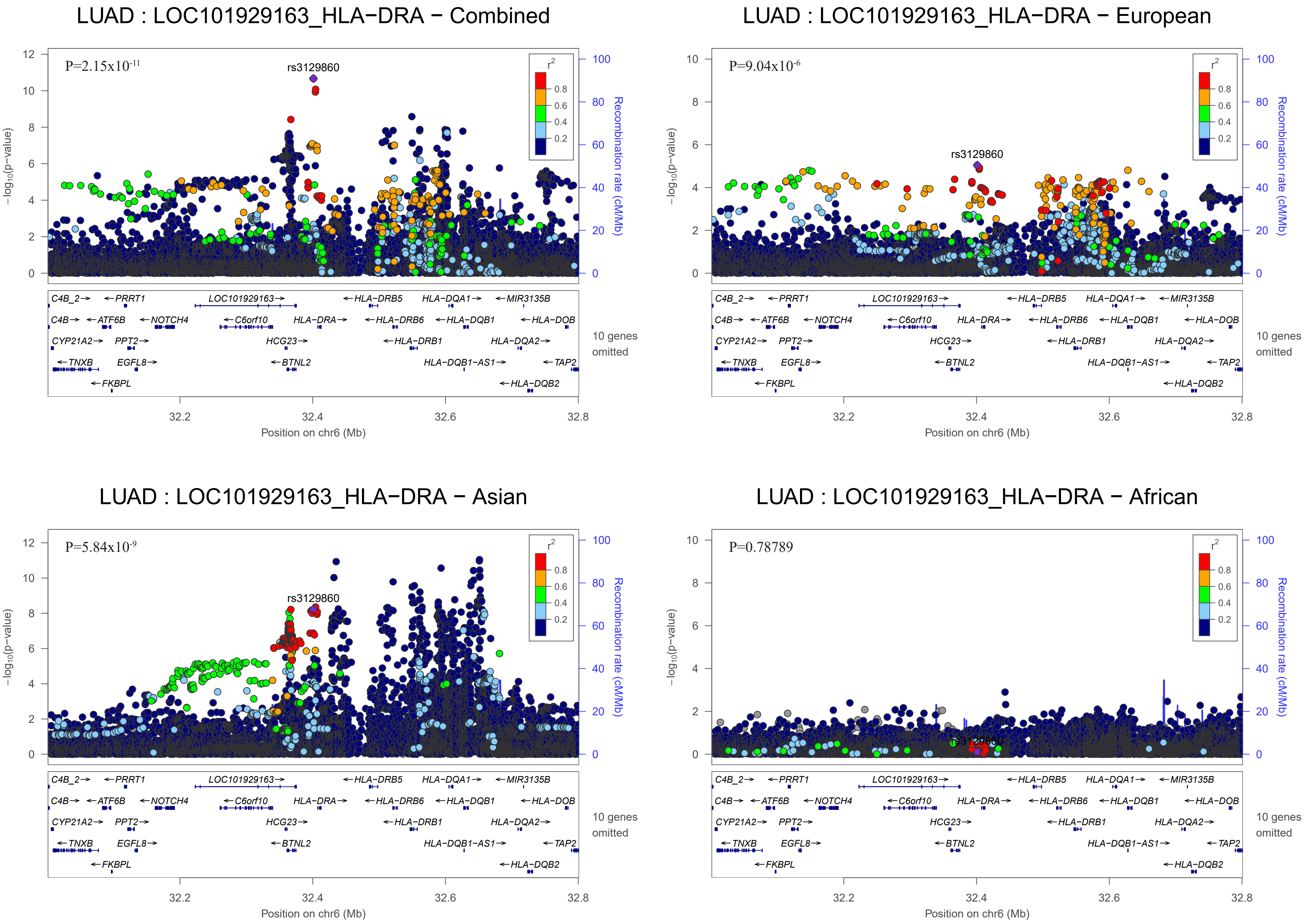
LUAD : LOC101929163_HLA−DRA − Combined
P=2.15x10-11
rs3129860
LUAD : LOC101929163_HLA−DRA − European
P=9.04x10-6
rs3129860
LUAD : LOC101929163_HLA−DRA − Asian
P=5.84x10-9
rs3129860
LUAD : LOC101929163_HLA−DRA − African
P=0.78789
rs3129860
r2
0.8
0.6
0.4
0.2
−log10(p−value)
Recombination rate (cM/Mb)
Position on chr6 (Mb)
32.2
32.4
32.6
32.8
C4B_2
PRRT1
LOC101929163
HLA−DRB5
HLA−DQA1
MIR3135B
C4B
ATF6B
NOTCH4
C6orf10
HLA−DRA
HLA−DRB6
HLA−DQB1
HLA−DOB
CYP21A2
PPT2
HCG23
HLA−DRB1
HLA−DQA2
TNXB
EGFL8
BTNL2
HLA−DQB1−AS1
TAP2
FKBPL
HLA−DQB2
10 genes omitted

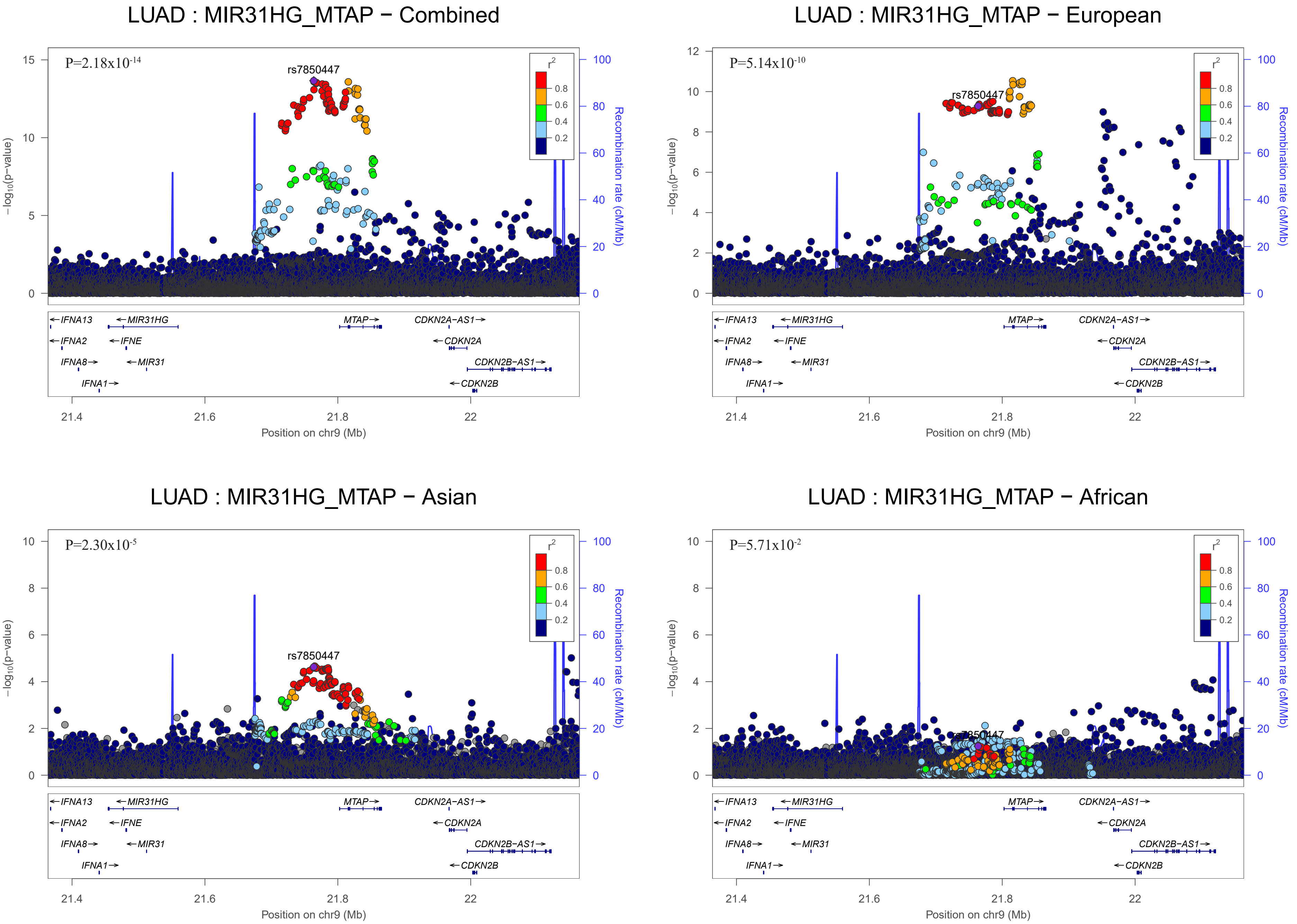

LUAD : MIR31HG_MTAP − Combined
P=2.18x10-14
rs7850447
LUAD : MIR31HG_MTAP − European
P=5.14x10-10
rs7850447
LUAD : MIR31HG_MTAP − Asian
P=2.30x10-5
rs7850447
LUAD : MIR31HG_MTAP − African
P=5.71x10-2
rs7850447
−log10(p−value)
Recombination rate (cM/Mb)
r2
0.8
0.6
0.4
0.2
IFNA13
MIR31HG
MTAP
CDKN2A−AS1
IFNA2
IFNE
CDKN2A
IFNA8
MIR31
CDKN2B−AS1
IFNA1
CDKN2B
21.4
21.6
21.8
22
Position on chr9 (Mb)

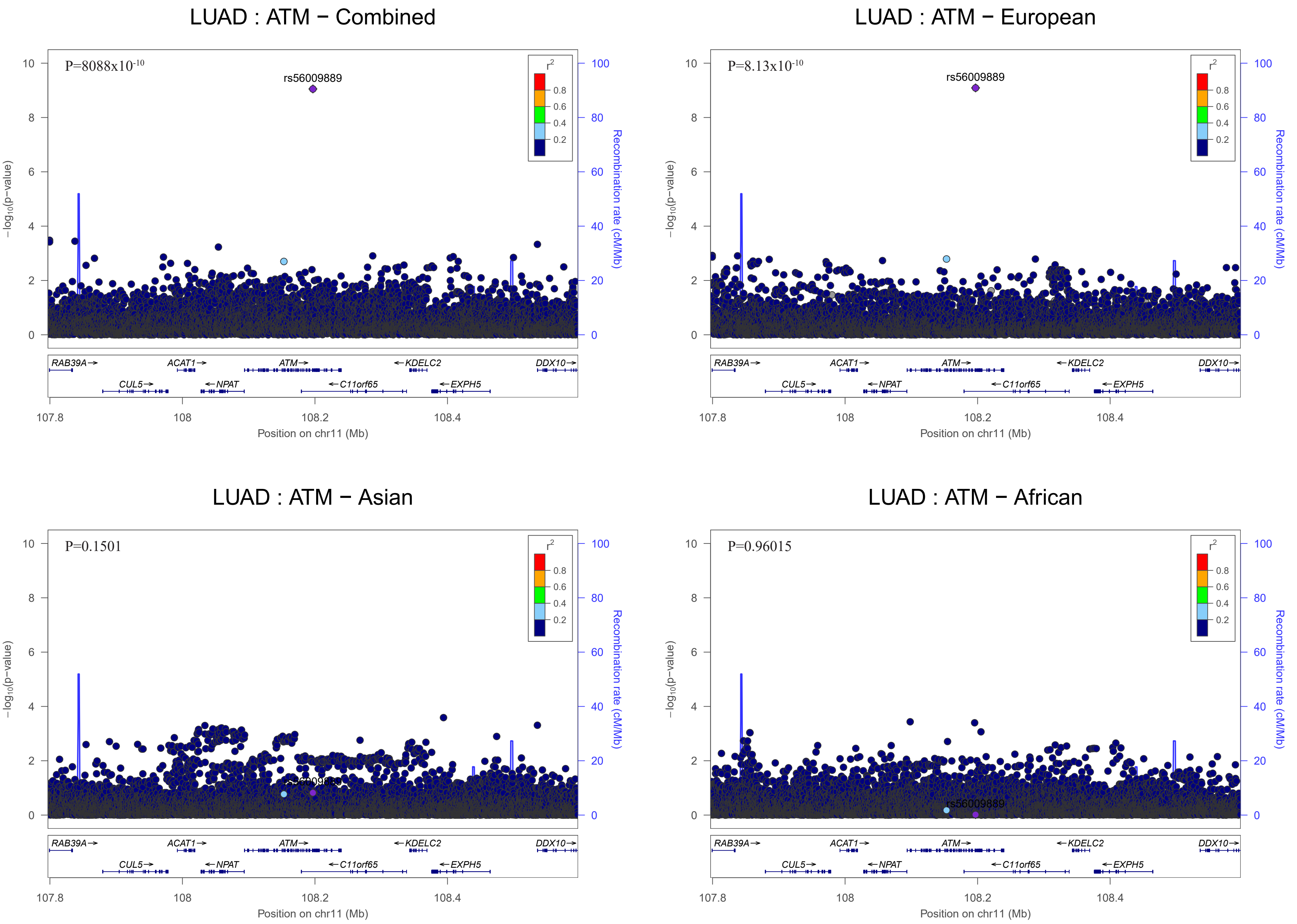

LUAD : ATM − Combined
P=8088x10-10
rs56009889
LUAD : ATM − European
P=8.13x10-10
rs56009889
LUAD : ATM − Asian
P=0.1501
rs56009889
LUAD : ATM − African
P=0.96015
rs56009889
r2
0.8
0.6
0.4
0.2
−log10(p−value)
Recombination rate (cM/Mb)
RAB39A
ACAT1
ATM
KDELC2
DDX10
CUL5
NPAT
C11orf65
EXPH5
107.8
108
108.2
108.4
Position on chr11 (Mb)

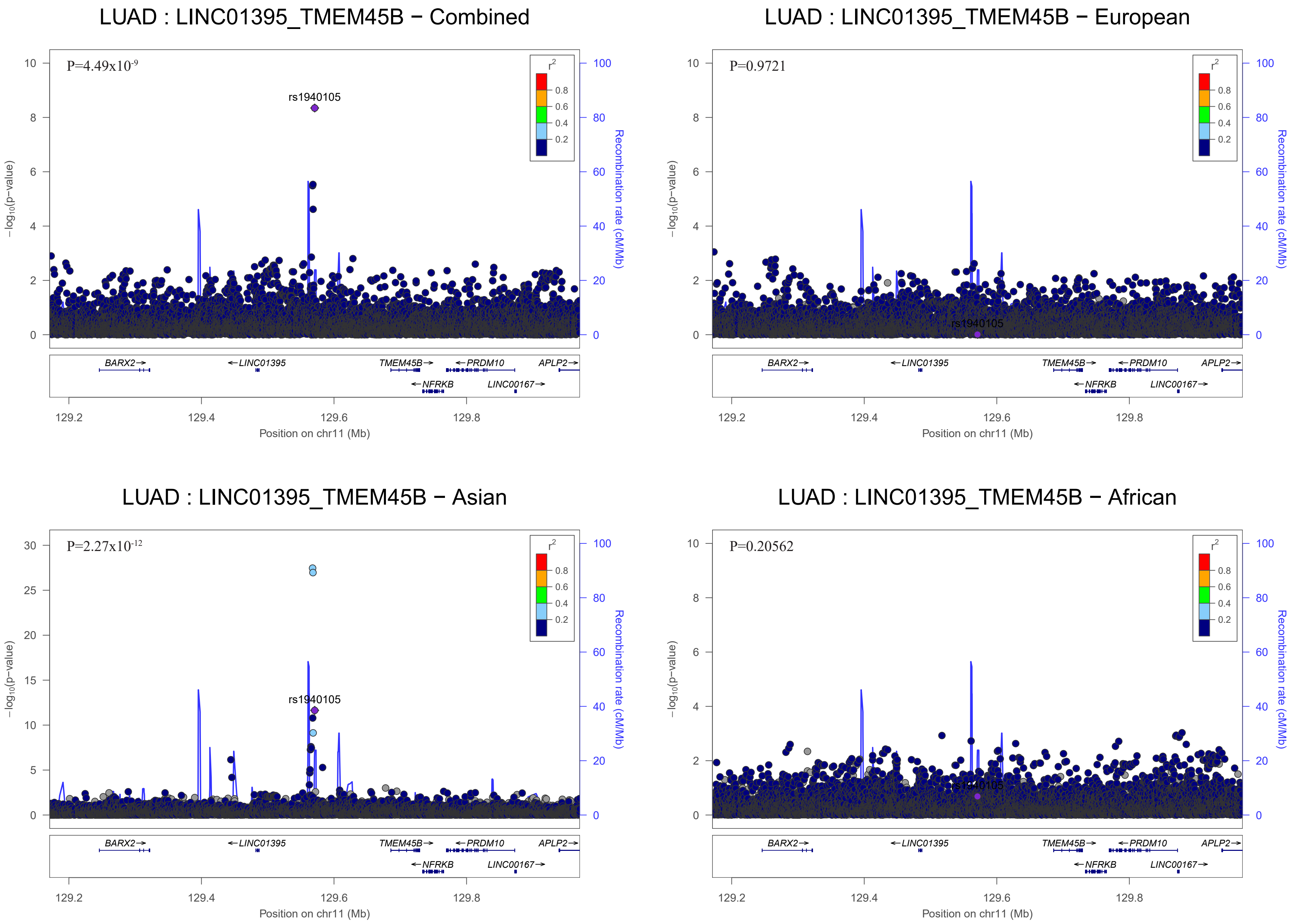
LUAD : LINC01395_TMEM45B − Combined
P=4.49x10-9
rs1940105
−log10(p−value)
Recombination rate (cM/Mb)
r2
0.8
0.6
0.4
0.2
BARX2→
←LINC01395
TMEM45B→
←PRDM10
APLP2→
←NFRKB
LINC00167→
129.2
129.4
129.6
129.8
Position on chr11 (Mb)
LUAD : LINC01395_TMEM45B − European
P=0.9721
rs1940105
LUAD : LINC01395_TMEM45B − Asian
P=2.27x10-12
rs1940105
LUAD : LINC01395_TMEM45B − African
P=0.20562
rs1940105

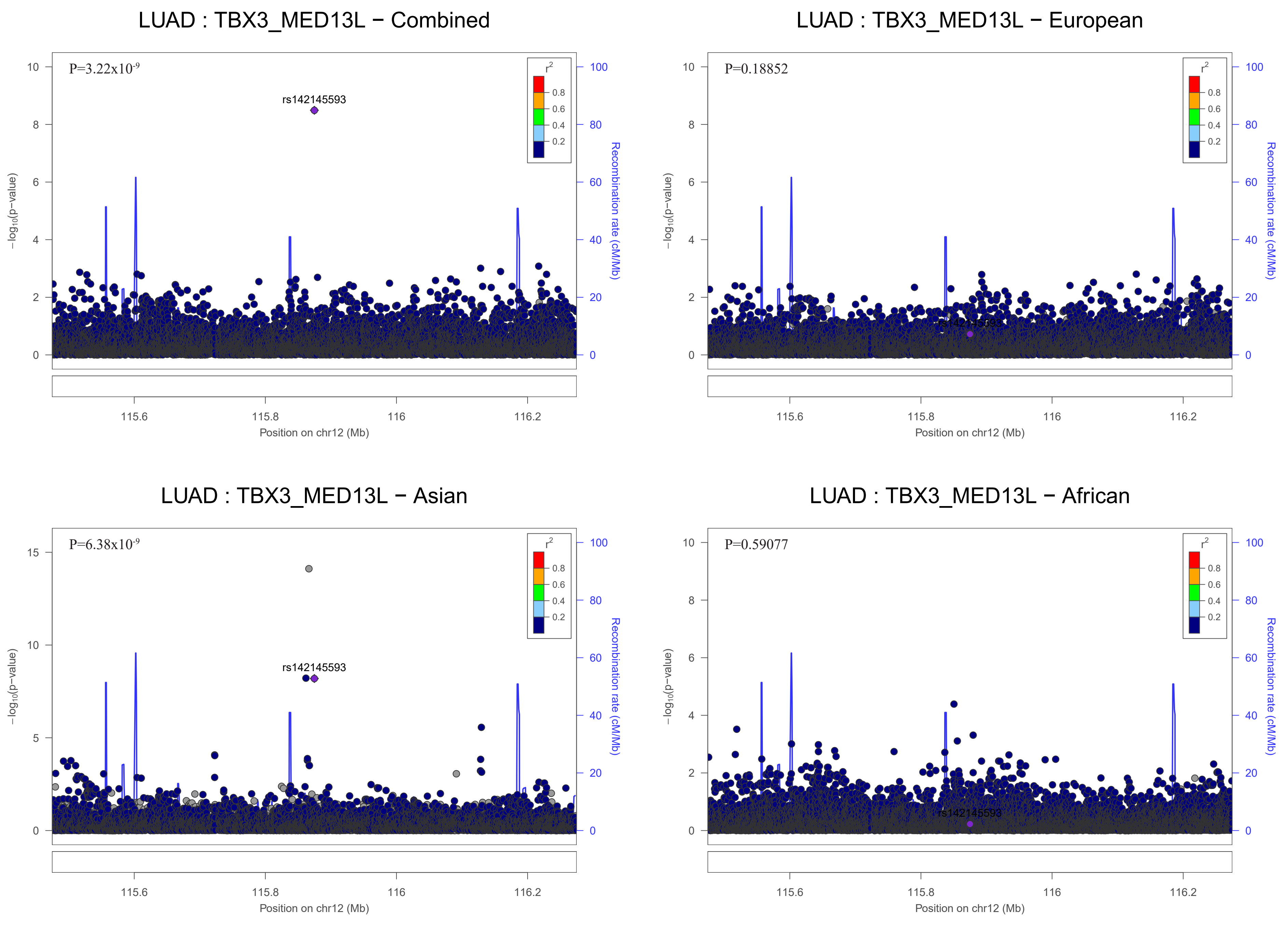

LUAD : TBX3_MED13L − Combined
P=3.22x10-9
rs142145593
−log10(p−value)
Recombination rate (cM/Mb)
Position on chr12 (Mb)
LUAD : TBX3_MED13L − European
P=0.18852
rs142145593
−log10(p−value)
Recombination rate (cM/Mb)
Position on chr12 (Mb)
LUAD : TBX3_MED13L − Asian
P=6.38x10-9
rs142145593
−log10(p−value)
Recombination rate (cM/Mb)
Position on chr12 (Mb)
LUAD : TBX3_MED13L − African
P=0.59077
rs142145593
−log10(p−value)
Recombination rate (cM/Mb)
Position on chr12 (Mb)

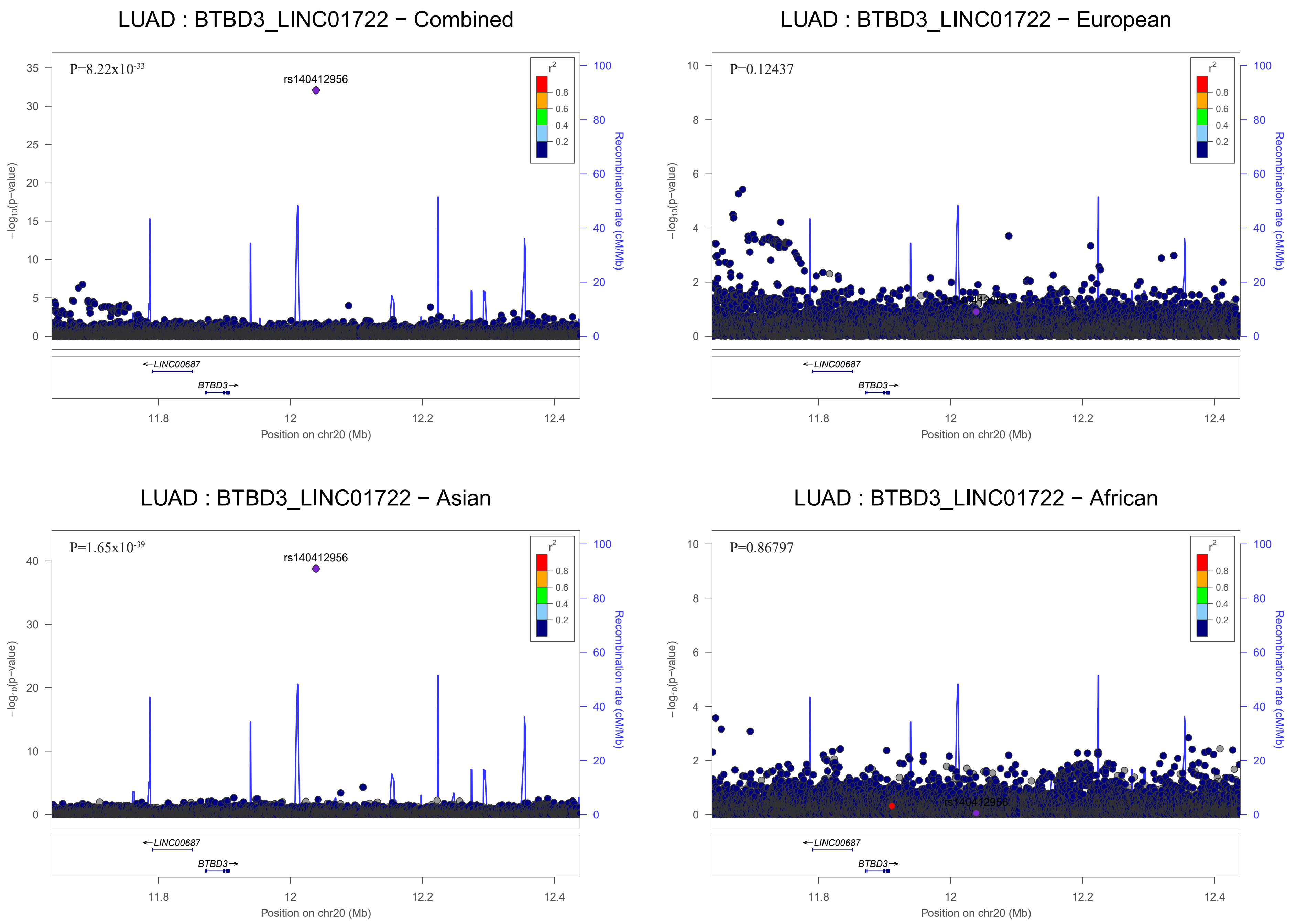

LUAD : BTBD3_LINC01722 − Combined
P=8.22x10-33
rs140412956
LUAD : BTBD3_LINC01722 − European
P=0.12437
LUAD : BTBD3_LINC01722 − Asian
P=1.65x10-39
rs140412956
LUAD : BTBD3_LINC01722 − African
P=0.86797
rs140412956
r2
0.8
0.6
0.4
0.2
−log10(p−value)
Recombination rate (cM/Mb)
LINC00687
BTBD3
11.8
12
12.2
12.4
Position on chr20 (Mb)

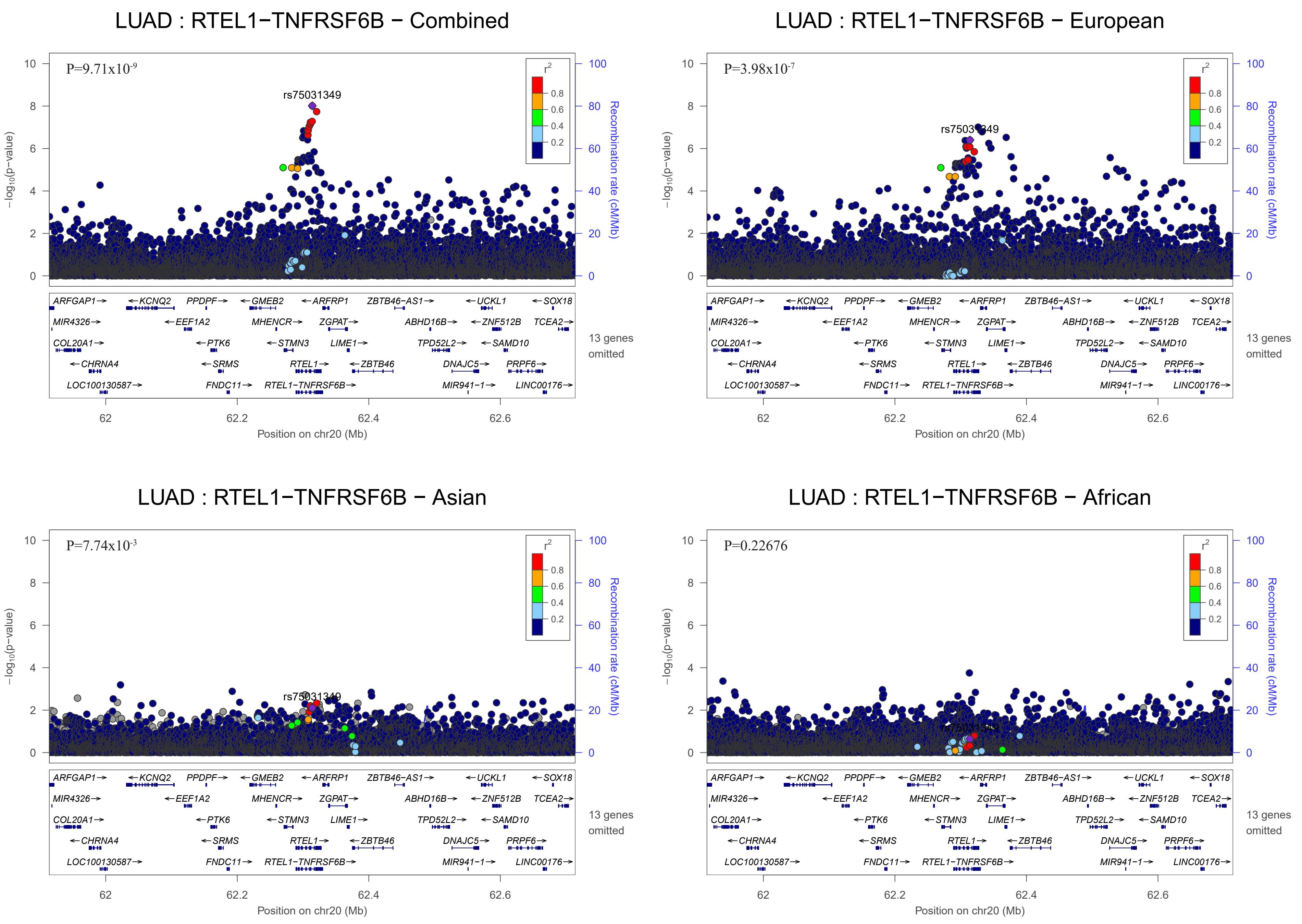
LUAD : RTEL1−TNFRSF6B − Combined
P=9.71x10-9
rs75031349
LUAD : RTEL1−TNFRSF6B − European
P=3.98x10-7
rs75031349
LUAD : RTEL1−TNFRSF6B − Asian
P=7.74x10-3
rs75031349
LUAD : RTEL1−TNFRSF6B − African
P=0.22676
−log10(p−value)
Recombination rate (cM/Mb)
r2
0.8
0.6
0.4
0.2
ARFGAP1→ ←KCNQ2 PPDPF→ ←GMEB2 ←ARFRP1 ZBTB46−AS1→ ←UCKL1 ←SOX18
MIR4326→ ←EEF1A2 MHENCR→ ZGPAT→ ABHD16B→ ←ZNF512B TCEA2→
COL20A1→ ←PTK6 ←STMN3 LIME1→ TPD52L2→ ←SAMD10
←CHRNA4 ←SRMS RTEL1→ ←ZBTB46 DNAJC5→ PRPF6→
LOC100130587→ FNDC11→ RTEL1−TNFRSF6B→ MIR941−1→ LINC00176→
13 genes omitted
62
62.2
62.4
62.6
Position on chr20 (Mb)

### Squamous Cell Carcinoma

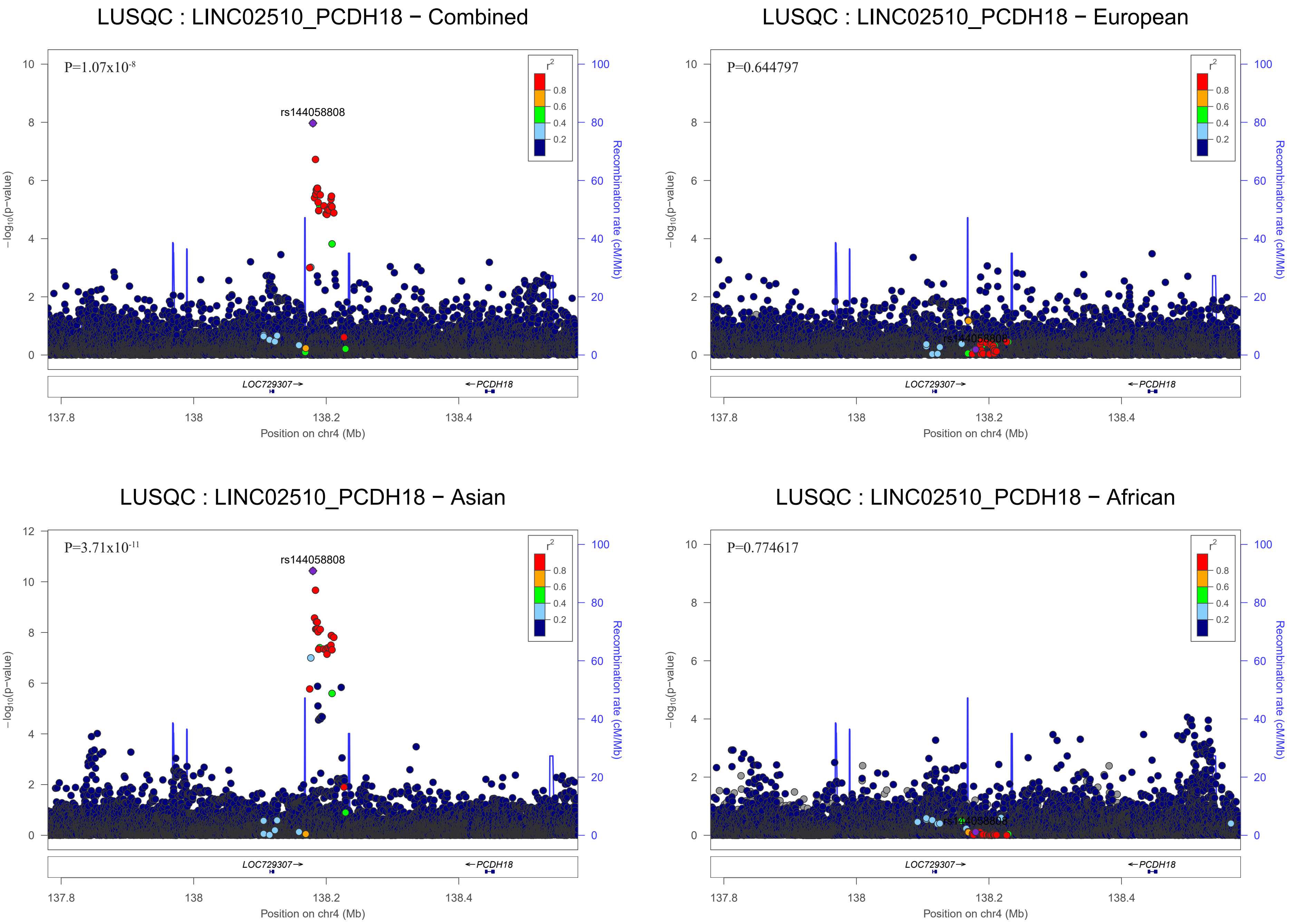

LUSQC : LINC02510_PCDH18 − Combined
P=1.07x10-8
rs144058808
LUSQC : LINC02510_PCDH18 − European
P=0.644797
rs144058808
LUSQC : LINC02510_PCDH18 − Asian
P=3.71x10-11
rs144058808
LUSQC : LINC02510_PCDH18 − African
P=0.774617
rs144058808
r2
0.8
0.6
0.4
0.2
−log10(p−value)
Recombination rate (cM/Mb)
LOC729307→
←PCDH18
137.8
138
138.2
138.4
Position on chr4 (Mb)

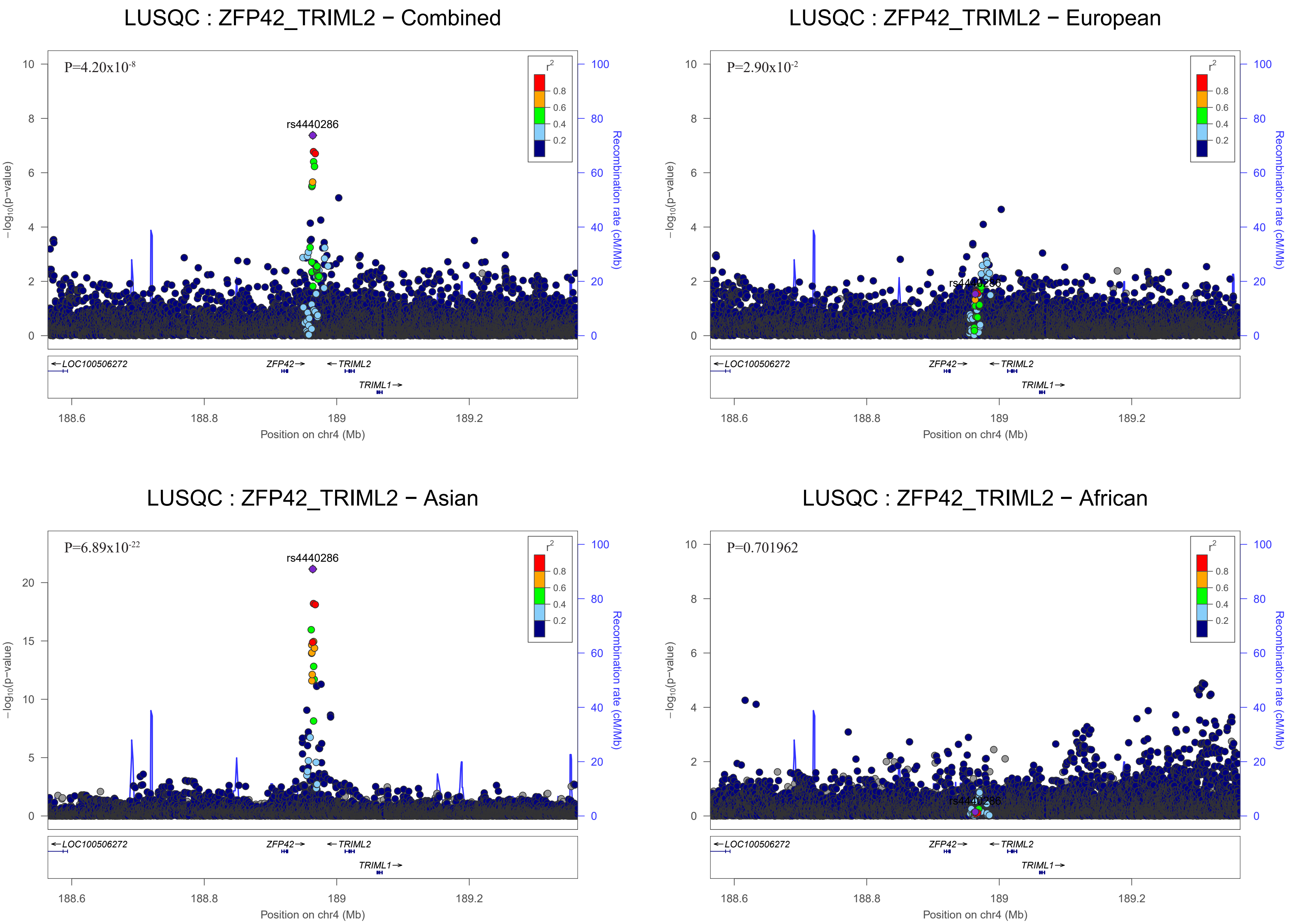
LUSQC : ZFP42_TRIML2 − Combined
P=4.20x10^-8
rs4440286
r^2
0.8
0.6
0.4
0.2
−log10(p−value)
Recombination rate (cM/Mb)
LOC100506272
ZFP42
TRIML2
TRIML1
188.6
188.8
189
189.2
Position on chr4 (Mb)
LUSQC : ZFP42_TRIML2 − European
P=2.90x10^-2
LUSQC : ZFP42_TRIML2 − Asian
P=6.89x10^-22
LUSQC : ZFP42_TRIML2 − African
P=0.701962

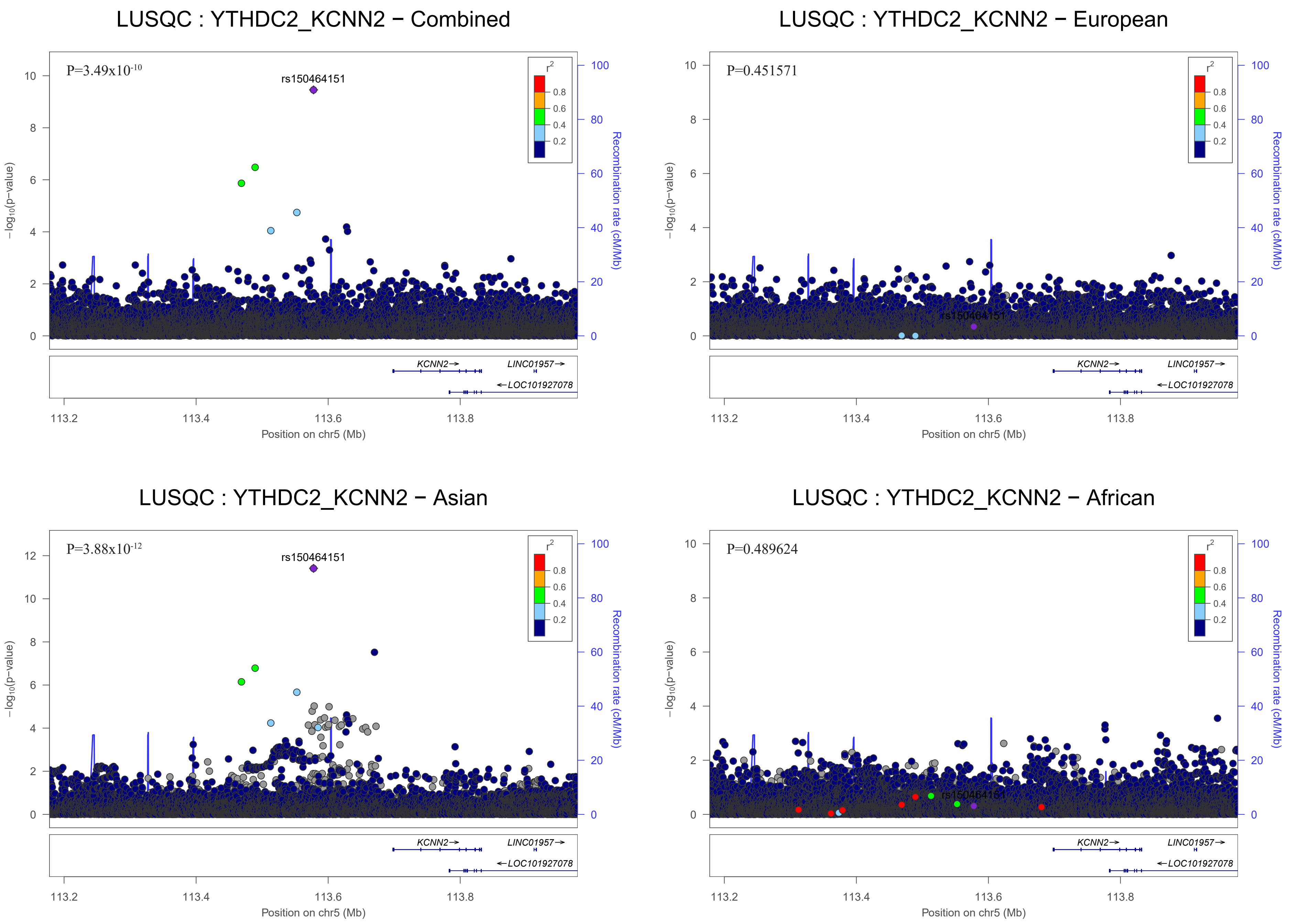
LUSQC : YTHDC2_KCNN2 − Combined
P=3.49x10-10
rs150464151
LUSQC : YTHDC2_KCNN2 − European
P=0.451571
rs150464151
LUSQC : YTHDC2_KCNN2 − Asian
P=3.88x10-12
rs150464151
LUSQC : YTHDC2_KCNN2 − African
P=0.489624
rs150464151
r2
0.8
0.6
0.4
0.2
−log10(p−value)
Recombination rate (cM/Mb)
KCNN2
LINC01957
LOC101927078
113.2
113.4
113.6
113.8
Position on chr5 (Mb)

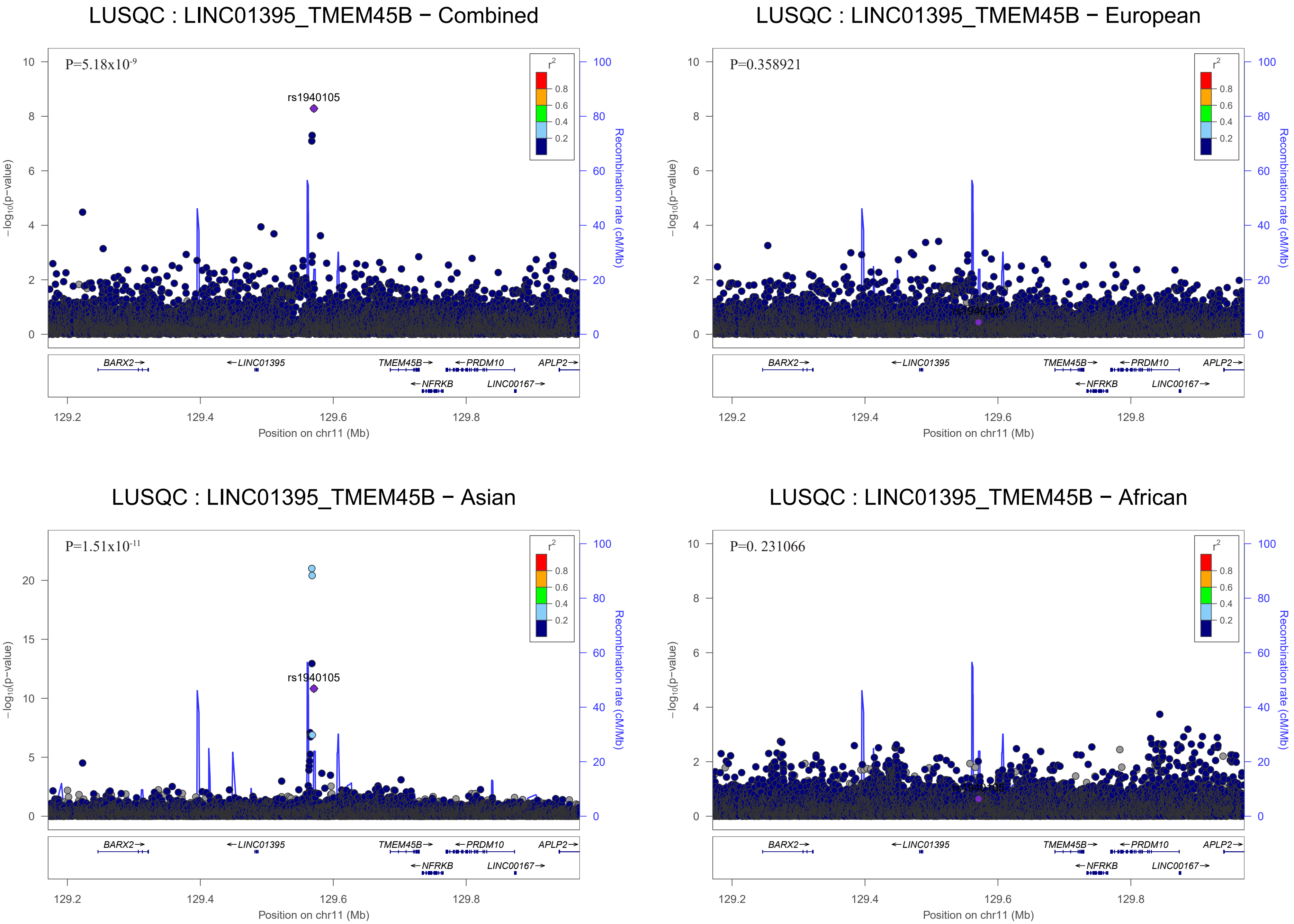

LUSQC : LINC01395_TMEM45B − Combined
P=5.18x10-9
rs1940105
r2
0.8
0.6
0.4
0.2
−log10(p−value)
Recombination rate (cM/Mb)
BARX2
LINC01395
TMEM45B
PRDM10
APLP2
NFRKB
LINC00167
Position on chr11 (Mb)
LUSQC : LINC01395_TMEM45B − European
P=0.358921
rs1940105
LUSQC : LINC01395_TMEM45B − Asian
P=1.51x10-11
rs1940105
LUSQC : LINC01395_TMEM45B − African
P=0. 231066
rs1940105

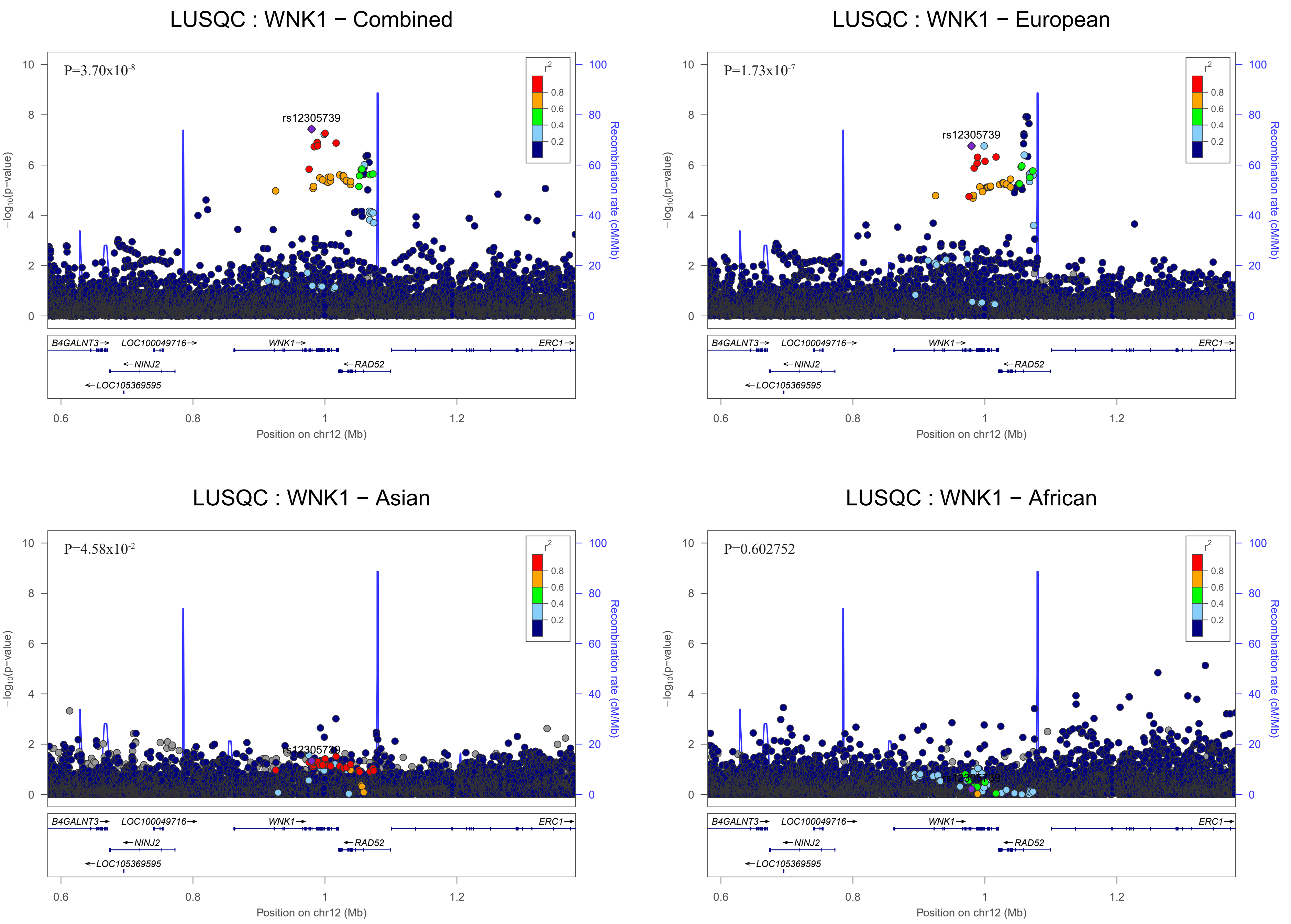

LUSQC : WNK1 − Combined
P=3.70x10-8
rs12305739
LUSQC : WNK1 − European
P=1.73x10-7
rs12305739
LUSQC : WNK1 − Asian
P=4.58x10-2
rs12305739
LUSQC : WNK1 − African
P=0.602752
r2
0.8
0.6
0.4
0.2
−log10(p−value)
Recombination rate (cM/Mb)
B4GALNT3
LOC100049716
WNK1
ERC1
NINJ2
RAD52
LOC105369595
0.6
0.8
1
1.2
Position on chr12 (Mb)

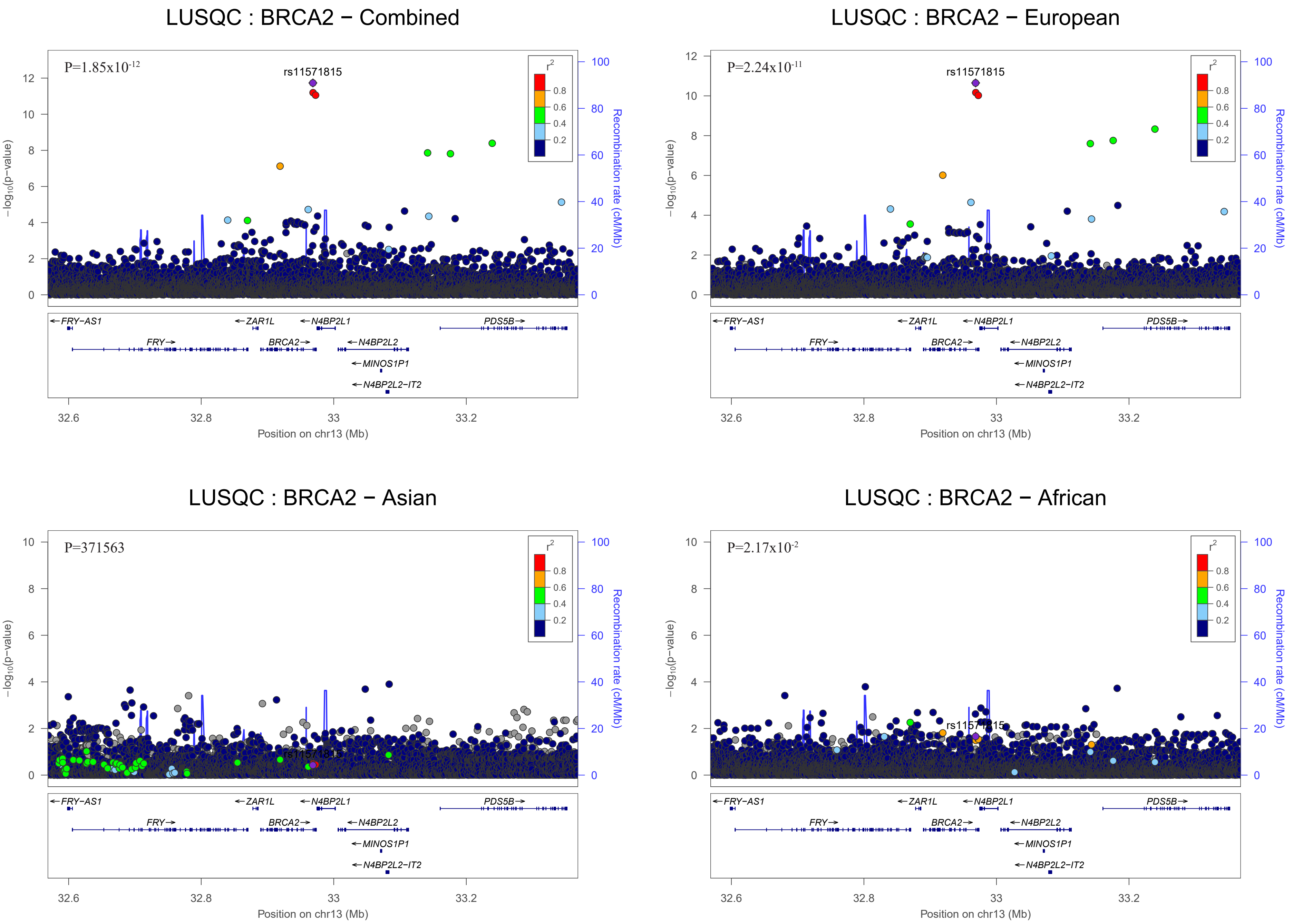
LUSQC : BRCA2 − Combined
P=1.85x10^-12
rs11571815
LUSQC : BRCA2 − European
P=2.24x10^-11
rs11571815
LUSQC : BRCA2 − Asian
P=371563
LUSQC : BRCA2 − African
P=2.17x10^-2
rs11571815
r²
0.8
0.6
0.4
0.2
−log10(p−value)
Recombination rate (cM/Mb)
FRY−AS1
FRY
ZAR1L
BRCA2
N4BP2L1
N4BP2L2
MINOS1P1
N4BP2L2−IT2
PDS5B
32.6
32.8
33
33.2
Position on chr13 (Mb)

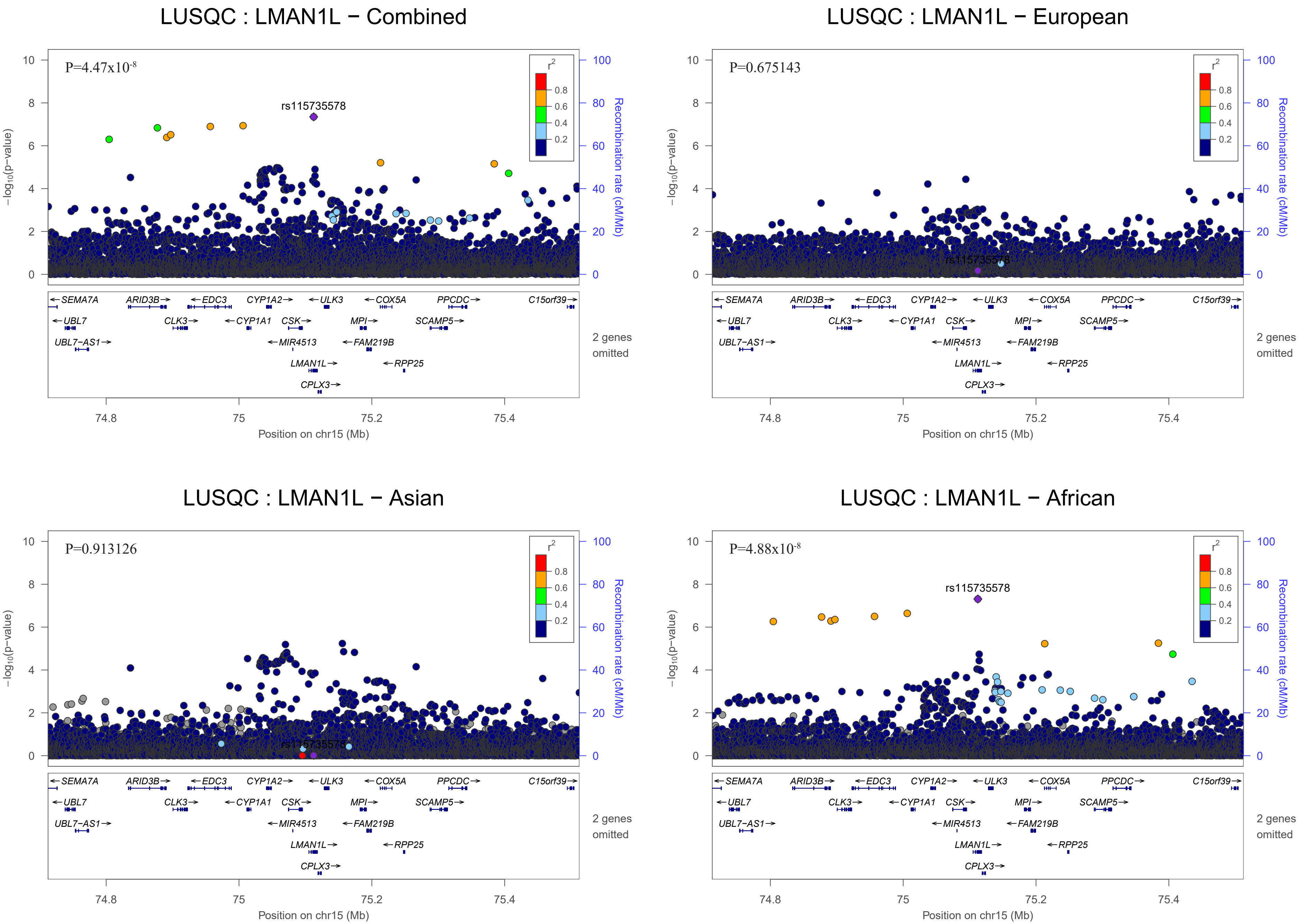
LUSQC : LMAN1L − Combined
P=4.47x10-8
rs115735578
LUSQC : LMAN1L − European
P=0.675143
LUSQC : LMAN1L − Asian
P=0.913126
LUSQC : LMAN1L − African
P=4.88x10-8
rs115735578
r2
0.8
0.6
0.4
0.2
−log10(p−value)
Recombination rate (cM/Mb)
Position on chr15 (Mb)
74.8
75
75.2
75.4
SEMA7A
ARID3B
EDC3
CYP1A2
ULK3
COX5A
PPCDC
C15orf39
UBL7
CLK3
CYP1A1
CSK
MPI
SCAMP5
UBL7−AS1
MIR4513
FAM219B
LMAN1L
RPP25
CPLX3
2 genes omitted

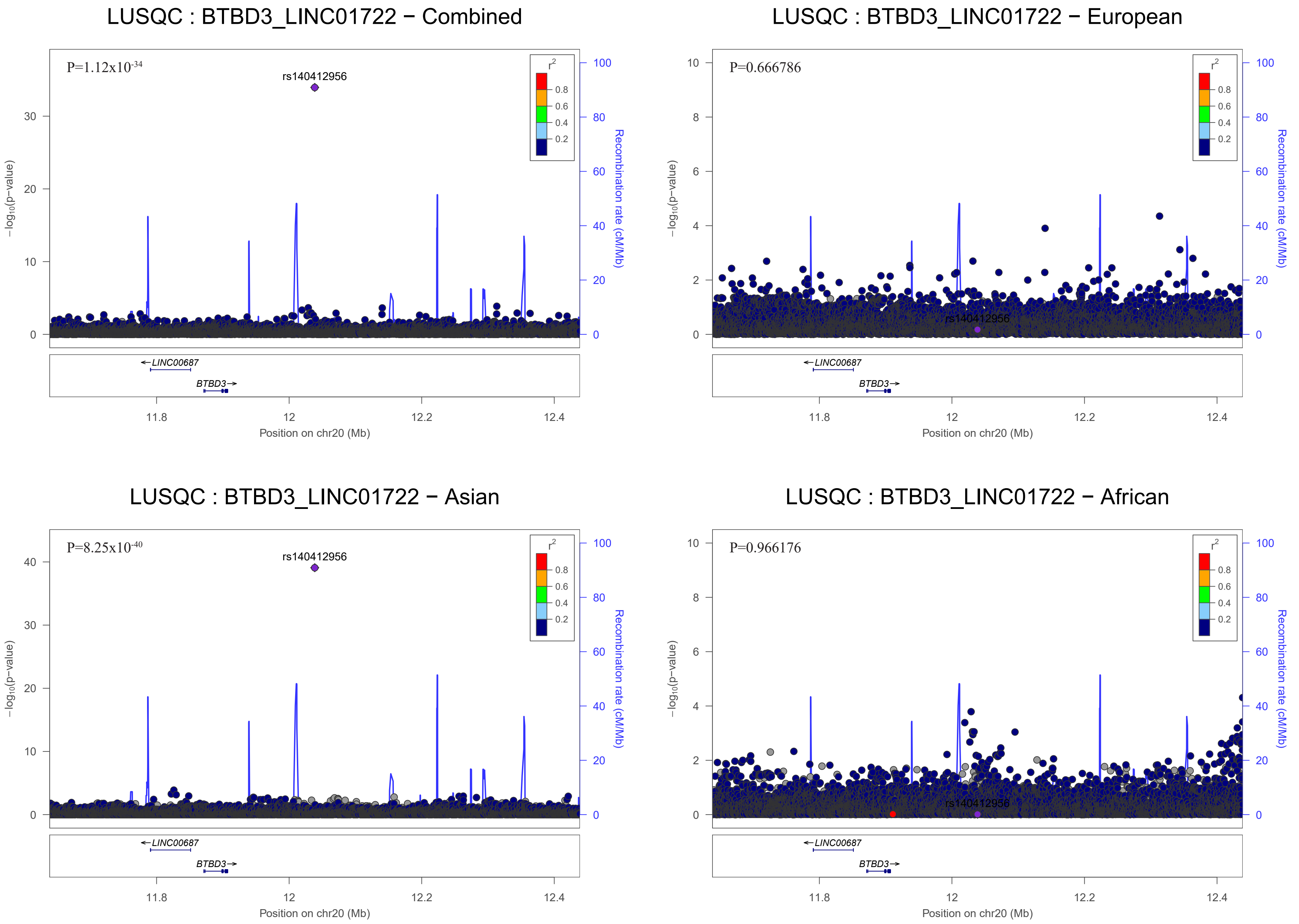

LUSQC : BTBD3_LINC01722 − Combined
P=1.12x10-34
rs140412956
LUSQC : BTBD3_LINC01722 − European
P=0.666786
rs140412956
LUSQC : BTBD3_LINC01722 − Asian
P=8.25x10-40
rs140412956
LUSQC : BTBD3_LINC01722 − African
P=0.966176
rs140412956
r2
0.8
0.6
0.4
0.2
−log10(p−value)
Recombination rate (cM/Mb)
LINC00687
BTBD3
Position on chr20 (Mb)
11.8
12
12.2
12.4

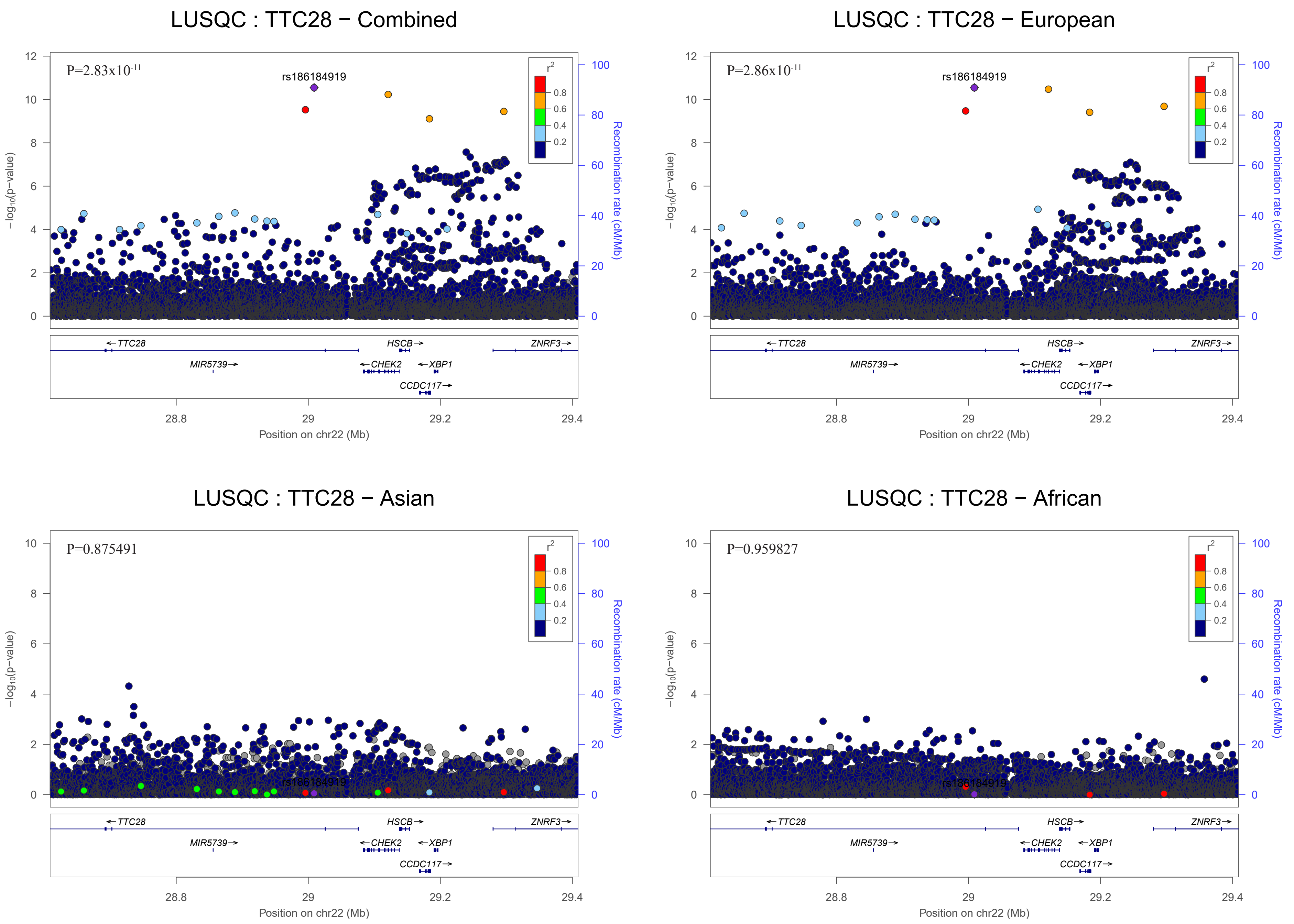
LUSQC : TTC28 − Combined
P=2.83x10-11
rs186184919
LUSQC : TTC28 − European
P=2.86x10-11
rs186184919
LUSQC : TTC28 − Asian
P=0.875491
rs186184919
LUSQC : TTC28 − African
P=0.959827
rs186184919
r2
0.8
0.6
0.4
0.2
−log10(p−value)
Recombination rate (cM/Mb)
TTC28
HSCB
ZNRF3
MIR5739
CHEK2
XBP1
CCDC117
28.8
29
29.2
29.4
Position on chr22 (Mb)

### Small Cell Carcinoma

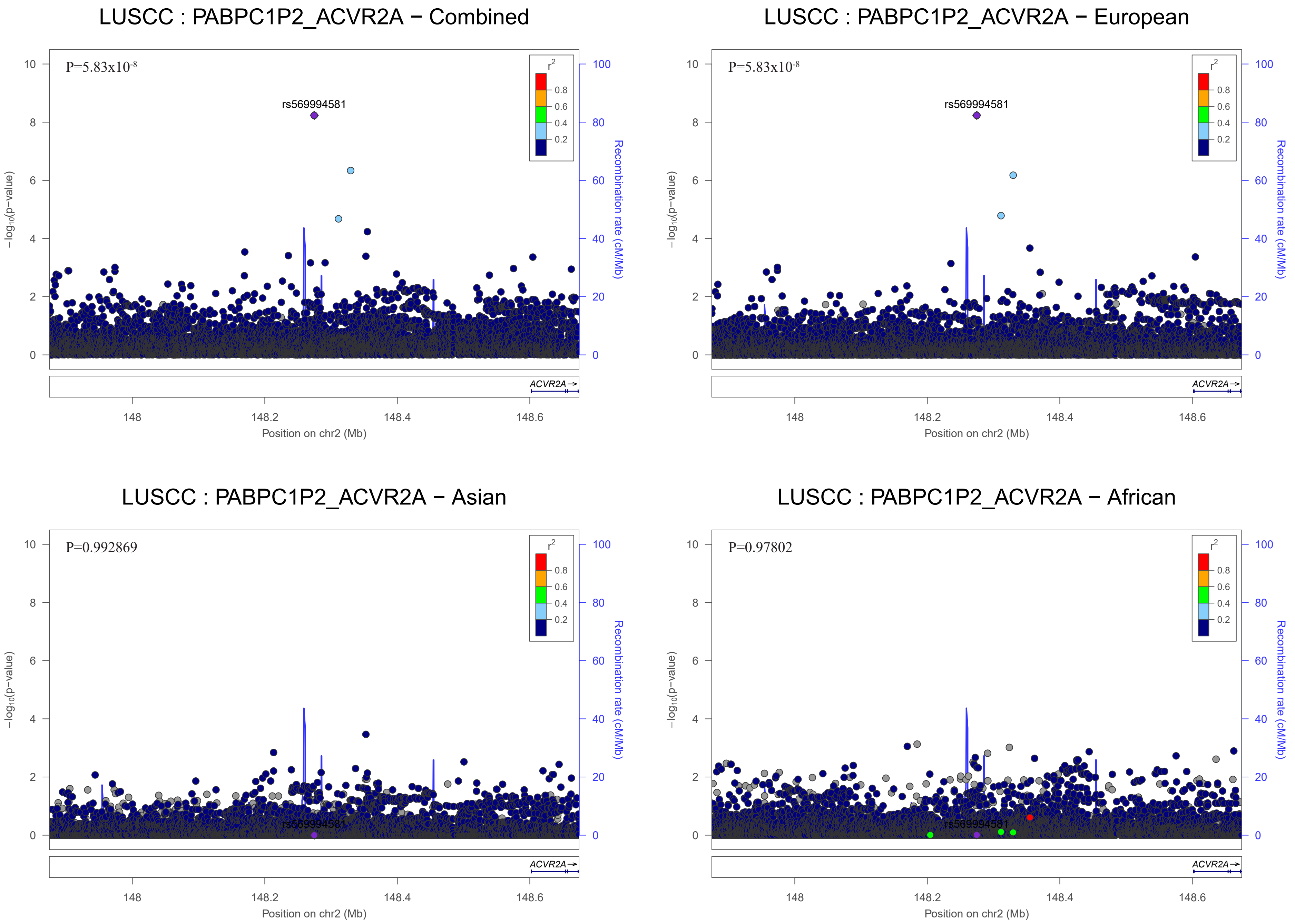
LUSCC : PABPC1P2_ACVR2A − Combined
P=5.83x10-8
rs569994581
LUSCC : PABPC1P2_ACVR2A − European
P=5.83x10-8
rs569994581
LUSCC : PABPC1P2_ACVR2A − Asian
P=0.992869
rs569994581
LUSCC : PABPC1P2_ACVR2A − African
P=0.97802
rs569994581
r2
0.8
0.6
0.4
0.2
−log10(p−value)
Recombination rate (cM/Mb)
ACVR2A
Position on chr2 (Mb)
148
148.2
148.4
148.6

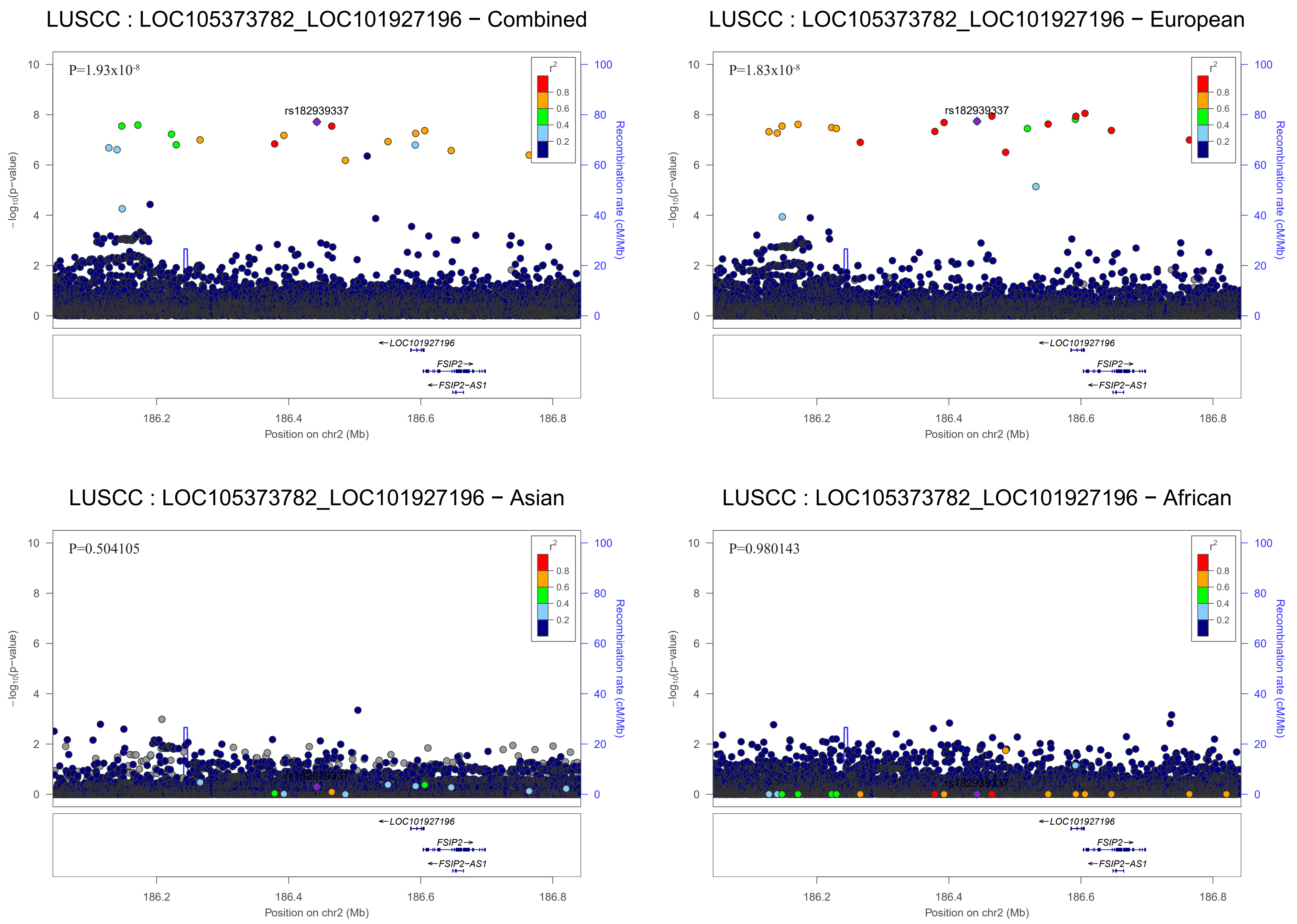
LUSCC : LOC105373782_LOC101927196 − Combined
P=1.93x10-8
rs182939337
LUSCC : LOC105373782_LOC101927196 − European
P=1.83x10-8
rs182939337
LUSCC : LOC105373782_LOC101927196 − Asian
P=0.504105
rs182939337
LUSCC : LOC105373782_LOC101927196 − African
P=0.980143
rs182939337
r2
0.8
0.6
0.4
0.2
−log10(p−value)
Recombination rate (cM/Mb)
LOC101927196
FSIP2
FSIP2−AS1
186.2
186.4
186.6
186.8
Position on chr2 (Mb)

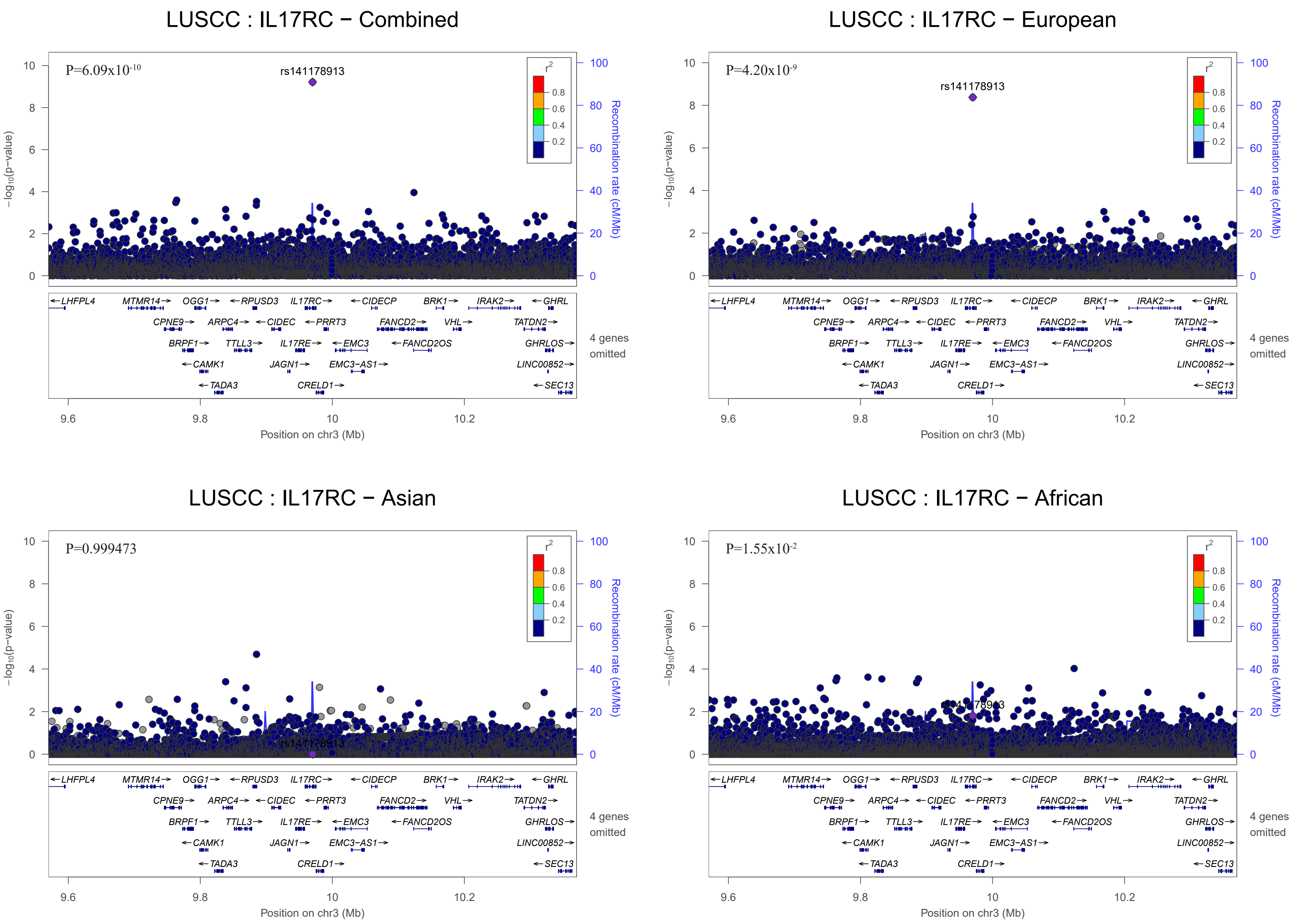
LUSCC : IL17RC − Combined
P=6.09x10-10
rs141178913
LUSCC : IL17RC − European
P=4.20x10-9
rs141178913
LUSCC : IL17RC − Asian
P=0.999473
rs141178913
LUSCC : IL17RC − African
P=1.55x10-2
rs141178913
−log10(p−value)
Recombination rate (cM/Mb)
r2
Position on chr3 (Mb)
4 genes omitted
LHFPL4
MTMR14
OGG1
RPUSD3
IL17RC
CIDECP
BRK1
IRAK2
GHRL
CPNE9
ARPC4
CIDEC
PRRT3
FANCD2
VHL
TATDN2
BRPF1
TTLL3
IL17RE
EMC3
FANCD2OS
GHRLOS
CAMK1
JAGN1
EMC3−AS1
LINC00852
TADA3
CRELD1
SEC13

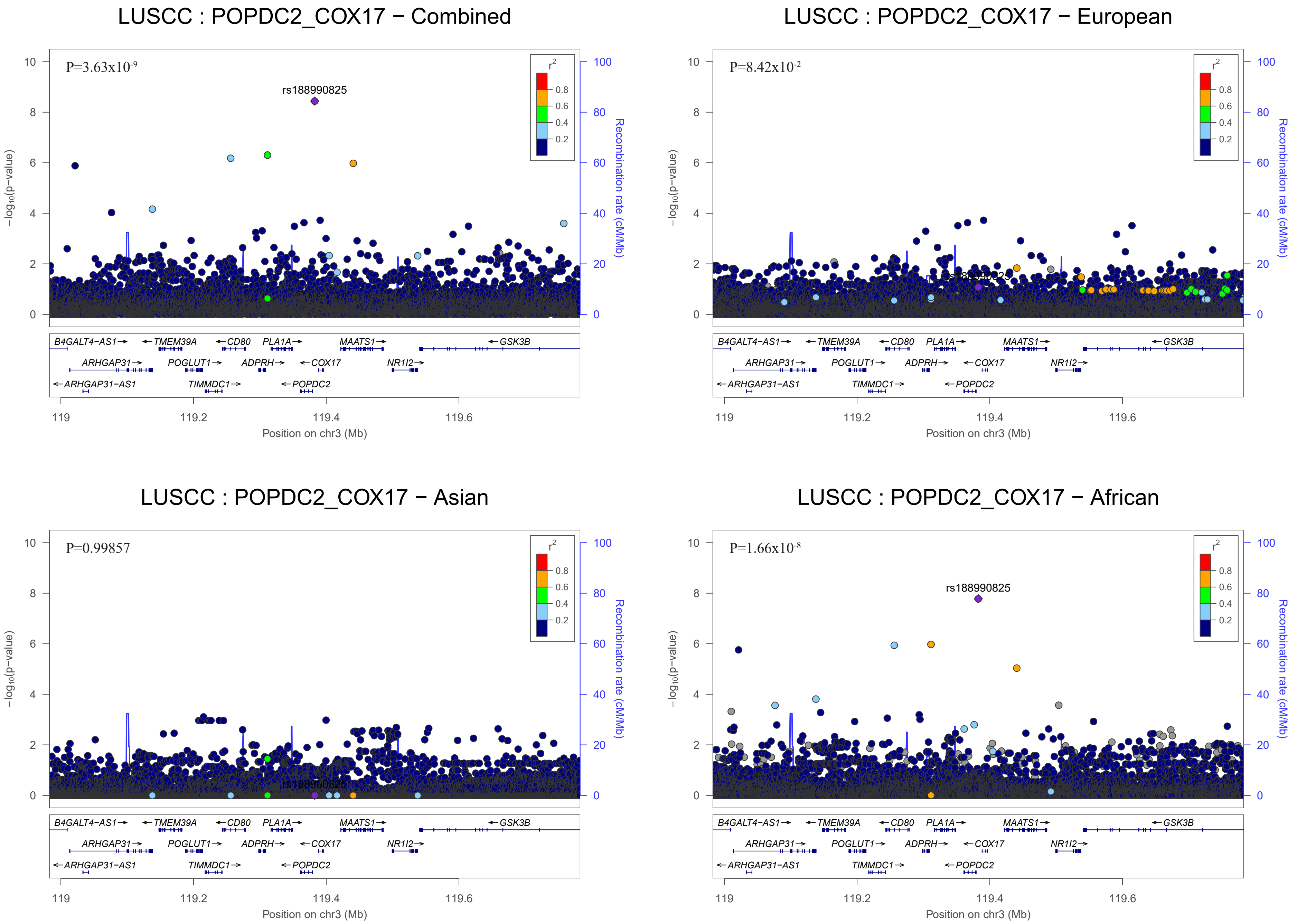
LUSCC : POPDC2_COX17 − Combined
P=3.63x10-9
rs188990825
LUSCC : POPDC2_COX17 − European
P=8.42x10-2
LUSCC : POPDC2_COX17 − Asian
P=0.99857
LUSCC : POPDC2_COX17 − African
P=1.66x10-8
rs188990825
r2
0.8
0.6
0.4
0.2
−log10(p−value)
Recombination rate (cM/Mb)
Position on chr3 (Mb)
119
119.2
119.4
119.6
B4GALT4−AS1→
←TMEM39A
←CD80
PLA1A→
MAATS1→
←GSK3B
ARHGAP31→
POGLUT1→
ADPRH→
←COX17
NR1I2→
←ARHGAP31−AS1
TIMMDC1→
←POPDC2

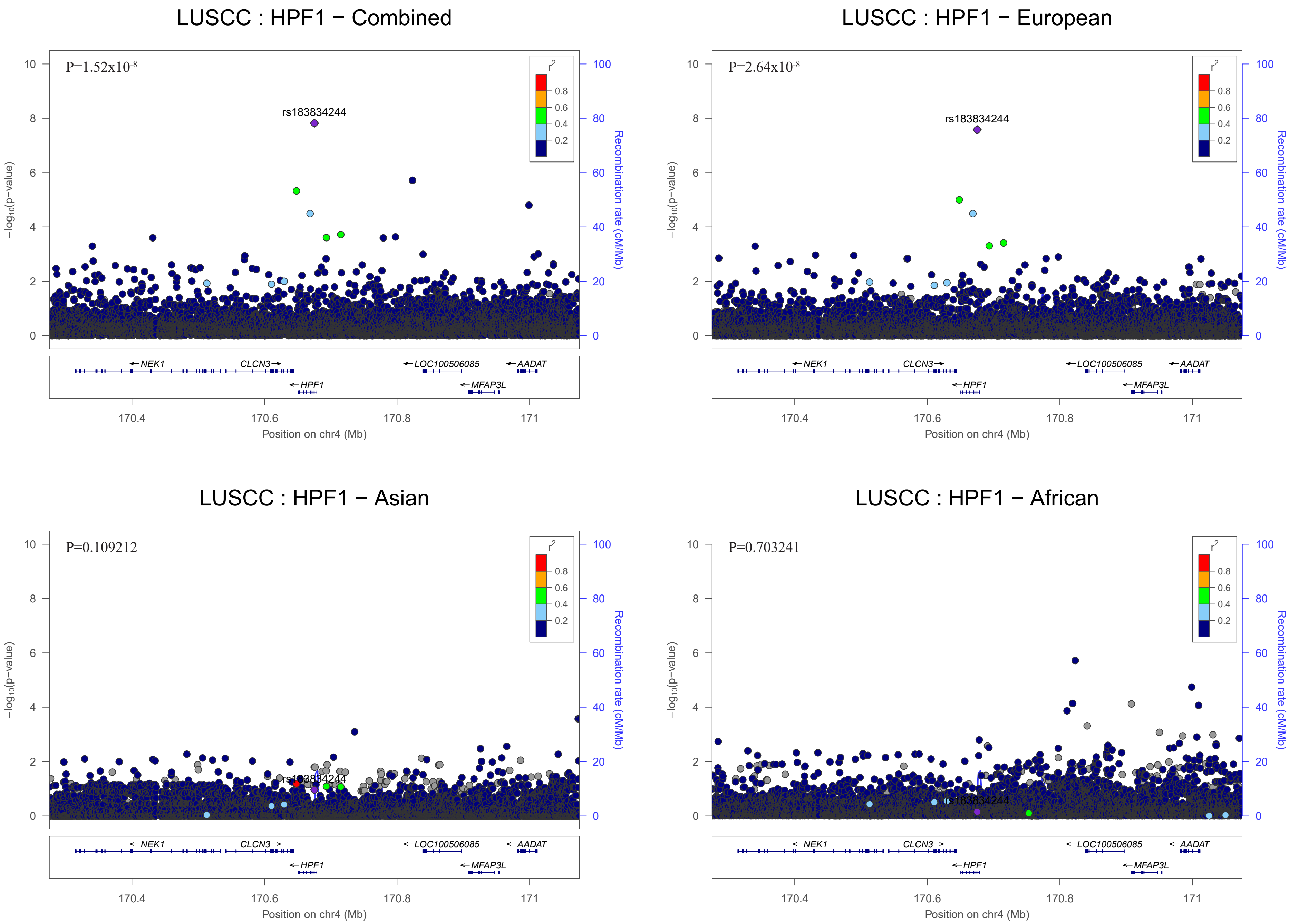
LUSCC : HPF1 − Combined
P=1.52x10-8
rs183834244
LUSCC : HPF1 − European
P=2.64x10-8
rs183834244
LUSCC : HPF1 − Asian
P=0.109212
rs183834244
LUSCC : HPF1 − African
P=0.703241
rs183834244
r2
0.8
0.6
0.4
0.2
−log10(p−value)
Recombination rate (cM/Mb)
NEK1
CLCN3
HPF1
LOC100506085
MFAP3L
AADAT
170.4
170.6
170.8
171
Position on chr4 (Mb)

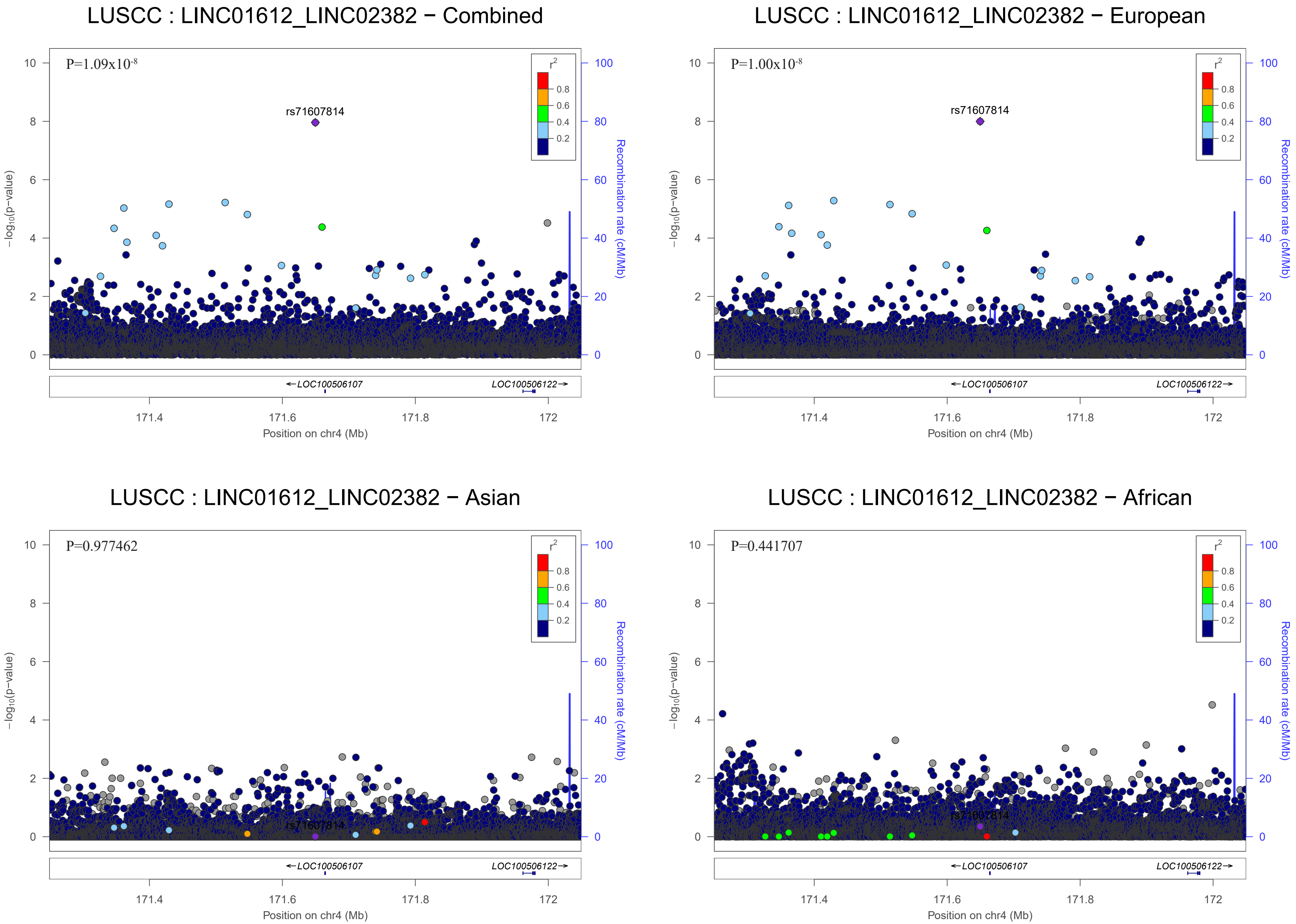

LUSCC : LINC01612_LINC02382 − Combined
P=1.09x10-8
rs71607814
LUSCC : LINC01612_LINC02382 − European
P=1.00x10-8
rs71607814
LUSCC : LINC01612_LINC02382 − Asian
P=0.977462
rs71607814
LUSCC : LINC01612_LINC02382 − African
P=0.441707
rs71607814
r2
0.8
0.6
0.4
0.2
−log10(p−value)
Recombination rate (cM/Mb)
LOC100506107
LOC100506122
171.4
171.6
171.8
172
Position on chr4 (Mb)

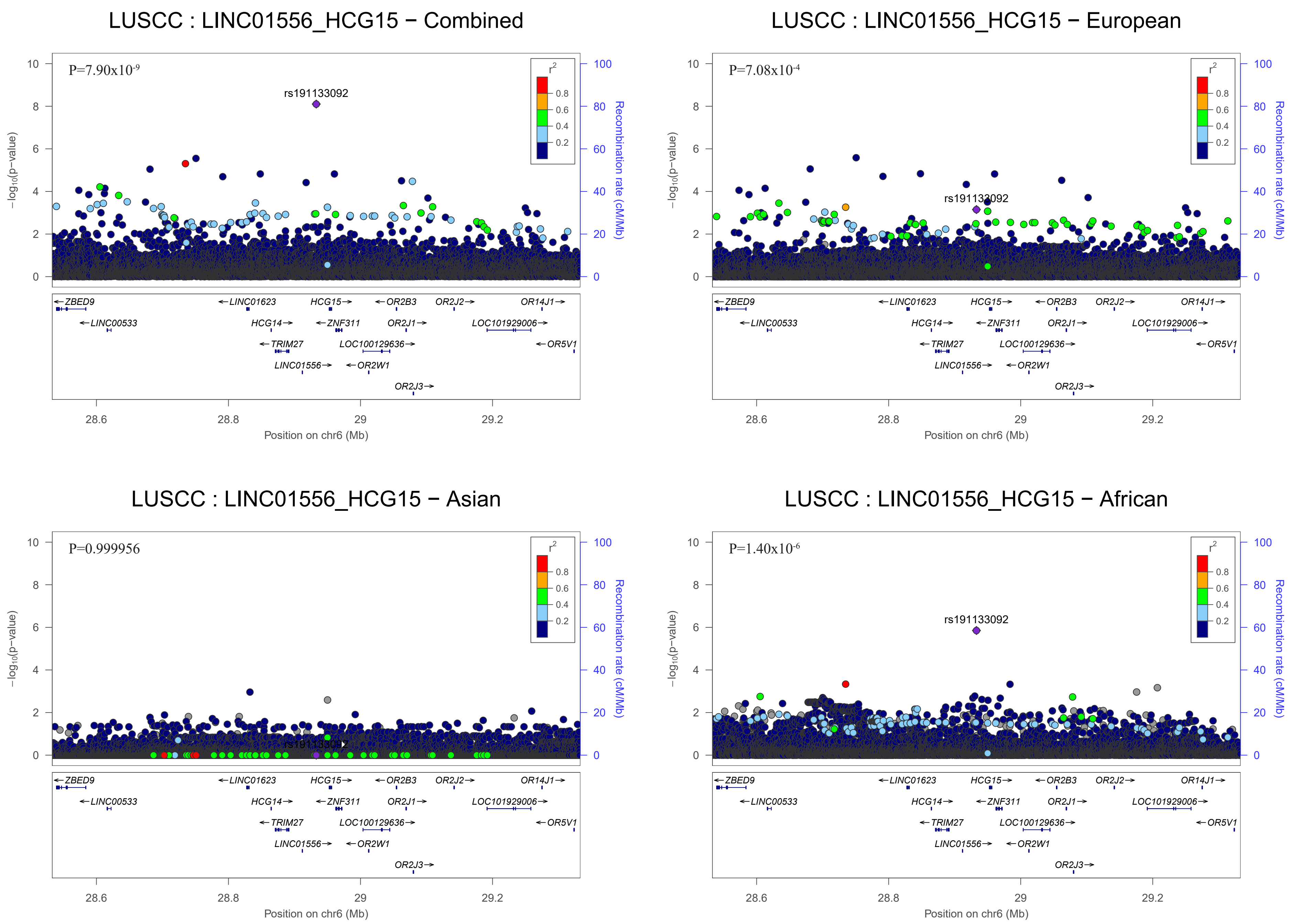
LUSCC : LINC01556_HCG15 − Combined
P=7.90x10-9
rs191133092
LUSCC : LINC01556_HCG15 − European
P=7.08x10-4
rs191133092
LUSCC : LINC01556_HCG15 − Asian
P=0.999956
rs191133092
LUSCC : LINC01556_HCG15 − African
P=1.40x10-6
rs191133092
r2
0.8
0.6
0.4
0.2
−log10(p−value)
Recombination rate (cM/Mb)
Position on chr6 (Mb)
28.6
28.8
29
29.2
ZBED9
LINC00533
LINC01623
HCG14
TRIM27
LINC01556
HCG15
ZNF311
OR2W1
OR2B3
OR2J1
LOC100129636
OR2J3
OR2J2
LOC101929006
OR14J1
OR5V1

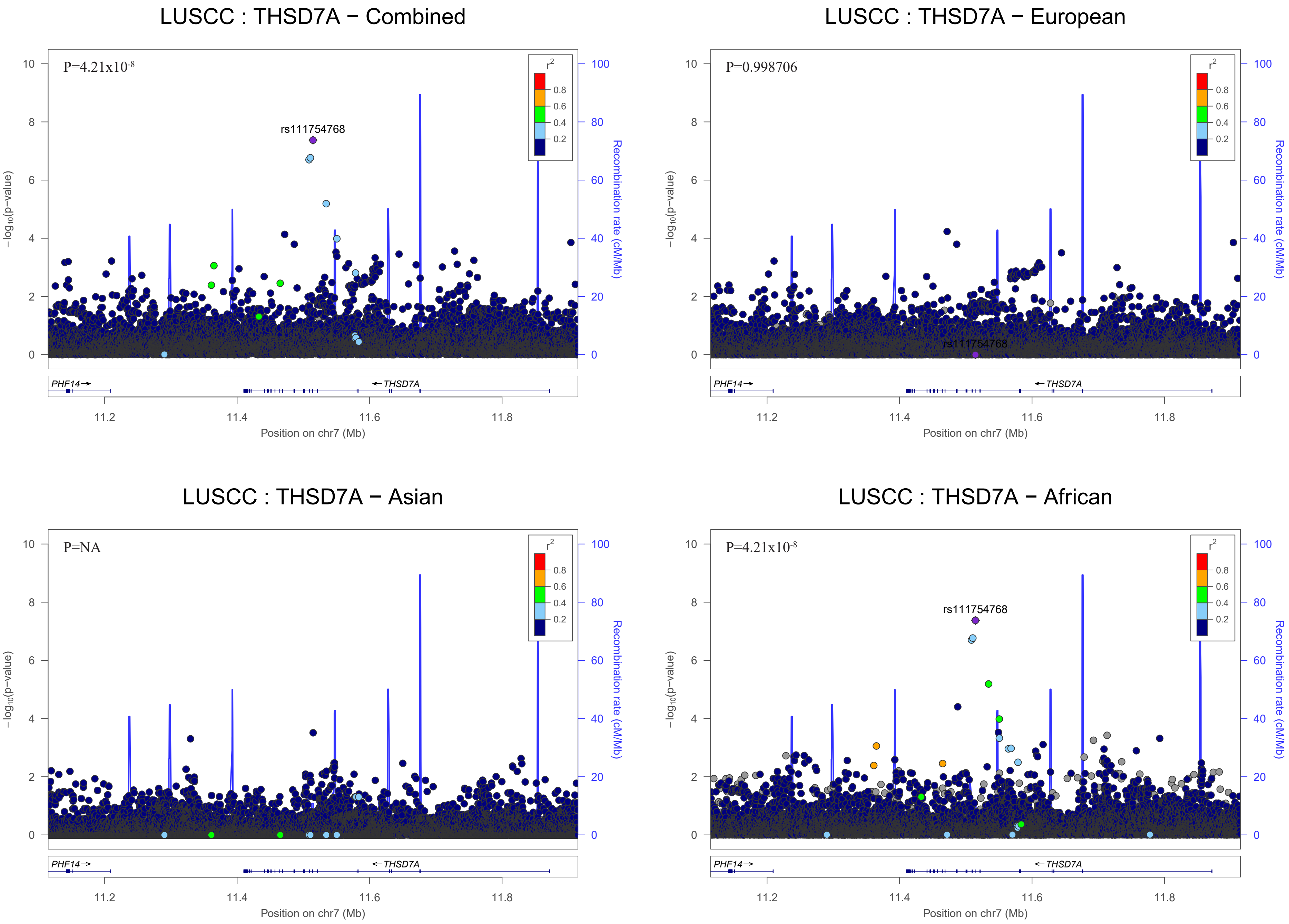

LUSCC : THSD7A − Combined
P=4.21x10-8
rs111754768
LUSCC : THSD7A − European
P=0.998706
rs111754768
LUSCC : THSD7A − Asian
P=NA
LUSCC : THSD7A − African
P=4.21x10-8
rs111754768
r2
0.8
0.6
0.4
0.2
−log10(p−value)
Recombination rate (cM/Mb)
PHF14
THSD7A
11.2
11.4
11.6
11.8
Position on chr7 (Mb)

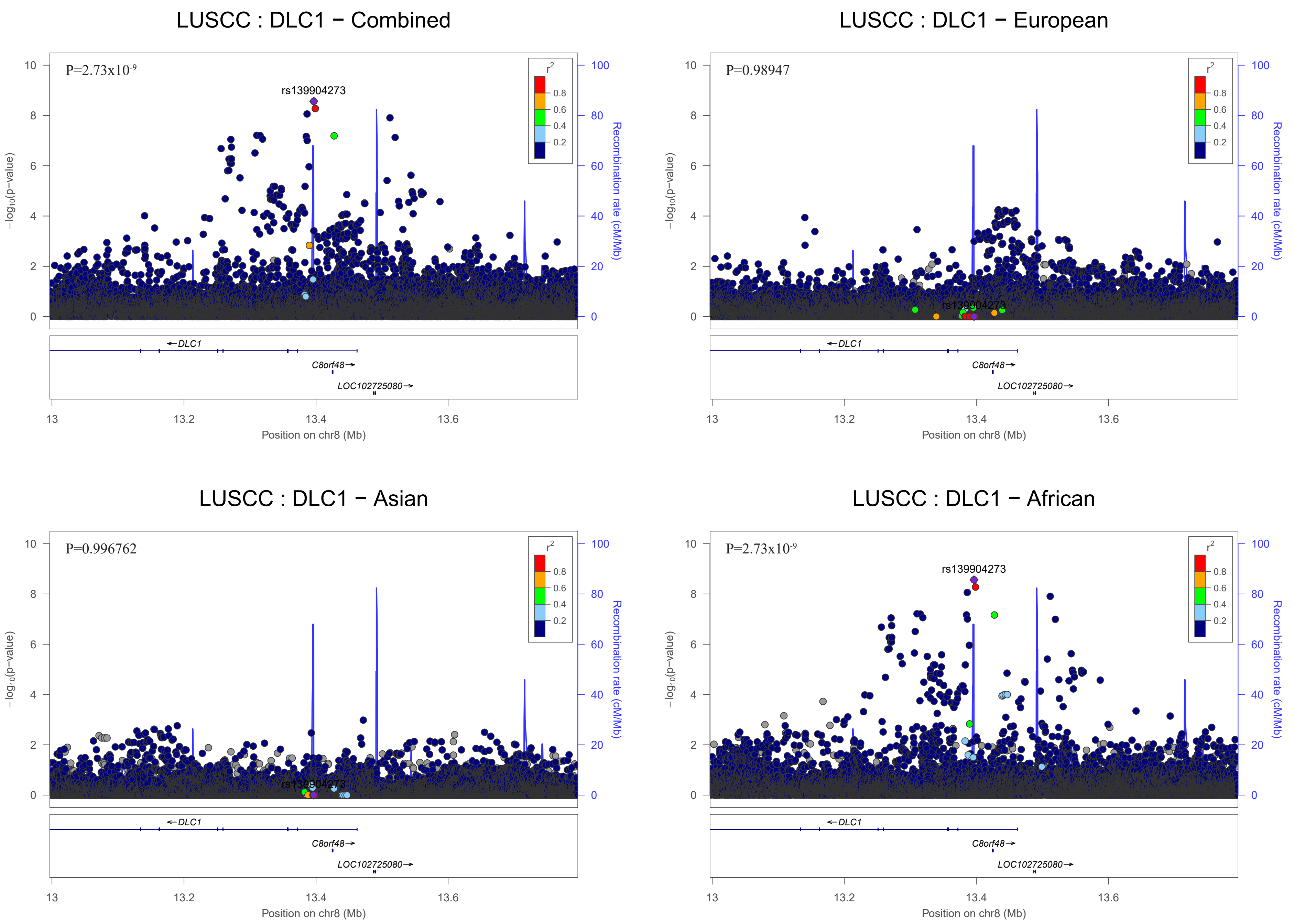
LUSCC : DLC1 − Combined
P=2.73x10-9
rs139904273
LUSCC : DLC1 − European
P=0.98947
rs139904273
LUSCC : DLC1 − Asian
P=0.996762
rs139904273
LUSCC : DLC1 − African
P=2.73x10-9
rs139904273
r2
0.8
0.6
0.4
0.2
−log10(p−value)
Recombination rate (cM/Mb)
←DLC1
C8orf48→
LOC102725080→
13
13.2
13.4
13.6
Position on chr8 (Mb)

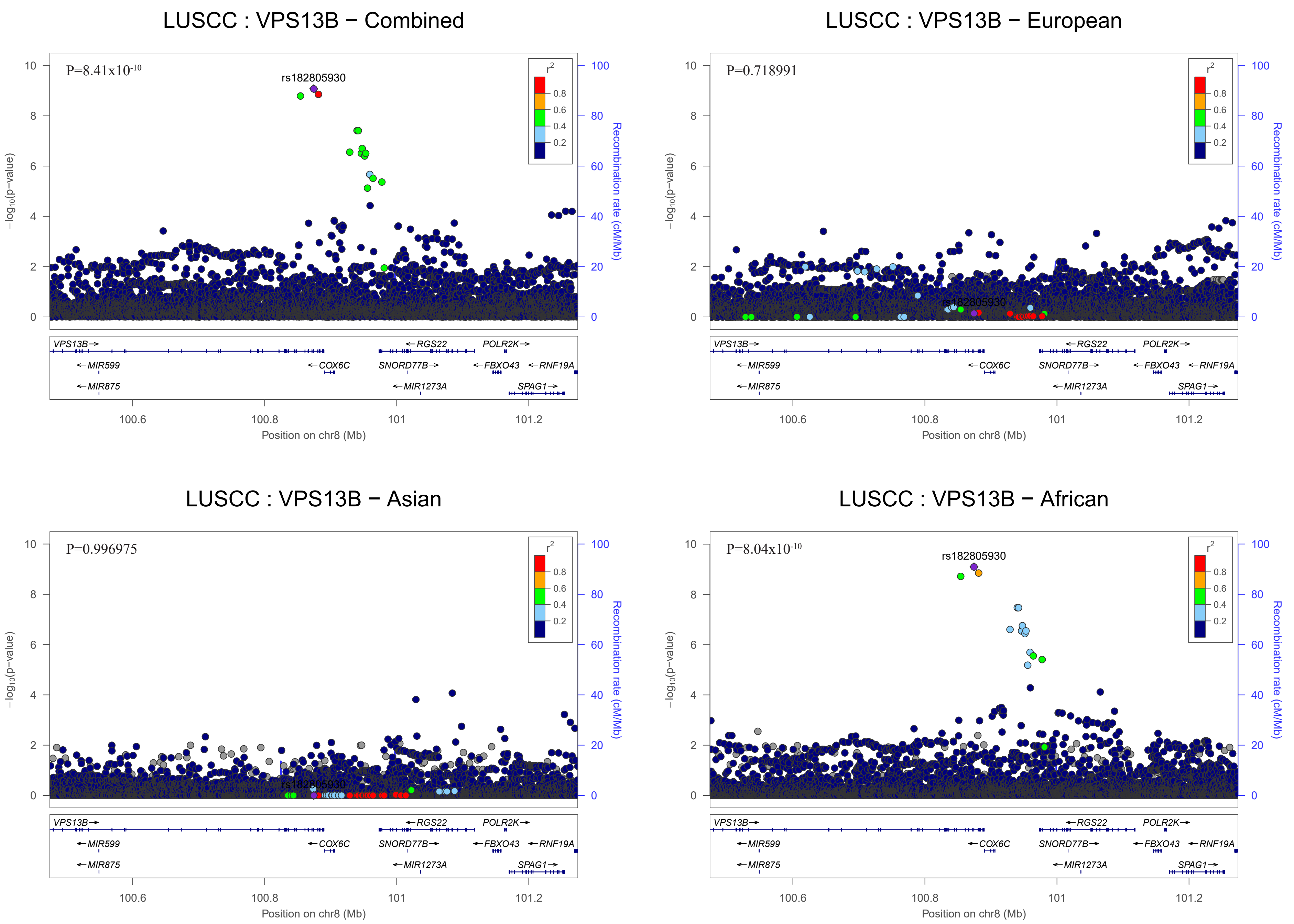

LUSCC : VPS13B − Combined
P=8.41x10-10
rs182805930
LUSCC : VPS13B − European
P=0.718991
rs182805930
LUSCC : VPS13B − Asian
P=0.996975
rs182805930
LUSCC : VPS13B − African
P=8.04x10-10
rs182805930
r2
0.8
0.6
0.4
0.2
−log10(p−value)
Recombination rate (cM/Mb)
VPS13B
MIR599
MIR875
COX6C
RGS22
SNORD77B
MIR1273A
POLR2K
FBXO43
RNF19A
SPAG1
100.6
100.8
101
101.2
Position on chr8 (Mb)

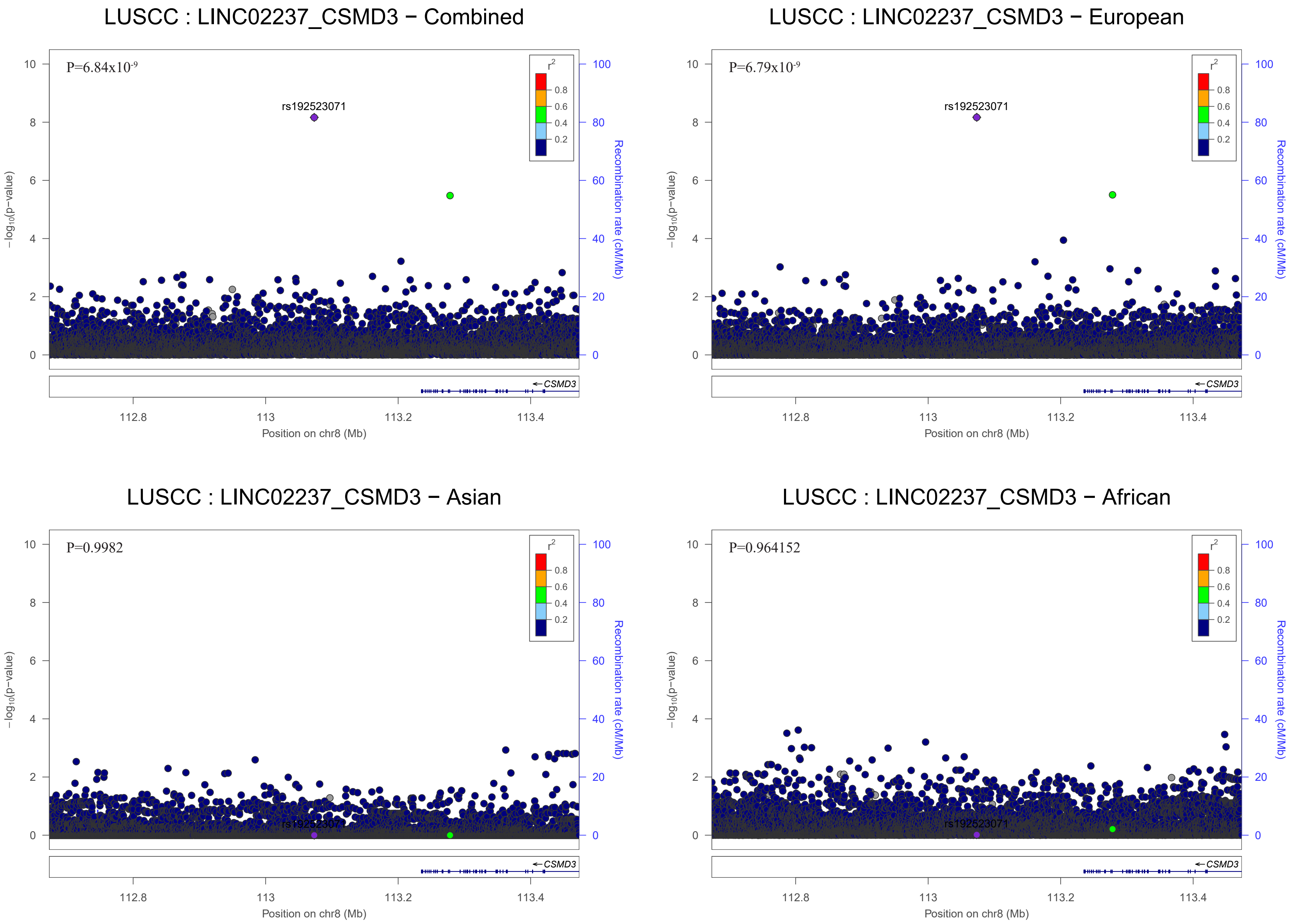
LUSCC : LINC02237_CSMD3 − Combined
P=6.84x10-9
rs192523071
LUSCC : LINC02237_CSMD3 − European
P=6.79x10-9
rs192523071
LUSCC : LINC02237_CSMD3 − Asian
P=0.9982
rs192523071
LUSCC : LINC02237_CSMD3 − African
P=0.964152
rs192523071
r2
0.8
0.6
0.4
0.2
CSMD3
−log10(p-value)
Recombination rate (cM/Mb)
Position on chr8 (Mb)
112.8
113
113.2
113.4

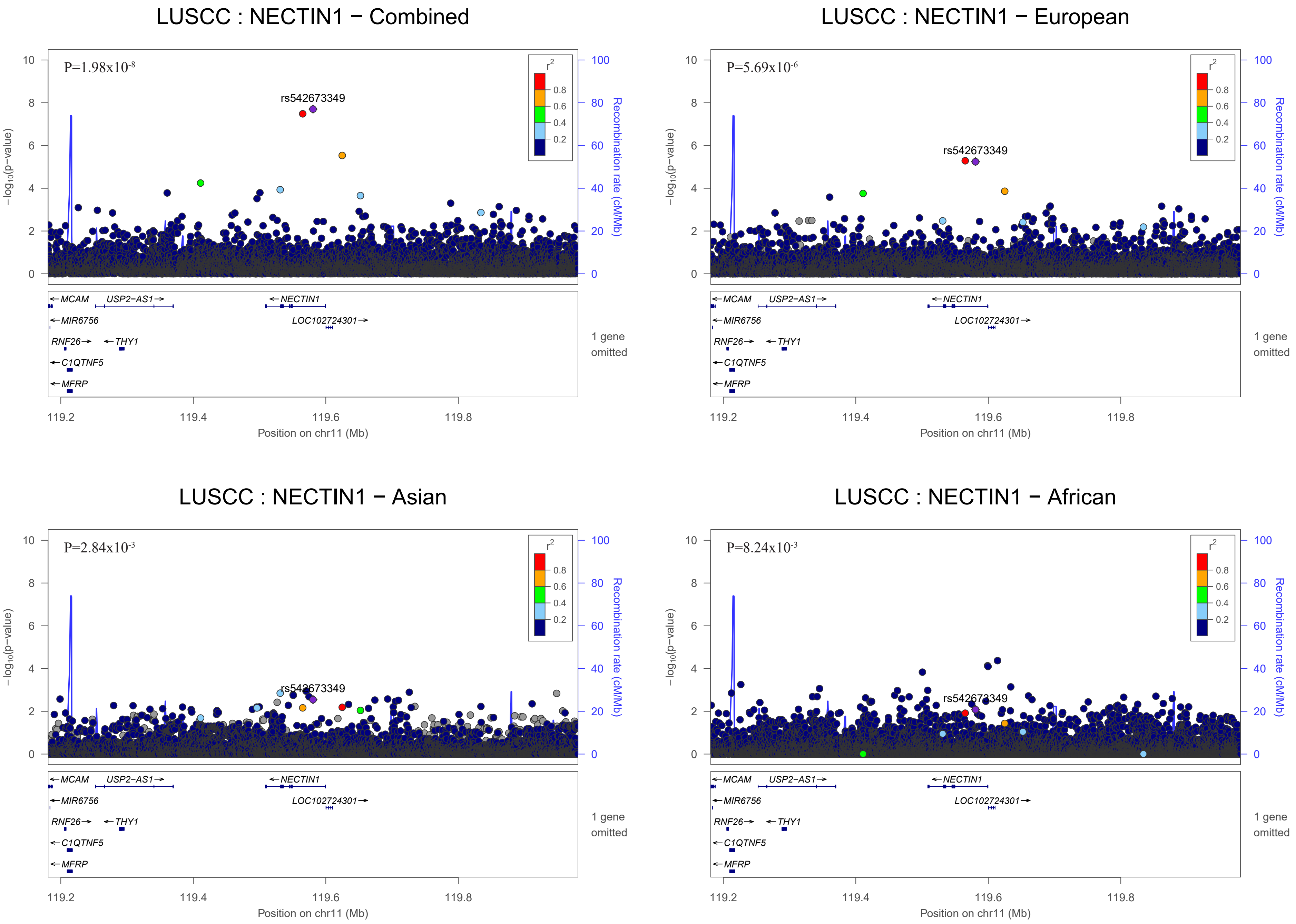
LUSCC : NECTIN1 − Combined
P=1.98x10-8
rs542673349
LUSCC : NECTIN1 − European
P=5.69x10-6
rs542673349
LUSCC : NECTIN1 − Asian
P=2.84x10-3
rs542673349
LUSCC : NECTIN1 − African
P=8.24x10-3
rs542673349
r2
0.8
0.6
0.4
0.2
−log10(p−value)
Recombination rate (cM/Mb)
MCAM
USP2−AS1
NECTIN1
MIR6756
LOC102724301
RNF26
THY1
C1QTNF5
MFRP
1 gene omitted
119.2
119.4
119.6
119.8
Position on chr11 (Mb)

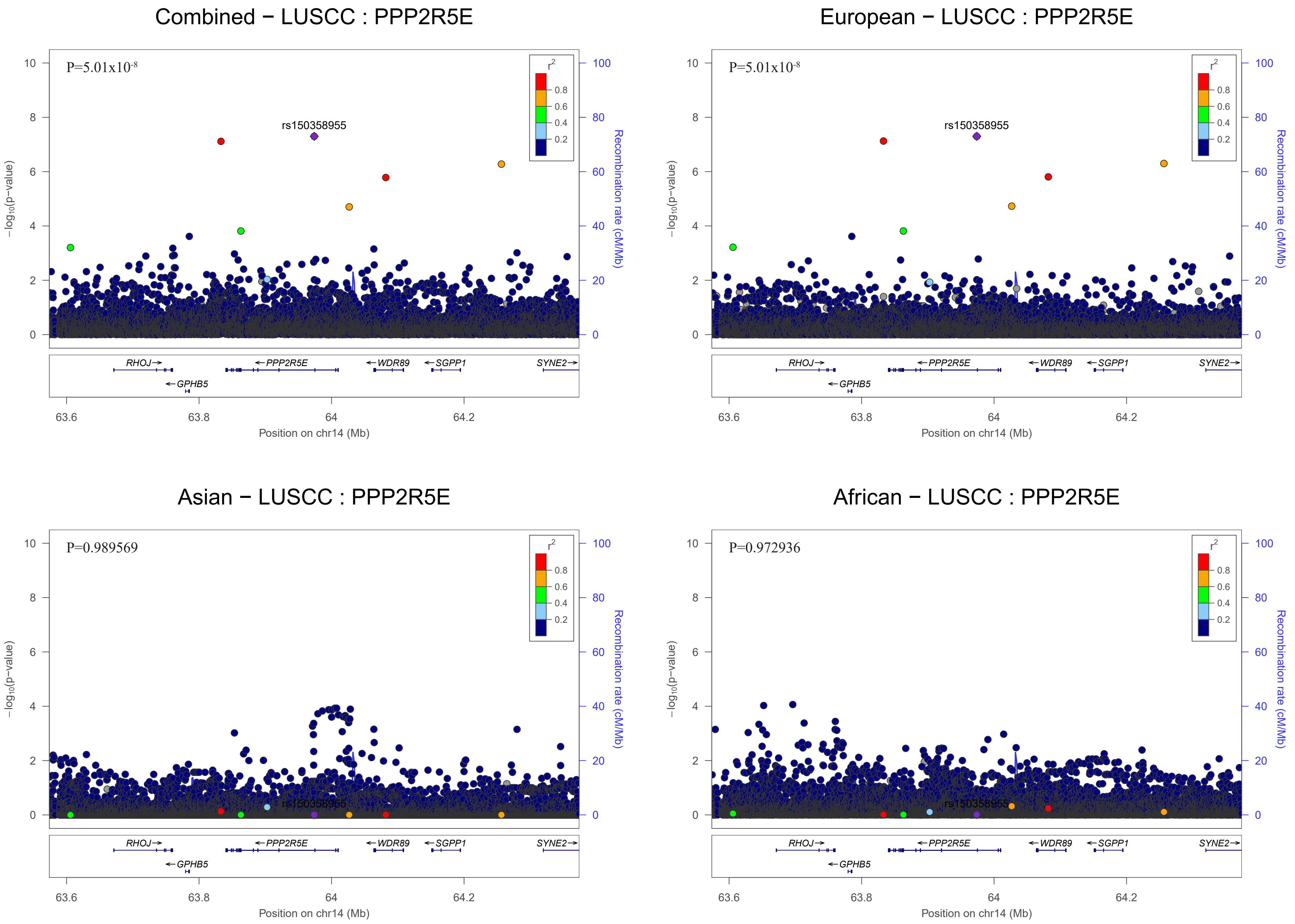

Combined − LUSCC : PPP2R5E
P=5.01x10-8
rs150358955
European − LUSCC : PPP2R5E
P=5.01x10-8
rs150358955
Asian − LUSCC : PPP2R5E
P=0.989569
African − LUSCC : PPP2R5E
P=0.972936
r2
0.8
0.6
0.4
0.2
−log10(p−value)
Recombination rate (cM/Mb)
RHOJ
GPHB5
PPP2R5E
WDR89
SGPP1
SYNE2
63.6
63.8
64
64.2
Position on chr14 (Mb)

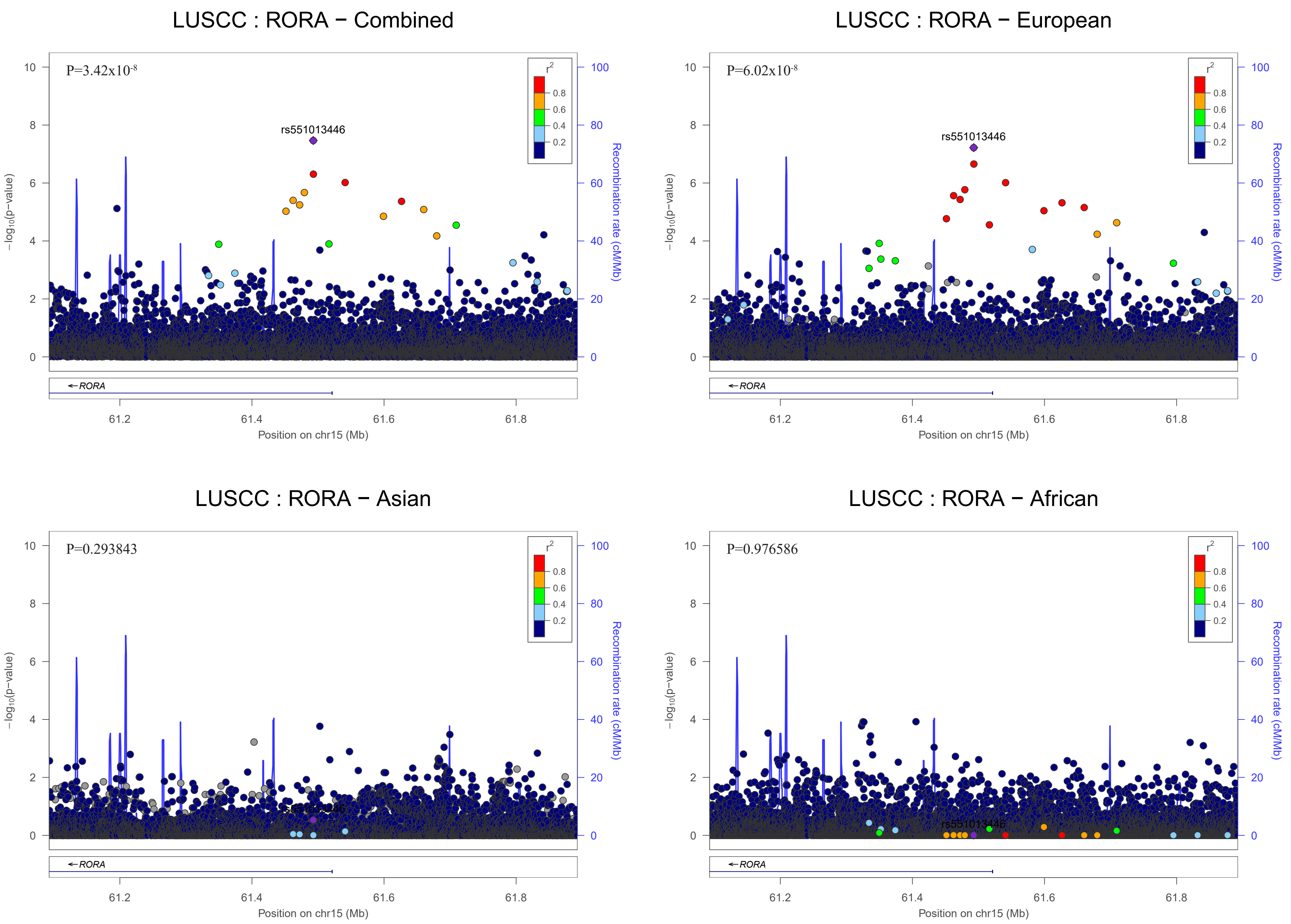
LUSCC : RORA − Combined
P=3.42x10-8
rs551013446
LUSCC : RORA − European
P=6.02x10-8
rs551013446
LUSCC : RORA − Asian
P=0.293843
LUSCC : RORA − African
P=0.976586
rs551013446
r2
−log10(p−value)
Recombination rate (cM/Mb)
RORA
Position on chr15 (Mb)

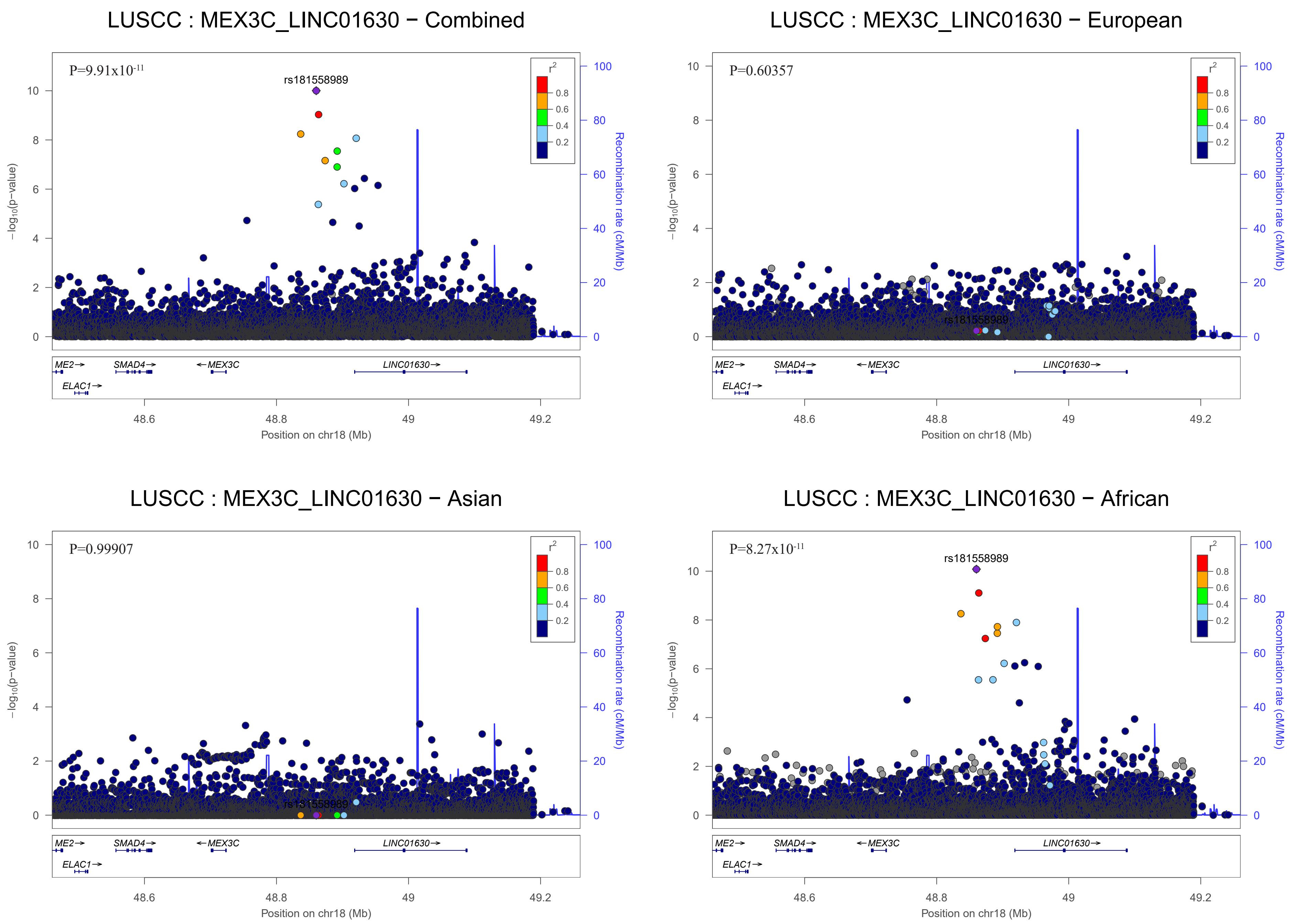
LUSCC : MEX3C_LINC01630 − Combined
P=9.91x10-11
rs181558989
LUSCC : MEX3C_LINC01630 − European
P=0.60357
rs181558989
LUSCC : MEX3C_LINC01630 − Asian
P=0.99907
rs181558989
LUSCC : MEX3C_LINC01630 − African
P=8.27x10-11
rs181558989
r2
0.8
0.6
0.4
0.2
−log10(p−value)
Recombination rate (cM/Mb)
ME2→
SMAD4→
←MEX3C
LINC01630→
ELAC1→
48.6
48.8
49
49.2
Position on chr18 (Mb)

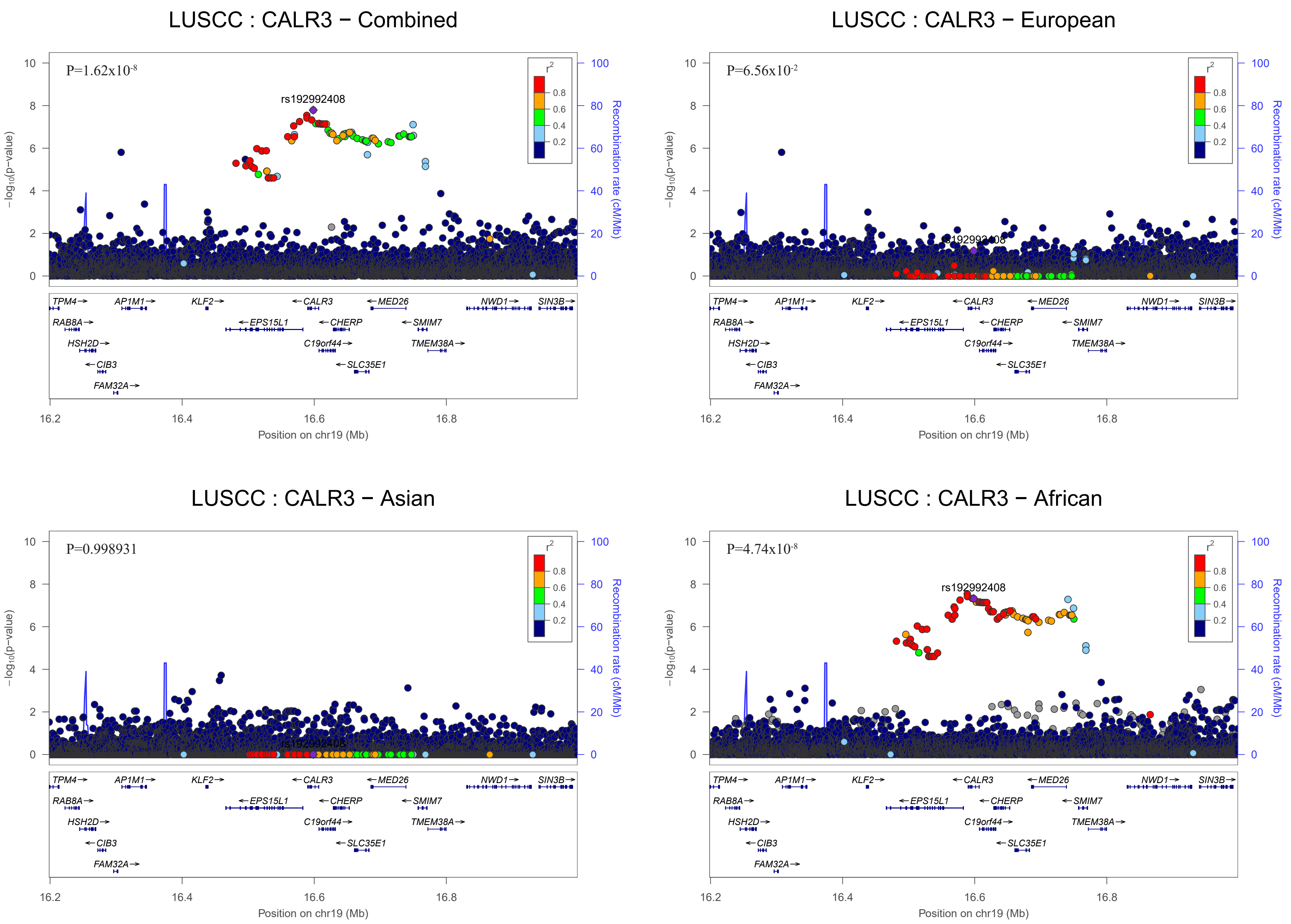
LUSCC : CALR3 − Combined
P=1.62x10-8
rs192992408
LUSCC : CALR3 − European
P=6.56x10-2
LUSCC : CALR3 − Asian
P=0.998931
LUSCC : CALR3 − African
P=4.74x10-8
r2
0.8
0.6
0.4
0.2
−log10(p−value)
Recombination rate (cM/Mb)
Position on chr19 (Mb)
16.2
16.4
16.6
16.8
TPM4
AP1M1
KLF2
CALR3
MED26
NWD1
SIN3B
RAB8A
EPS15L1
CHERP
SMIM7
HSH2D
C19orf44
TMEM38A
CIB3
SLC35E1
FAM32A

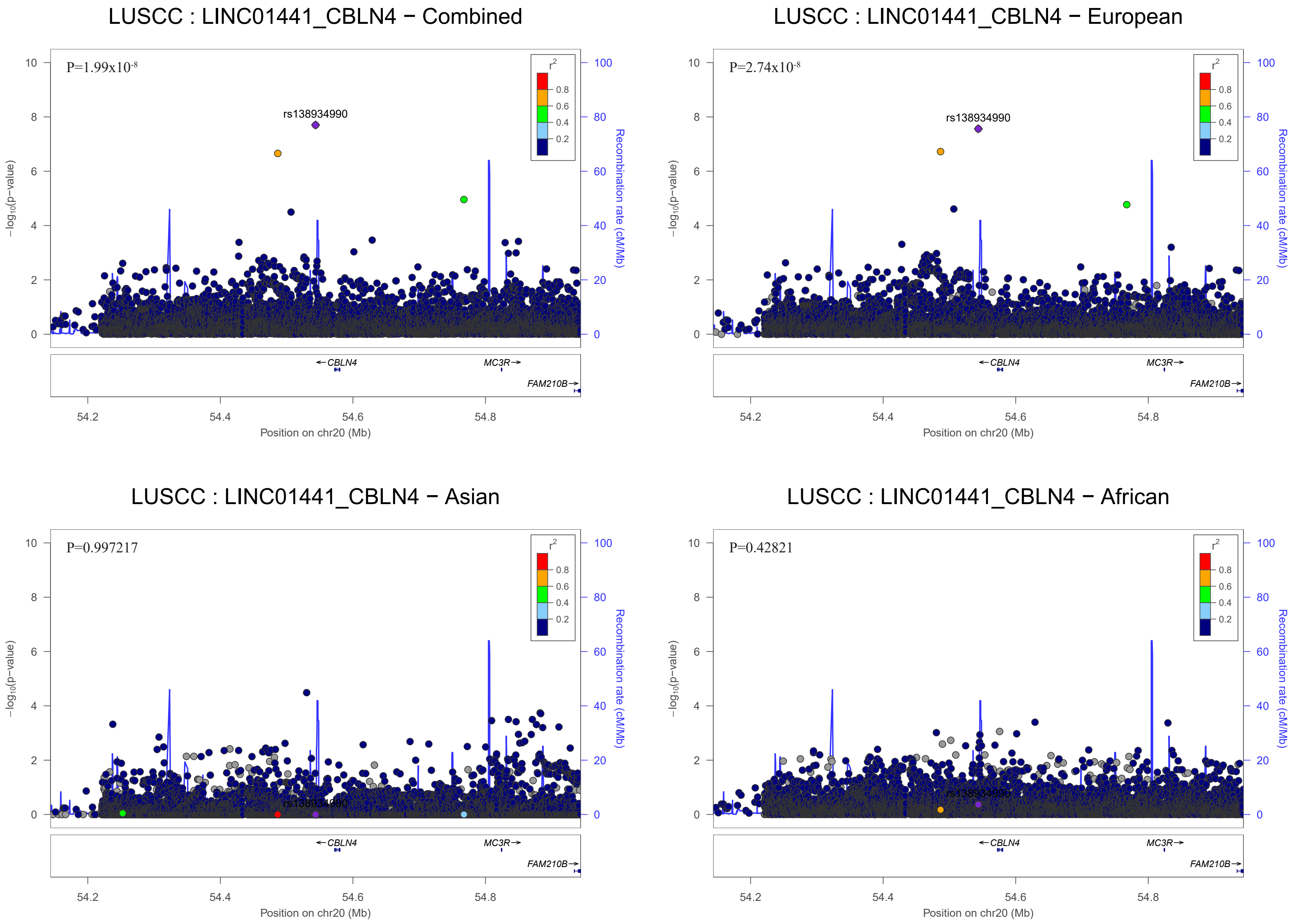
LUSCC : LINC01441_CBLN4 − Combined
P=1.99x10-8
rs138934990
LUSCC : LINC01441_CBLN4 − European
P=2.74x10-8
rs138934990
LUSCC : LINC01441_CBLN4 − Asian
P=0.997217
rs138934990
LUSCC : LINC01441_CBLN4 − African
P=0.42821
rs138934990
r2
0.8
0.6
0.4
0.2
−log10(p−value)
Recombination rate (cM/Mb)
CBLN4
MC3R
FAM210B
Position on chr20 (Mb)
54.2
54.4
54.6
54.8

**Title:** Trans-ethnic genome-wide meta-analysis of 35,732 cases and 34,424 controls identifies novel genomic cross-ancestry loci contributing to lung cancer susceptibility

### Supplementary Figure 3
### Genomic Regional Plots from Conditional Analysis

### Model : Analyses by Race and then meta-analysis

| | SNP | chr:pos | Conditioning On | Conditional Pvalue | GENE | Pvalue before conditioning |
|---|---|---|---|---|---|---|
| step0 | rs2853677 | 5:1287194:G:A | No condition | | TERT;Known variant | 2.91E-50 |
| step1 | rs31487 | 5:1341101:G:C | rs2853677 | 9.87E-31 | CLPTM1L | 1.53E-41 |
| step2 | rs13167280 | 5:1280477:G:A | rs2853677;rs3148 | 8.82E-11 | TERT | 5.17E-23 |
| step3 | rs7705526 | 5:1285974:C:A | rs2853677;rs3148;rs13167280 | 1.23E-05 | TERT;Known variant | 4.83E-38 |

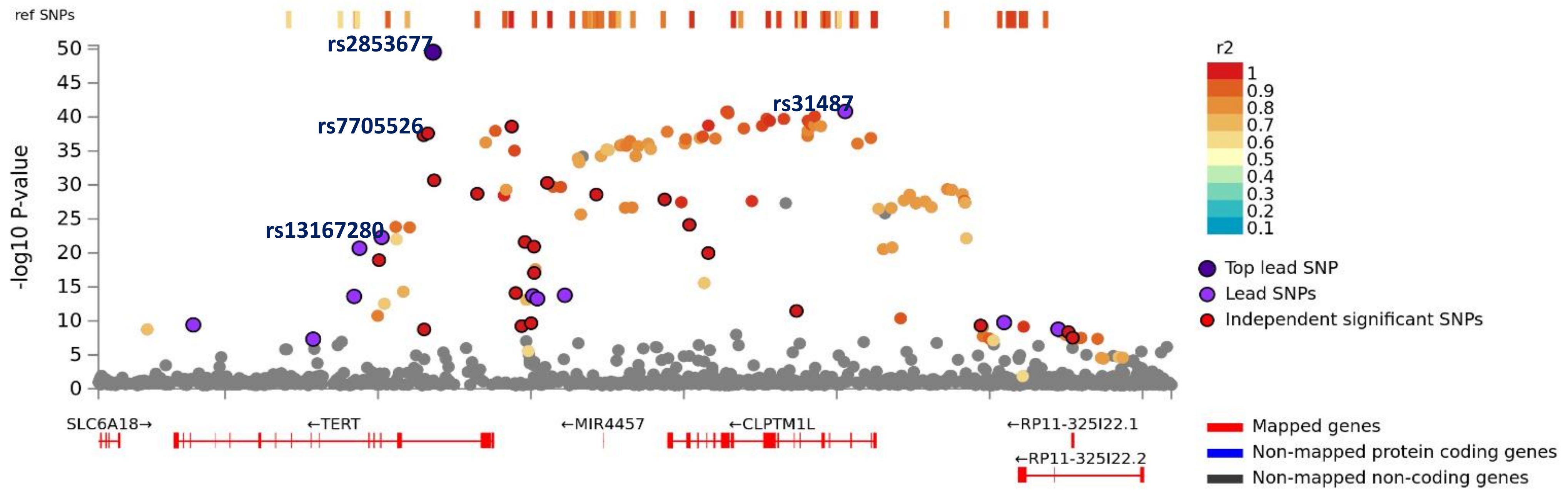

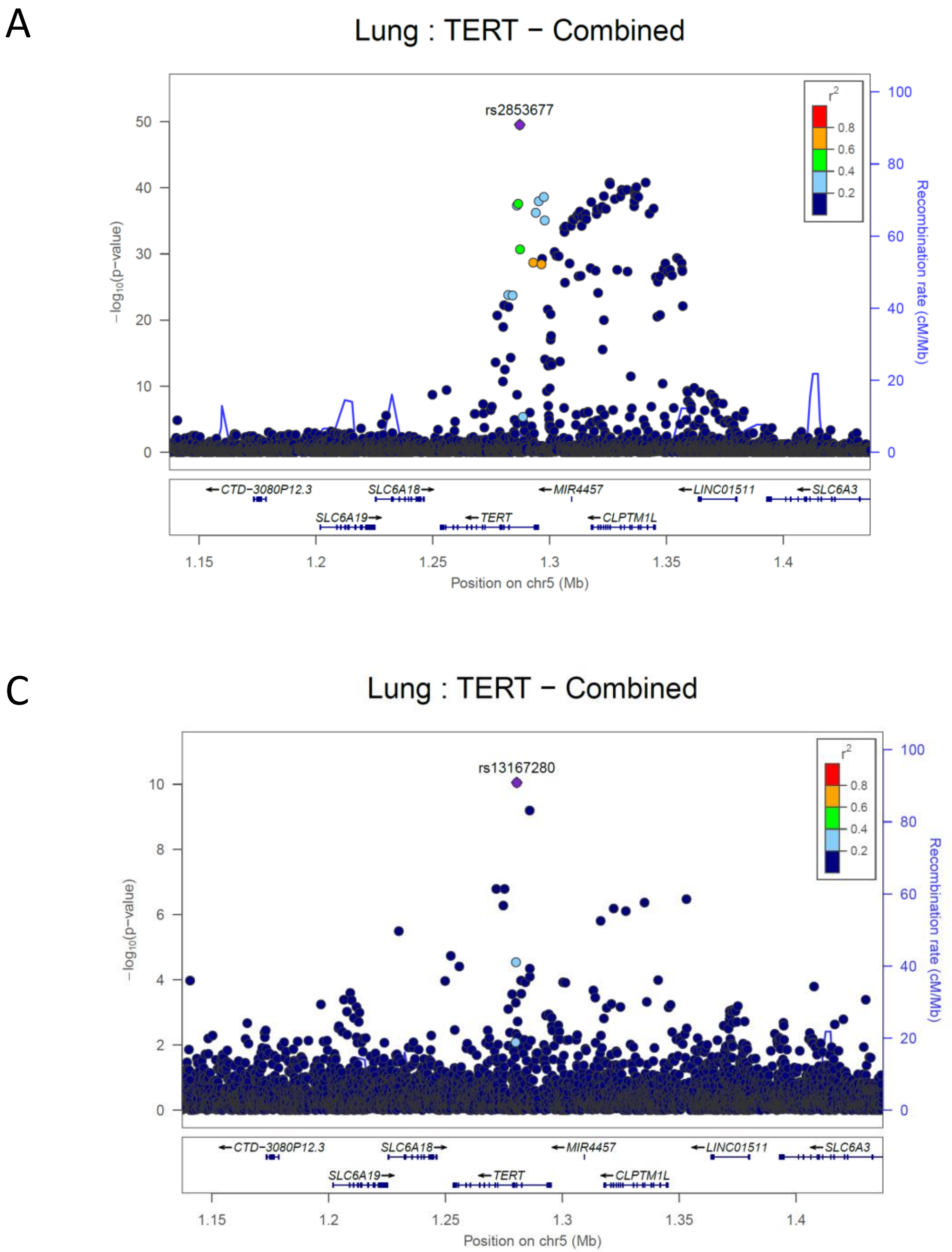


**A : Before Conditioning**

**B : After Conditioned rs2853677**

**C : After Conditioned rs2853677, rs31487**

**Note : Conditioned 1Mb loci**

A

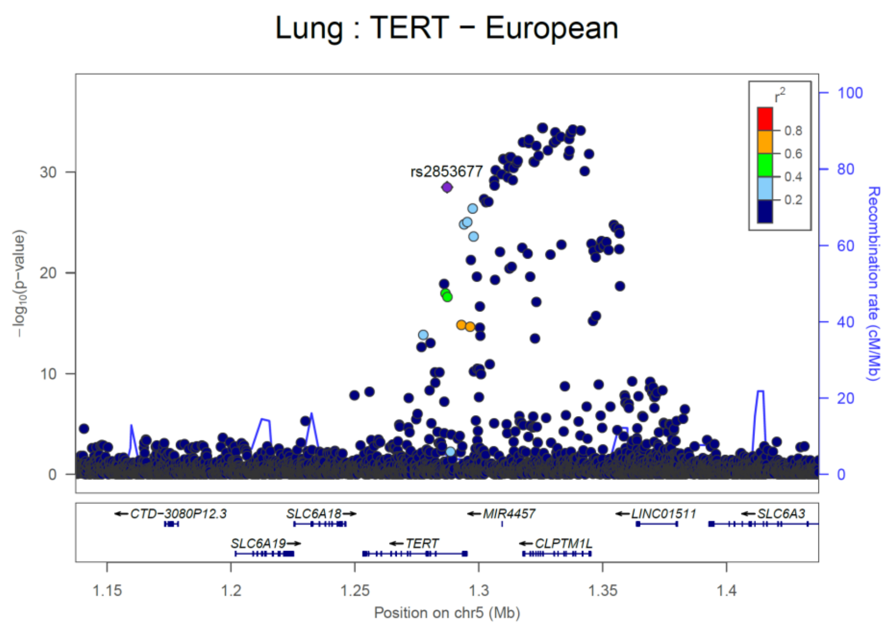


B

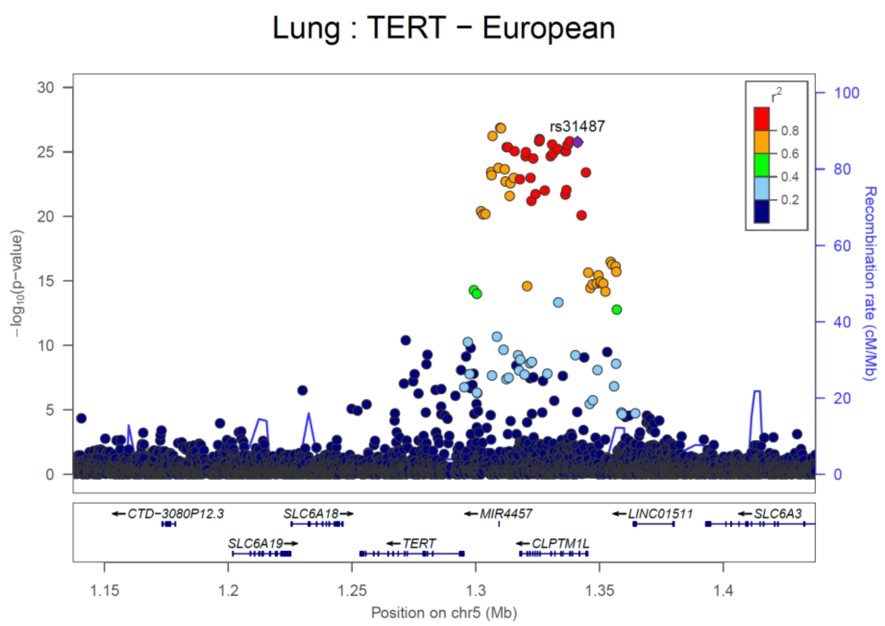


C

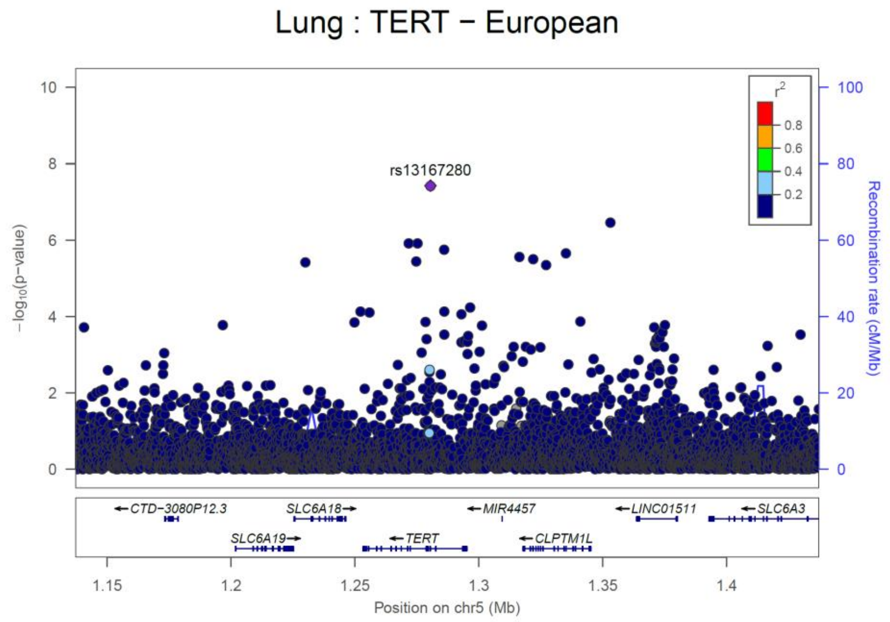


**A : Before Conditioning**
**B : After Conditioned rs2853677**
**C : After Conditioned rs2853677, rs31487**
**Note : Conditioned 1Mb loci**

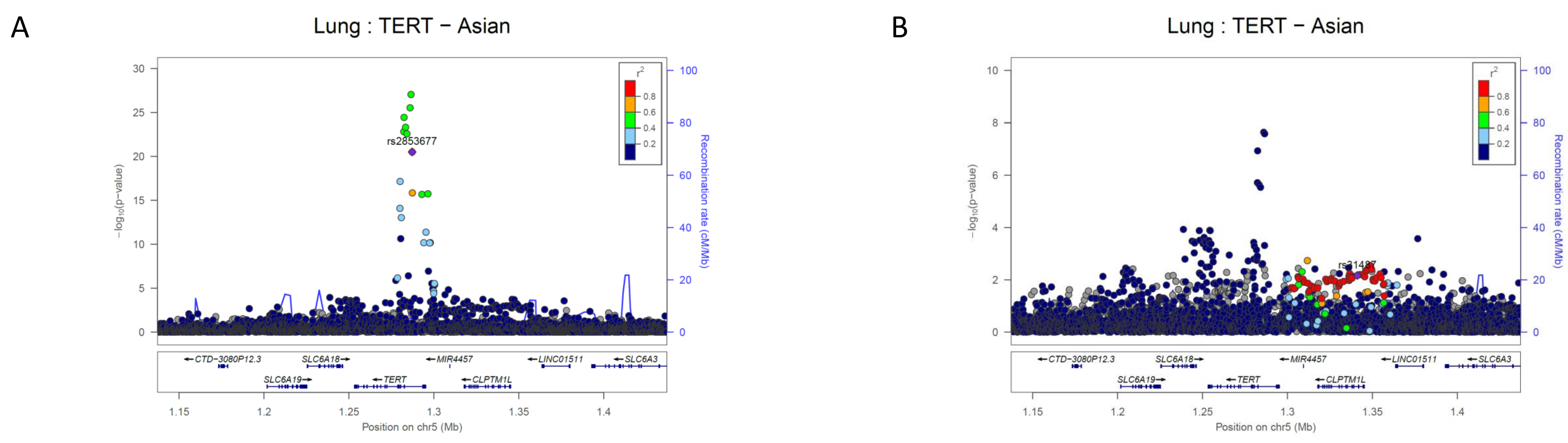


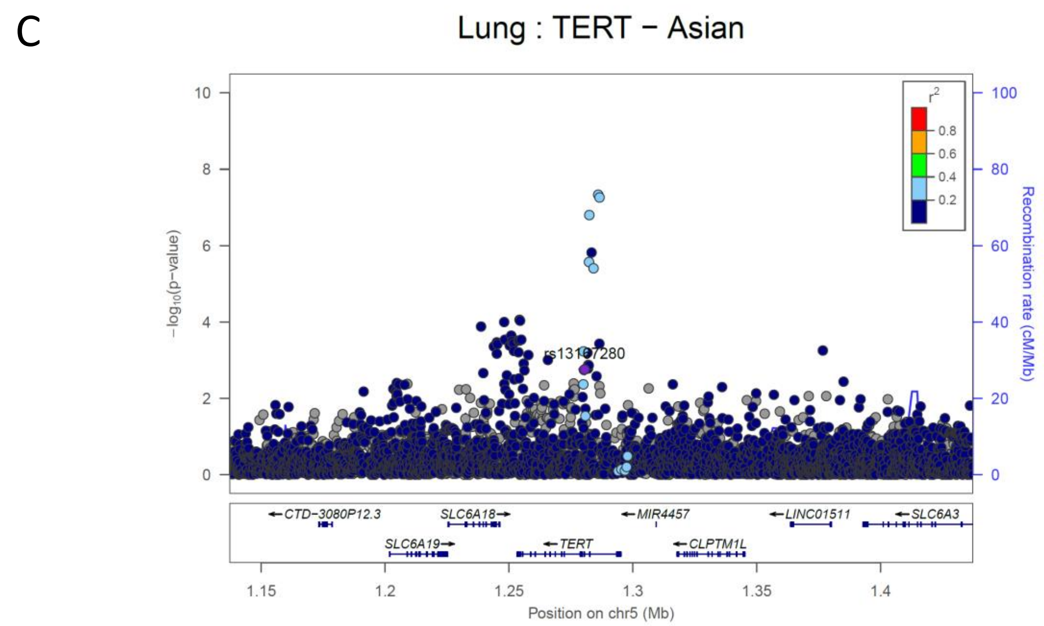


**A : Before Conditioning**
**B : After Conditioned rs2853677**
**C : After Conditioned rs2853677, rs31487**
**Note : Conditioned 1Mb loci**

A

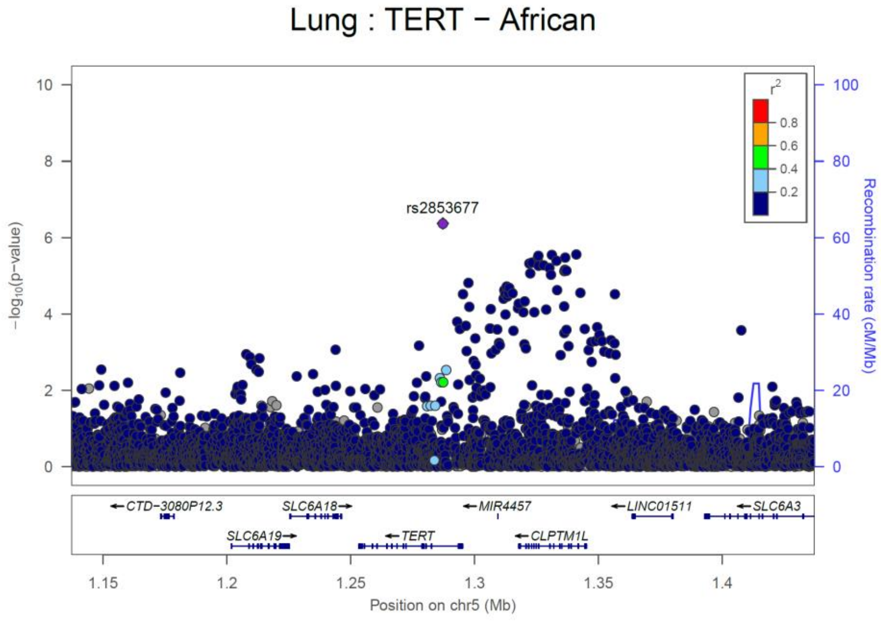


B

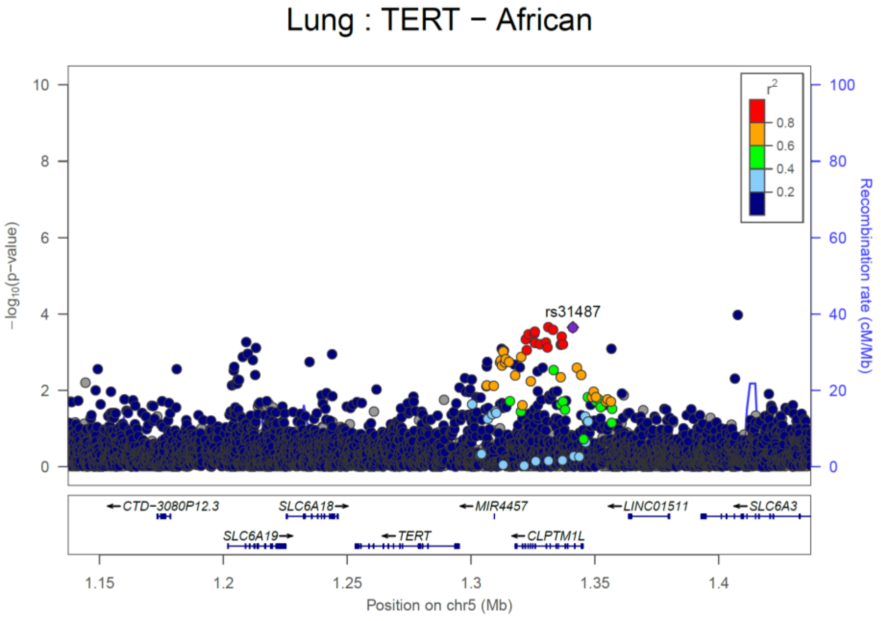


C

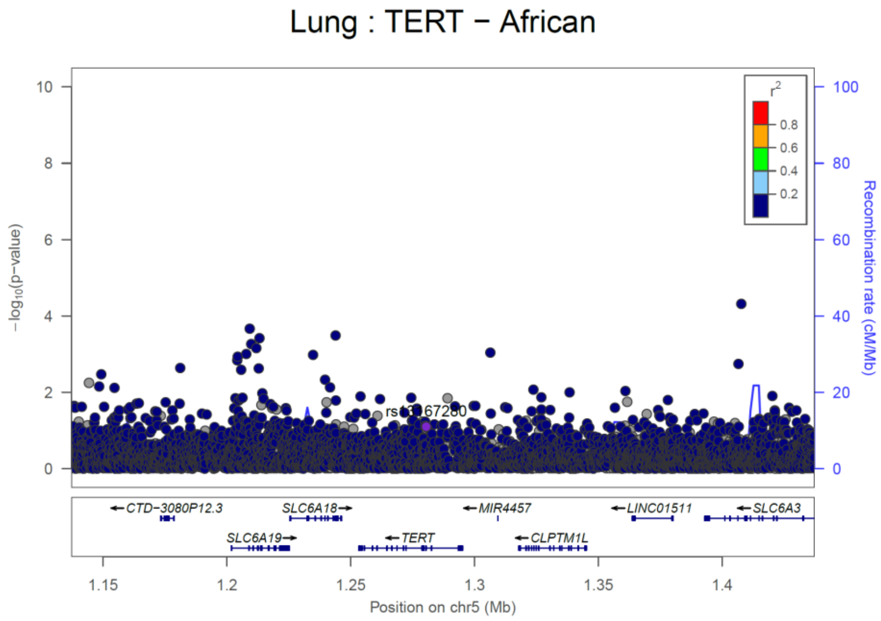


**A : Before Conditioning**
**B : After Conditioned rs2853677**
**C : After Conditioned rs2853677, rs31487**
**Note : Conditioned 1Mb loci**

**Title:** Trans-ethnic genome-wide meta-analysis of 35,732 cases and 34,424 controls identifies novel genomic cross-ancestry loci contributing to lung cancer susceptibility

### Supplementary Figure 4
### eQTL analysis of *IRF4* using GTEx/v8

### IRF (Interferon Regulatory Factor 4)

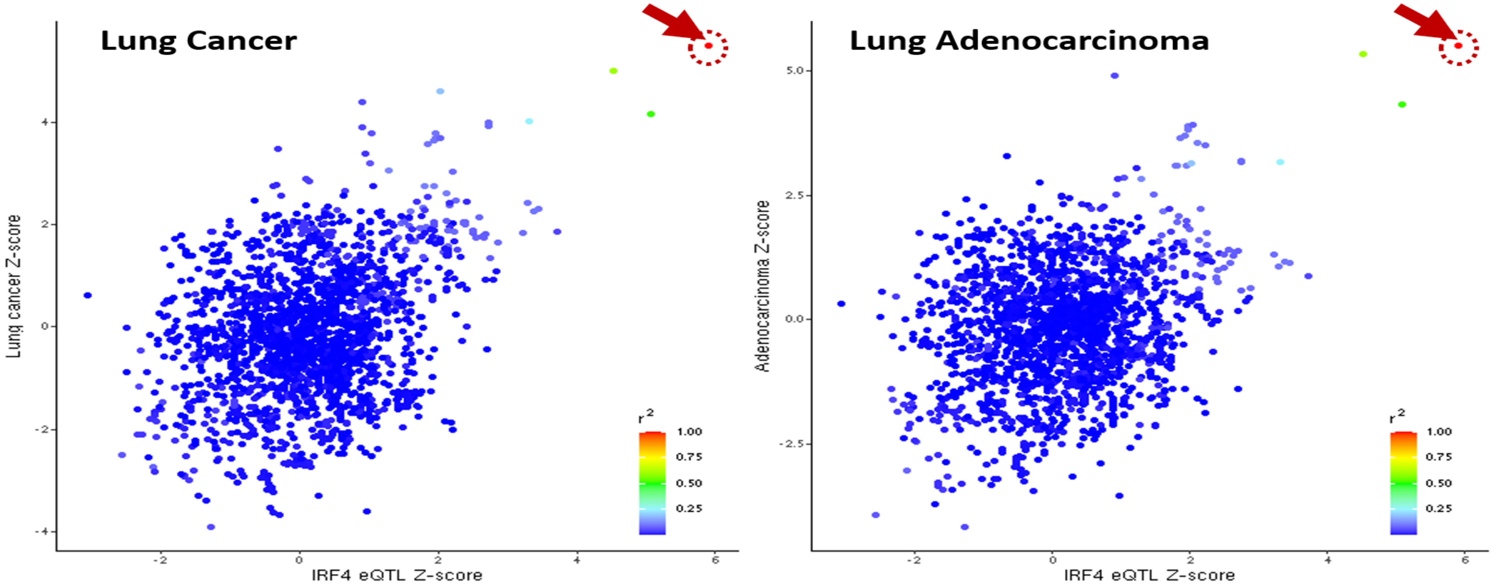


IRF4 (ENSG00000137265)

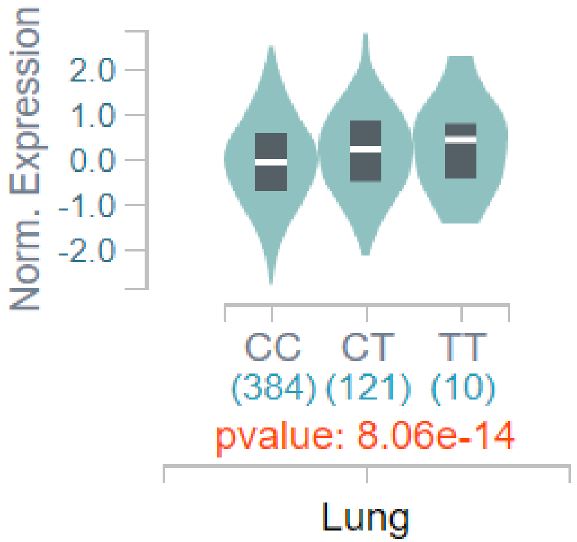

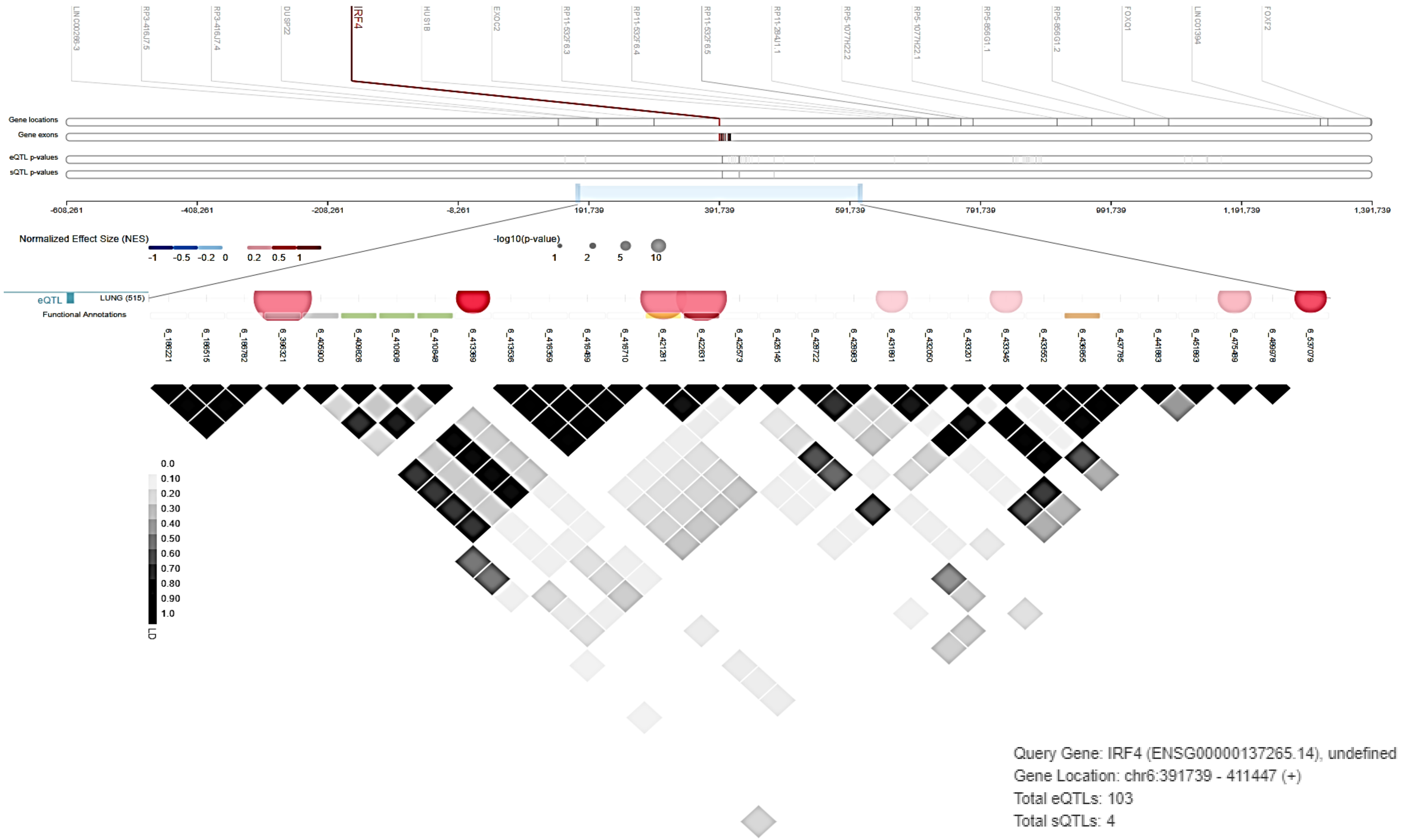
IRF4
Gene locations
Gene exons
eQTL p-values
sQTL p-values
-608,261
-408,261
-208,261
-8,261
191,739
391,739
591,739
791,739
991,739
1,191,739
1,391,739
Normalized Effect Size (NES)
-1 -0.5 -0.2 0
0.2 0.5 1
-log10(p-value)
1 2 5 10
eQTL
LUNG (515)
Functional Annotations
0.0
0.10
0.20
0.30
0.40
0.50
0.60
0.70
0.80
0.90
1.0
LD
Query Gene: IRF4 (ENSG00000137265.14), undefined
Gene Location: chr6:391739 - 411447 (+)
Total eQTLs: 103
Total sQTLs: 4

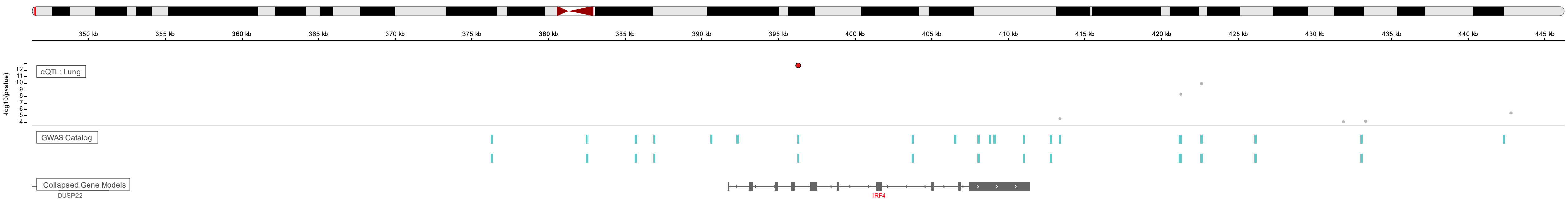


Isoform Expression of IRF4: ENSG00000137265.14 interferon regulatory factor 4 [Source:HGNC Symbol;Acc:HGNC:6119]

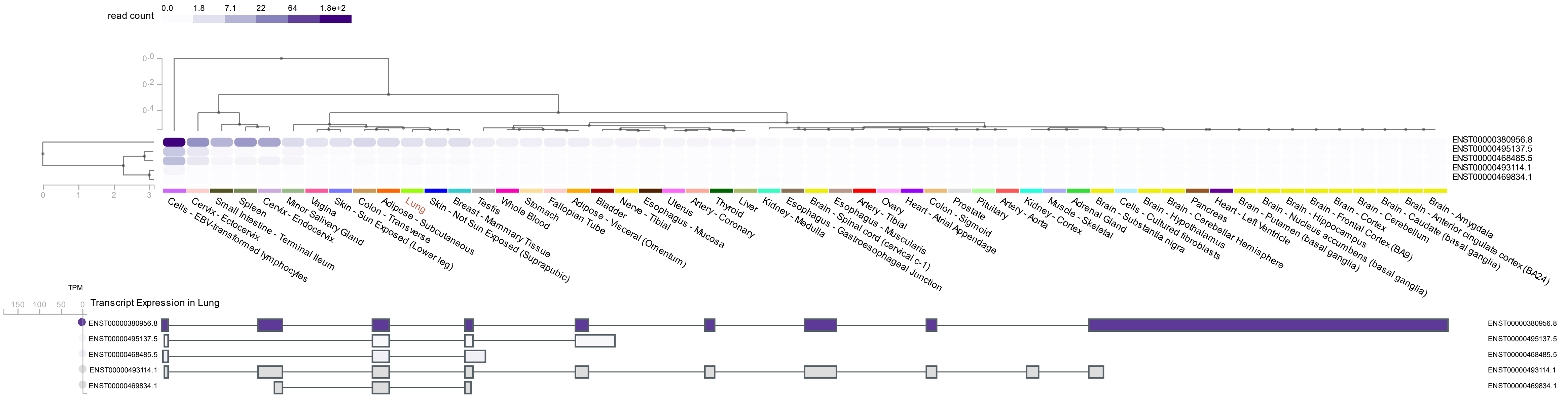

Exon Expression of IRF4: ENSG00000137265.14 interferon regulatory factor 4 [Source:HGNC Symbol;Acc:HGNC:6119]

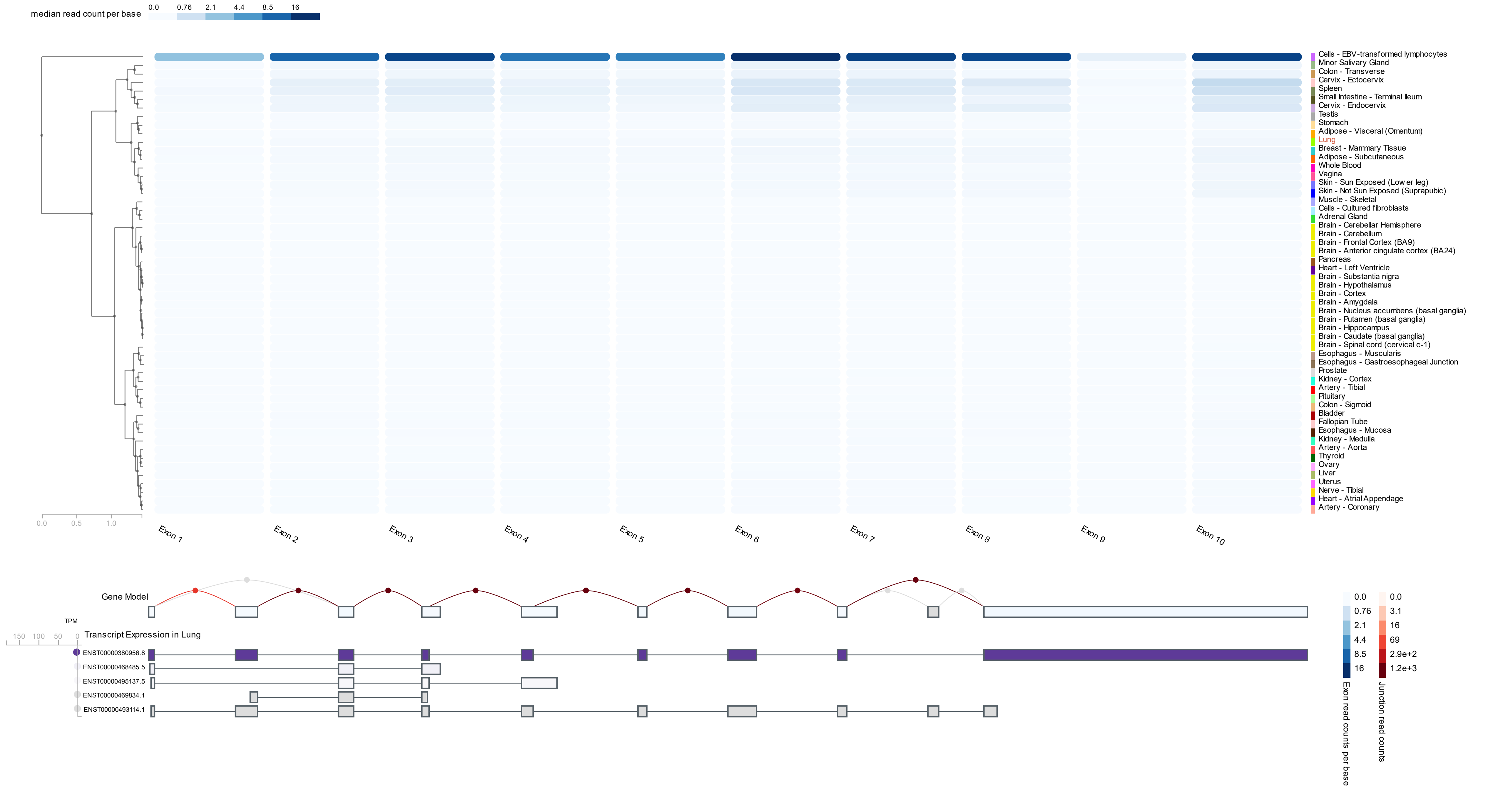

Junction Expression of IRF4: ENSG00000137265.14 interferon regulatory factor 4 [Source:HGNC Symbol;Acc:HGNC:6119]

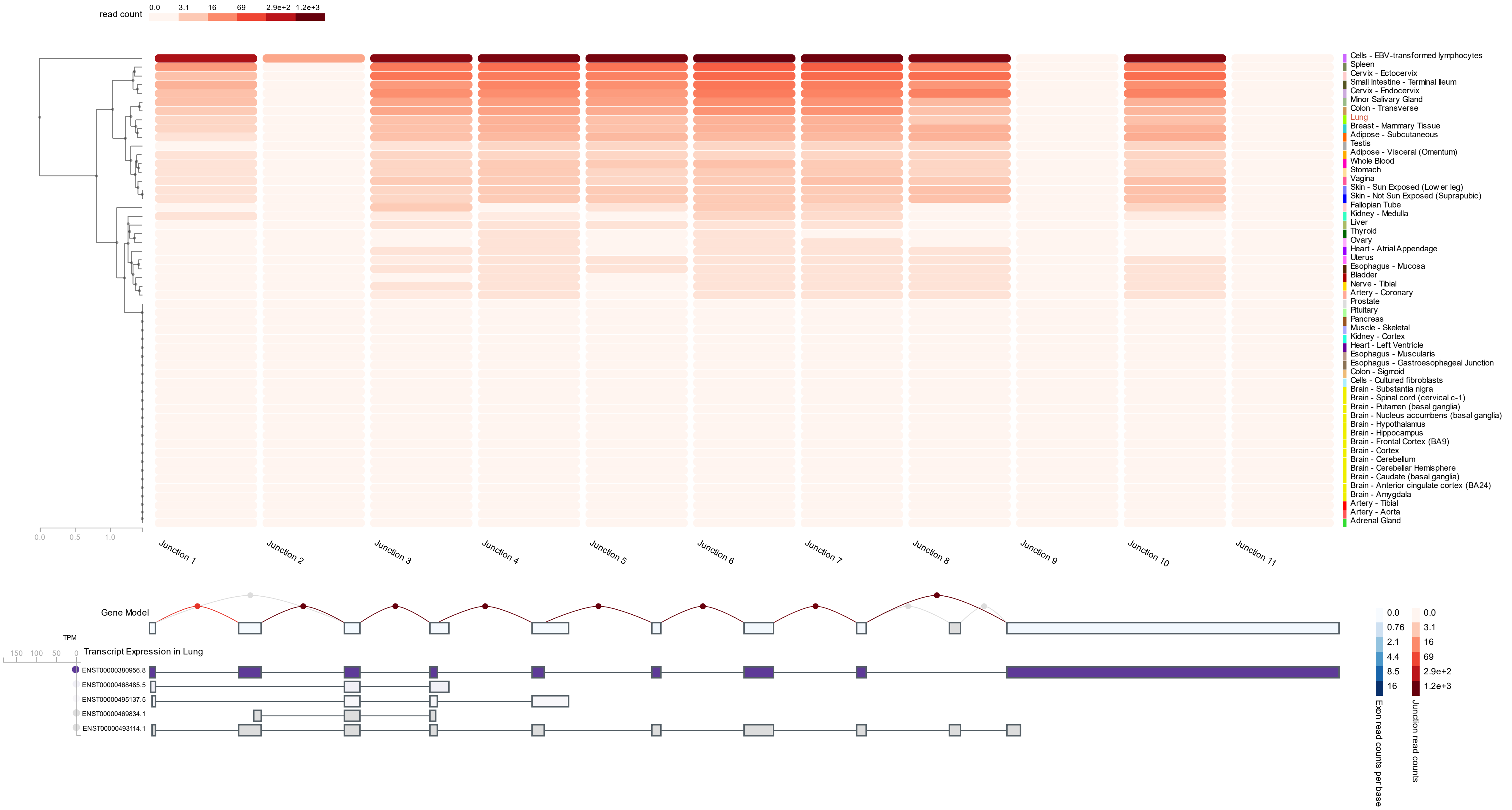

| Tissue | Samples | NES | p-value | m-value |
|---|---|---|---|---|
| Cells - EBV-transformed lymphocytes | 147 | 0.535 | 6.4e-6 | 1.00 |
| Minor Salivary Gland | 144 | 0.431 | 4.8e-5 | 1.00 |
| Testis | 322 | 0.356 | 3.1e-8 | 1.00 |
| Lung | 515 | 0.341 | 8.1e-14 | 1.00 |
| Pancreas | 305 | 0.333 | 8.1e-4 | 1.00 |
| Spleen | 227 | 0.328 | 1.5e-10 | 1.00 |
| Adipose - Visceral (Omentum) | 469 | 0.314 | 3.2e-6 | 1.00 |
| Nerve - Tibial | 532 | 0.310 | 1.6e-6 | 1.00 |
| Esophagus - Muscularis | 465 | 0.283 | 6.4e-5 | 1.00 |
| Prostate | 221 | 0.280 | 2.2e-3 | 0.999 |
| Vagina | 141 | 0.277 | 0.05 | 0.956 |
| Small Intestine - Terminal Ileum | 174 | 0.268 | 1.3e-6 | 1.00 |
| Ovary | 167 | 0.263 | 0.08 | 0.963 |
| Uterus | 129 | 0.247 | 0.05 | 0.972 |
| Whole Blood | 670 | 0.224 | 2.1e-12 | 1.00 |
| Artery - Coronary | 213 | 0.220 | 3.3e-3 | 0.997 |
| Heart - Left Ventricle | 386 | 0.210 | 0.01 | 0.995 |
| Colon - Sigmoid | 318 | 0.200 | 0.009 | 1.00 |
| Esophagus - Mucosa | 497 | 0.199 | 2.7e-6 | 1.00 |
| Thyroid | 574 | 0.194 | 3.1e-5 | 1.00 |
| Colon - Transverse | 368 | 0.180 | 9.1e-6 | 1.00 |
| Artery - Tibial | 584 | 0.177 | 1.7e-3 | 1.00 |
| Breast - Mammary Tissue | 396 | 0.173 | 3.9e-3 | 1.00 |
| Stomach | 324 | 0.172 | 2.0e-5 | 1.00 |
| Adipose - Subcutaneous | 581 | 0.164 | 8.2e-4 | 0.997 |
| Muscle - Skeletal | 706 | 0.158 | 0.009 | 0.990 |
| Artery - Aorta | 387 | 0.156 | 3.6e-3 | 0.994 |
| Brain - Substantia nigra | 114 | 0.155 | 0.3 | 0.856 |
| Brain - Spinal cord (cervical c-1) | 126 | 0.151 | 0.3 | 0.866 |
| Brain - Amygdala | 129 | 0.145 | 0.4 | 0.833 |
| Kidney - Cortex | 73 | 0.110 | 0.5 | 0.818 |
| Heart - Atrial Appendage | 372 | 0.109 | 0.2 | 0.862 |
| Pituitary | 237 | 0.109 | 0.2 | 0.837 |
| Esophagus - Gastroesophageal Junction | 330 | 0.0890 | 0.3 | 0.717 |
| Brain - Frontal Cortex (BA9) | 175 | 0.0760 | 0.6 | 0.751 |
| Skin - Not Sun Exposed (Suprapubic) | 517 | 0.0709 | 0.3 | 0.396 |
| Brain - Hypothalamus | 170 | 0.0658 | 0.6 | 0.667 |
| Adrenal Gland | 233 | 0.0580 | 0.6 | 0.654 |
| Liver | 208 | 0.0505 | 0.6 | 0.477 |
| Brain - Caudate (basal ganglia) | 194 | 0.0288 | 0.8 | 0.551 |
| Cells - Cultured fibroblasts | 483 | 0.00843 | 0.9 | 0.0150 |
| Brain - Anterior cingulate cortex (BA24) | 147 | - | - | - |
| Brain - Cerebellar Hemisphere | 175 | - | - | - |
| Brain - Cerebellum | 209 | - | - | - |
| Brain - Cortex | 205 | -0.0147 | 0.9 | 0.486 |
| Brain - Hippocampus | 165 | -0.0379 | 0.8 | 0.460 |
| Brain - Putamen (basal ganglia) | 170 | -0.0926 | 0.6 | 0.402 |
| Brain - Nucleus accumbens (basal ganglia) | 202 | -0.242 | 0.05 | 0.0560 |
| Skin - Sun Exposed (Lower leg) | 605 | -0.318 | 1.1e-7 | 0.00 |

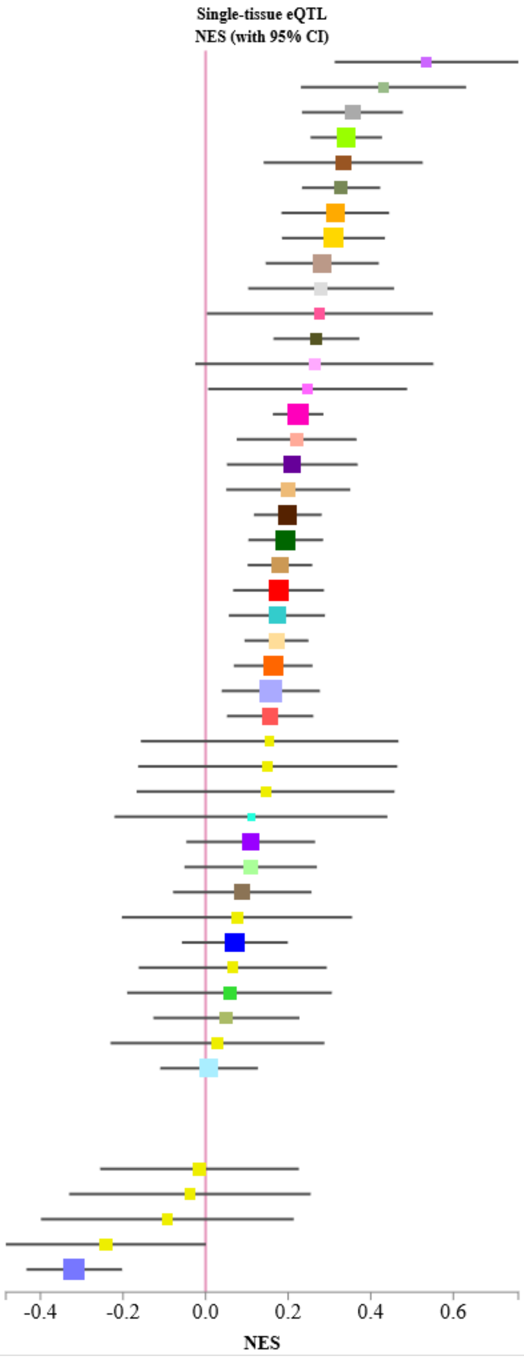


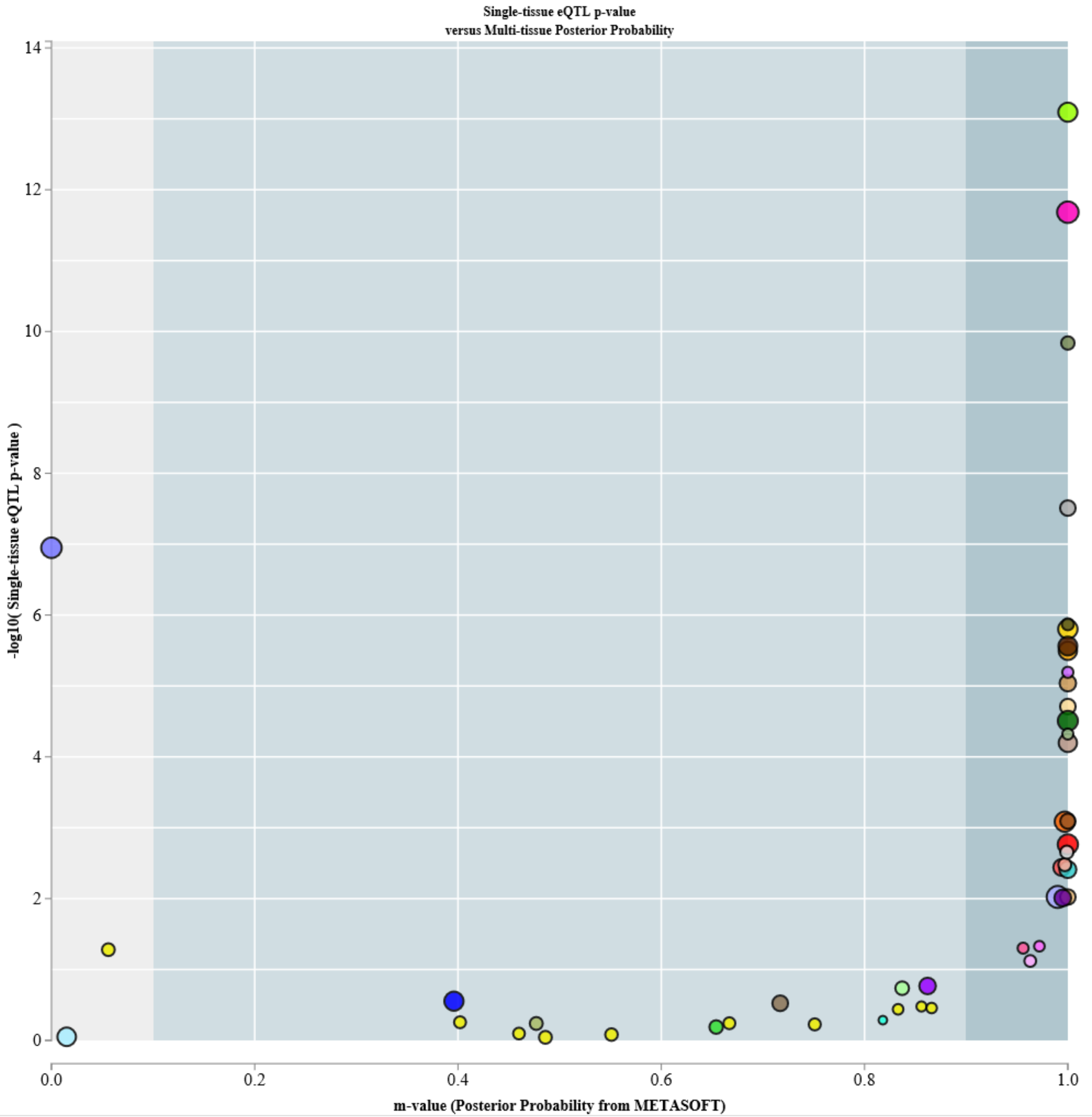

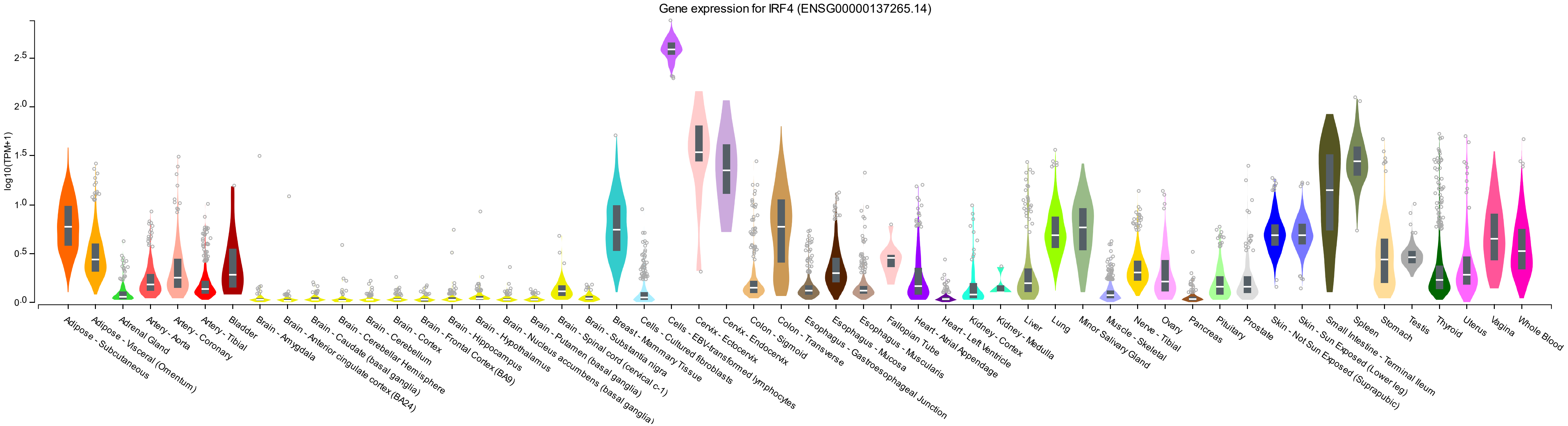
Gene expression for IRF4 (ENSG00000137265.14)
log10(TPM+1)
0.0
0.5
1.0
1.5
2.0
2.5
Adipose - Subcutaneous
Adipose - Visceral (Omentum)
Adrenal Gland
Artery - Aorta
Artery - Coronary
Artery - Tibial
Bladder
Brain - Amygdala
Brain - Anterior cingulate cortex (BA24)
Brain - Caudate (basal ganglia)
Brain - Cerebellar Hemisphere
Brain - Cerebellum
Brain - Cortex
Brain - Frontal Cortex (BA9)
Brain - Hippocampus
Brain - Hypothalamus
Brain - Nucleus accumbens (basal ganglia)
Brain - Putamen (basal ganglia)
Brain - Spinal cord (cervical c-1)
Brain - Substantia nigra
Breast - Mammary Tissue
Cells - Cultured fibroblasts
Cells - EBV-transformed lymphocytes
Cervix - Ectocervix
Cervix - Endocervix
Colon - Sigmoid
Colon - Transverse
Esophagus - Gastroesophageal Junction
Esophagus - Mucosa
Esophagus - Muscularis
Fallopian Tube
Heart - Atrial Appendage
Heart - Left Ventricle
Kidney - Cortex
Kidney - Medulla
Liver
Lung
Minor Salivary Gland
Muscle - Skeletal
Nerve - Tibial
Ovary
Pancreas
Pituitary
Prostate
Skin - Not Sun Exposed (Suprapubic)
Skin - Sun Exposed (Lower leg)
Small Intestine - Terminal Ileum
Spleen
Stomach
Testis
Thyroid
Uterus
Vagina
Whole Blood
