## Supplementary Note for "Trans-ethnic genome-wide meta-analysis of 35,732 cases and 34,424 controls identifies novel genomic cross-ancestry loci contributing to lung cancer susceptibility"

**Multi-Ethnic Lung Genome-Wide Association Study Subjects.** Trans-ethnic genome-wide meta-analyses were conducted in the twelve different studies consisting of Affymetrix Axiome Array Study (AFFY), the Female Lung Cancer Consortium in Asia (FLCCA), the Genetic Epidemiology of Lung Cancer (GELCC) Consortium, the Environment and Genetics in Lung cancer Etiology study (EAGLE), Helmholtz-Gemeinschaft Deutscher Forschungszentren Lung Cancer GWAS (GERMAN), the International Agency for Research on Cancer (IARC), the Institute of Cancer Research (ICR), MD Anderson Cancer Center Study (MDACC), NCI Lung Cancer and Smoking Phenotypes in African-American Cases and Controls (NCI), OncoArray Consortium Lung Study (OncoArray), the Prostate, Lung, Colorectal and Ovarian Cancer Screening Trial (PLCO) and Samuel Lunenfeld Research Institute Study (SLRI). All participants in this study signed an information consent, approved by local internal review board or ethics review committee and administered by trained personnel. The number of samples in the participating studies are individually described in the Supplementary Note Table 1.

**Supplementary Note Table 1. Sample description from 12 studies**

| STUDY | MISS | CONTROL | CASE | TOTAL |
|---|---|---|---|---|
| AFFY | 689 | 6,223 | 5,739 | 12,651 |
| FLCCA | 0 | 3,814 | 4,939 | 8,753 |
| GELCC | 18 | 796 | 757 | 1,571 |
| EAGLE | 0 | 1,977 | 1,923 | 3,900 |
| GERMAN | 0 | 473 | 455 | 928 |
| IARC | 0 | 2,441 | 1,841 | 4,282 |
| ICR | 0 | 1,666 | 1,395 | 3,061 |
| MDACC | 0 | 1,137 | 1,154 | 2,291 |
| NCI | 112 | 3,617 | 1,735 | 5,464 |
| ONCO | 13,766 | 19,363 | 23,348 | 56,477 |
| PLCO | 0 | 830 | 788 | 1,618 |
| SLRI | 0 | 495 | 330 | 825 |
| **TOTAL** | **14,585** | **42,832** | **44,404** | **101,821** |

**Supplementary Note Figure 1. Overview of the quality control procedures and the numbers of individual and variants excluded across the different quality control steps.**

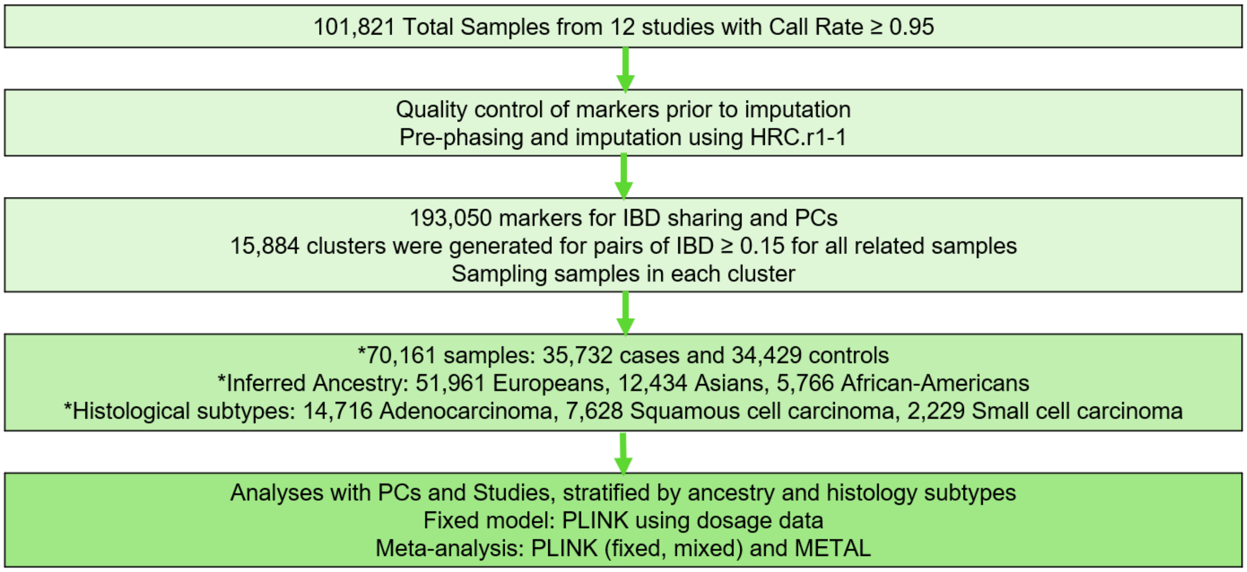


**Population Stratification.** We performed ancestry inference analysis using FastPop, a distance-based analysis with 505 HapMap samples with 2,042 ancestry informative markers. The colored points in grey indicate all 70,639 samples. Red, blue, and green denote HapMap samples with European(CEU), African(YRI), Asian(CHB) descent. Individuals were classified into 4 groups: European defined as >80% European descent, Asian as >40% Asian descent, African as > 20% African descent and others as not fulfilling any of the above criteria.

**Supplementary Note Figure 2. Inference of ancestry membership in multiple diverse populations using FastPop.**

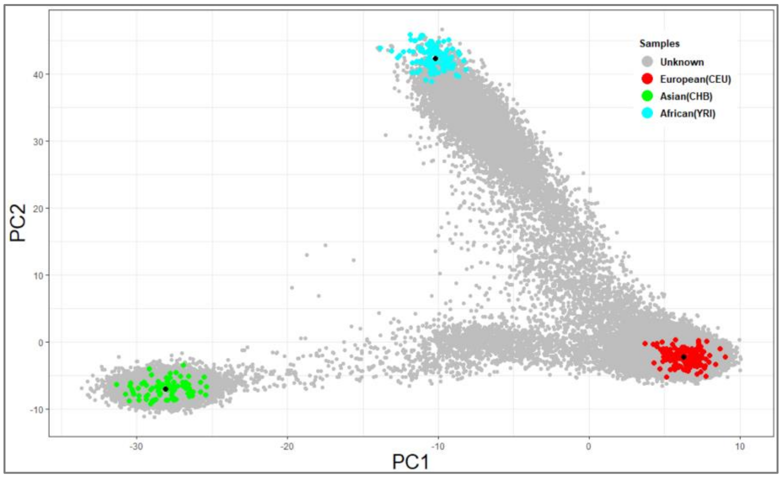

**Supplementary Note Table 2. Sample distribution in three intercontinental populations**

| STUDY | CONTROL | CASE | TOTAL |
|---|---|---|---|
| AFFY | 2,698 | 1,429 | 4,127 |
| FLCCA | 3,688 | 4,756 | 8,444 |
| GELCC | 789 | 754 | 1,543 |
| EAGLE | 1,972 | 1,919 | 3,891 |
| GERMAN | 471 | 420 | 891 |
| IARC | 2,441 | 1,838 | 4,279 |
| ICR | 1,627 | 1,366 | 2,993 |
| MDACC | 1,131 | 1,146 | 2,277 |
| NCI | 3,486 | 1,710 | 5,196 |
| ONCO | 15,683 | 20,153 | 35,836 |
| PLCO | 241 | 159 | 400 |
| SLRI | 437 | 325 | 762 |
| **TOTAL** | **34664** | **35,975** | **70,639** |

**Human cell lines and reagents.** MRC5-SV40, a human lung fibroblasts derived cell line was maintained in standard Dulbecco's modified Eagle's medium with 10% fetal bovine serum, 2 mM L-glutamine, 100µg/mL penicillin, and 100 µg/mL streptomycin as described in [1]. Cell line was authenticated by ATCC STR analysis and routinely check to be mycoplasma-free.

Full-length ACTR2, NECTIN1, ZFP42, PPIL6 and PPP2R5E entry clones for gateway cloning was synthesized, sequence-verified, and cloned into pDONR223 (Invitrogen) by Genscipt. RORA and IRF4 entry clones were acquired from the Kenneth Scott cDNA ORF library with the accession number: IOH43547 and ccsbBroadEn06459 respectively. All above clones were further subcloned into an N terminal GFP tagged vector (pcDNA6.2/N-EmGFP-DEST, Invitrogen), using Gateway LR Clonase II Enzyme Mix (Invitrogen). Overexpression plasmids transfections were performed using GenJet In Vitro DNA Transfection Reagent Ver. II (# SL100489, SignaGen).

Non-targeting pool siRNA (D-001810-10) and SMARTpool siRNAs each containing four targeting sequences of RORA(L-003440-00-0005), IRF4(L-019668-00-0005), ZFP42(L-017752-02-0005), PPIL6 (L-009111-00-0005), ANKRD33B (L-039238-01-0005) , WNK1(L-005362-02-0005), PPP2R5E (L-008531-00-0005), Nectin1 (L-04300-00-0005) and ACTR2 (L-012076-02-0005) were purchased from Dharmacon and siRNA transfections were carried out with lipofectamine RNAiMax Transfection Reagent (#13778075, Invitrogen), following the manufacturer's recommendations. SMARTpool ON-TARGET*plus* siRNA were designed and modified for greater specificity and reduce off-targets up to 90% utilizing a dual-strand modification.

**RT-qPCR.** Knockdown efficiency was quantified by real-time quantitative reverse transcription PCR (RT-qPCR) and shown in Supplemental Note Figure 3. RNeasy mini kit (Qiagen #74106) was used to extract total RNA from cells 72 hours post siRNA transfection. 300 ng of total RNA from each sample was used to synthesize cDNA by the Superscript III first strand synthesis system (Invitrogen, #18080051). qPCR reactions were performed using iTaq Universal SYBR Green Supermix (BioRad #172-5121) on a QuantStudio 3 Real-Time PCR System (Applied Biosystems). For each gene, three replicates were analyzed and the average threshold cycle (Ct) was calculated. The relative expression levels were calculated with the 2–ΔΔCt method [2]

Primers used included GADPH forward: CAA TGA CCC CTT CAT TGA CC; GADPH reverse: GAT CTC GCT CCT GGA AGA TG; RORA forward: CAC GAC GAC CTC AGT AAC TAC A; RORA reverse: TGG TGA ACG AAC AGT AGG GAA; IRF4 forward: GCG GTG CGC TTT GAA CAA G; IRF4 reverse: ACA CTT TGT ACG GGT CTG AGA; ZFP42 forward: GCC TTA TGT GAT GGC TAT GTG T; ZFP42 reverse: ACC CCT TAT GAC GCA TTC TAT GT; PPIL6 forward: TGC ACT TTA TGA CGC ACT CAC; PPIL6 reverse: TGT CCA AAA ACA CGA AAT CAT GC; ANKRD33B forward: GAC CCC GCT GAC TAC GAA G; ANKRD33B reverse: GGG TAG AAA GAG TCG TCC GAG; WNK1 forward: TGG TAG AGT CTG GGT ATG TCT G; WNK1 reverse: GAA GCA GTA GGT ATG CCG GTG; PPP2R5E forward: CAG AGG CTG TTT GAC AGA GCA; PPP2R5E reverse: TCA CTA GGA GGG AGA GTT CTG A; NECTIN1 forward: TCT ACA TCT GCG AGT TTG CTA CC; NECTIN1 reverse: CTT CTT GGC TCG AAG CAC TG; ACTR2 forward: CAC CTG TGG GAC TAC ACA TTT G; ACTR2 reverse: TGG TTG GGT TCA TAG GAG GTT C.

**Supplementary Note Figure 3. Validation of mRNA siRNA knockdown efficiency by RT-qPCR analysis. 9 candidate genes tested for DNA damage were knocked down by siRNA. n=3, mean ± S.E.M.**

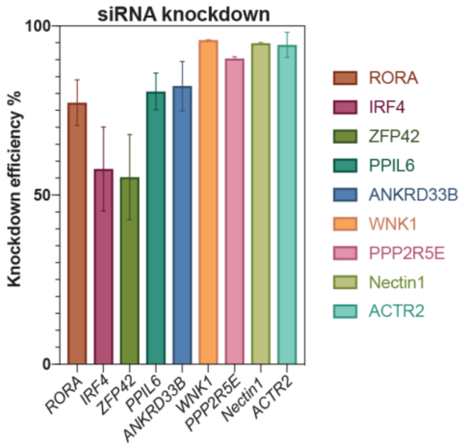


**Flow-cytometric DNA damage assays.** Sensitive DNA damage assays by flow cytometry were performed as previously described. γH2AX primary antibody (Sigma, Catalog #05-636) and goat anti-mouse secondary antibody, Alexa Fluor 647 (Thermo Fisher, Catalog #A21236) were used to stain cells. Stained cells were then analyzed by a BD LSRFortessa flow cytometer. FCS files were analyzed by FlowJo 10.5 software. For siRNA experiments, cells were collected 72 hours post transfection and 0.5% of the mock cells were gated as the γH2AX threshold as previously demonstrated. The percentage of γH2AX positive cells in each sample was calculated and compared to its corresponding non-targeting siRNA control.

For overproduction experiments, mock-transfected cells were used to set the gates to determine the GFP and γH2AX positive cells. 0.5% of the mock cells were gated as the γH2AX threshold. The DNA-damage ratios by protein overproduction for 72h are calculated as described. Briefly, the damage ratio is defined as (Q2/Q3)/(Q1/Q4), where Q2 is the portion of transfected γH2AX-positive cells; Q3 is the portion of transfected, γH2AX -negative cells; Q1 is the portion of untransfected, γH2AX-positive cells; and Q4 is the portion of untransfected, γH2AX-negative cells. The DNA damage ratios by candidate protein overproduction were compared with GFP-Tubulin as previously described.
